# Epidemiology of traumatic brain injury in South Asia: A Systematic Review

**DOI:** 10.1101/2025.09.26.25336780

**Authors:** Shahriar Hasan, Shudeshna Chakraborttye Purba, Rifat Hannan, Md. Ekramul Hasan, Fazzarna Shithi, K M Yasin Al Amin, Md. Masum Mrida, Joynal Abedin Imran

**Author notes:** **Corresponding author:** (SH).

## Abstract

**Background:** Traumatic brain injury (TBI) is a serious global health problem contributing significantly to disability and death. No systematic review existed on epidemiology of TBI in South Asia, where accurate data were essential for planning healthcare policy and injury prevention programs.

**Methods:** This systematic review adhered to PRISMA guideline and received registration from PROSPERO (CRD42022364511). This review included observational from 1 January 1950 to 28 December 2024. A systematic search was performed on PubMed, Scopus, and Google Scholar and included studies fulfilling predetermined criteria. The methodological quality of the included studies was evaluated by Methodological Evaluation of Observational Research (MORE) checklist and result were presented as narrative synthesis.

**Findings:** Analysis of 130 studies reported a high incidence of TBI in South Asia primarily from road traffic accidents (RTAs). Prevalence ranged from 8.4% to 95.9%, with elevated case fatality and mortality rates. RTAs, falls and assaults were the leading cause of severe TBI. Adult males aged 21-30 years showed highest risk.

**Conclusion:** The results highlighted urgent necessity for standardized case definitions, improved data collection, and strengthened healthcare capacity in South Asia. Further research is required to understand long-term consequences and guide evidence-based public health responses.

## Introduction

TBI is a major contributor to disability and death worldwide,^1^ and over 60 million individuals suffering from a TBI every year.^2^ The individuals surviving a TBI commonly experience prolonged physical, cognitive, and psychosocial disabilities.^3^ The treatment of TBI is complicated and costly, and its cost ranged throughout a person’s lifetime from $279 million to $1.22 billion.^4^

TBI is generally characterized as mild, moderate, and severe based on the Glasgow Coma Scale (GCS), a major prognosis predictor.^5^ However, novel approaches have been proposed for further precision in rating TBI severity.^6^ Newer tools, including advanced neuroimaging and biomarkers, are increasingly being combined with GCS scores.^7,8^

The epidemiology of traumatic brain injury (TBI) in South Asia is an emerging concern and rising rates have been documented in various countries in the region.^9^ Traumatic brain injury is an increasingly urgent public health problem due to urbanization, rising motorization, and poor safety infrastructure.^10,11^

South Asian traumatic brain injury (TBI) is diverse based on road infrastructure, economic conditions, and gaps in policy. Road traffic accidents (RTAs) are primarily responsible for causing it, with Bangladesh (62%), India (56%), and Nepal (59%) being heavily impacted.^12–14^

South Asian rates of traumatic brain injury (TBI) vary based on data collection differences, case definitions, and economic circumstances. Higher-income countries have superior data quality relative to lower-and middle-income countries (LMICs), resulting in more precise estimates.^15,16^

Inconsistent case ascertaining makes comparison challenging,^17,18^ whereas socioeconomic differences influence incidence and mortality.^16,19^ Standardizing case definitions and data collection using Common Data Elements (CDEs) enhances research quality as well as comparison.^20,21^ Standard practices and an improvement in healthcare infrastructure need to be instituted for improved management of TBI in South Asia.

High-quality observational research is essential for the understanding of TBI. Observational studies such as CENTER-TBI yield clinical and demographic data.^22^ TRACK-TBI and PRECISION-TBI biomarker analyses support individualized treatment.^23,24^ Observational studies contrast treatment outcomes^25^ and monitor recovery over the long term.^26^ Reliable and complete epidemiological data on South Asian TBI is vital for evidence-based planning at the level of public health as well as clinical management.^14^

Rapid urbanization and motorization and rising road traffic injuries resulted in a rising burden of TBI in the region, however available data remain fragmented and inconsistent. Recent advancements in healthcare infrastructure and the rising volume of observational studies conducted in the last few decades created a window of opportunity for a systematic review of existing evidence. Also, the absence of a previous systematic review on the subject points to an important gap in understanding the actual extent and magnitude of TBI in South Asia. With the pressure on public health systems to manage resources efficiently rising by the day, aggregating epidemiological data at present can be used to establish targeted prevention strategies. It can enhance trauma care and research directions for the future. This review therefore comes at a timely and imperative moment to facilitate policy-making informed by evidence and enhance clinical outcomes in the region. Our study addresses the following research questions:

1. What is the prevalence of traumatic brain injury in South Asia?
2. What is the mortality and case fatality of traumatic brain injury in South Asia?
3. What is the severity and mechanism of injury of traumatic brain injury in South Asia?
4. What are the age and sex distribution of traumatic brain injury in South Asia?

## 4. Methods

### Registration and Protocol

This systematic review was reported and executed according to the Preferred Reporting Items for Systematic Reviews and Meta-Analyses (PRISMA) Statement.^171^ The protocol for this systematic review was registered on PROSPERO (registration number 2022: CRD42022364511). It can be accessible at https://www.crd.york.ac.uk/prospero/display_record.php?ID=CRD42022364511

### Search strategy and inclusion criteria

The databases searched included PubMed, Google Scholar and Scopus from January 1, 1950 through December 28, 2024. For all of them, combined key words and MeSH terms (S1 Appendix) were used for searching. To minimize publication bias risk, reference lists in included studies also were searched. The studies included were observational in nature, encompassing cross-sectional, case-control, and cohort designs. Studies providing retrospective and prospective descriptive data on the epidemiology of TBI in South Asia were included. To qualify for this research, studies also had to be an original study, have measured and reported on prevalence, mortality, case fatality, severity, and mechanism of injury of TBI in South Asia. There were no exclusions based on study size as data from hospitals, police records, relatives/persons accompanying them were included. Also, there were no exclusions based on dates of data, study performance and publication dates, age of participants, nor based on TBI severity.

For certain of those terms, including TBI, incidence, mortality, and case fatality, definition among authors was varied. For the purposes of this research, these terms were defined in the following ways:

⍰ **TBI:** An injury to the head by blunt or penetrative injury causing enough harm so that the patient experiences a change in brain function; alternatively, more recently, as a change in brain function, or other indication of brain pathology due to an external pressure.^27^ Ascertainment of a case of TBI might be through any one of the following: clinical diagnosis based on history and physical examination and imaging (CT scan, X-ray scans, MRI), autopsy, Glasgow coma scale (GCS), CDC criteria, ICD 9th and 10th revision codes and others.
⍰ **Case fatality:** Proportion of people with TBI who subsequently died due to a cause related to the TBI at certain time-points.
⍰ **TBI severity:** Categories of severity (severe, moderate, mild), as defined by the GCS or other classification system used by the authors.^5^

### Exclusion criteria

Exclusion of studies were based on numerous criteria. First, those not from South Asia were excluded in order to be regionally relevant. Non-primary research studies, reviews, meta-analysis, and grey literature were excluded in order to secure primary data of observational studies. Studies without data on key outcomes, prevalence, mortality, case fatality, severity, or mechanism of injury were excluded. Those studies with inconsistent TBI case definitions or poor ascertainment methods were also excluded in order to ensure reliability. Studies with poor methodical reporting or ambiguous sampling and diagnostic requirements were also excluded in order to secure quality. The above exclusions ensured only studies of relevance, reliability, and quality were included in this review.

### Screening and study selection

SH and SCP completed the initial screening of titles and abstracts to select studies that met inclusion criteria. Non-eligible studies were excluded and included studies were selected for full-text review. RH and MEH independently assessed study methodologies and reported results from full-texts to ensure eligibility. Disagreements were resolved through consensus and discussion.

### Data Analysis

FS and KMYA extracted data from included studies using a standard data extraction form extracting study features, participant information, methodological details, and outcome data. MMM and JAI checked extracted data for accuracy and completeness by referencing against source articles. Discrepancies were discussed and resolved jointly. SH, SCP, RH, FS, KMYA, MMM, and JAI synthesized extracted data into an integrated narrative overview centered on primary epidemiological variables. They made sure that synthesis was thorough and all pertinent data were included.

SH, RH, MMM and JAI evaluated included study quality. The methodological quality of included studies was assessed using the Methodological Evaluation of Observational Research (MORE) checklist that has been utilized in an earlier systematic review.^17^ The checklist evaluated a number of areas related to bias or quality, including study aim, funding of study, conflict of interest, ethical clearance, study design, sampling, case definition, bias addressed, data sources, reliability of estimates, incidence and mortality. Each area was ranked according to specific criteria and scored as “OK, Minor Flaw, Major Flaw, or Poor reporting” (Appendix 2). No studies were removed from this research based on their methodological quality.

Narrative synthesis was written on this review findings. All data on prevalence, mortality, case fatality rates, severity and mechanism of TBI as well as important demographic information on age and sex distribution were extracted. Quantitative data on TBI outcomes were synthesized through categorization of results by prevalence rates, mortality and case fatality rates and severity classification (mild, moderate and severe) and mechanism of injury (e.g., road traffic accidents, falls and assaults).

### Ethical Considerations

This study is a systematic review of previously published data. All data were extracted from publicly available literature. Therefore, institutional review board approval and direct patient consent were not required for this study. The included primary studies were evaluated for their reporting of ethical approval. As this is a systematic review, it is not a clinical trial and does not require registration in a clinical trial registry. The review protocol was registered with PROSPERO.

## 5. Results

A total of 1347 articles were identified. After removing duplicates, 1122 were screened on title and abstract and 893 were excluded. Subsequently, 229 papers were screened for full-text eligibility, with 130 articles finally included in this review (Fig. 1).

**Fig. 1.**
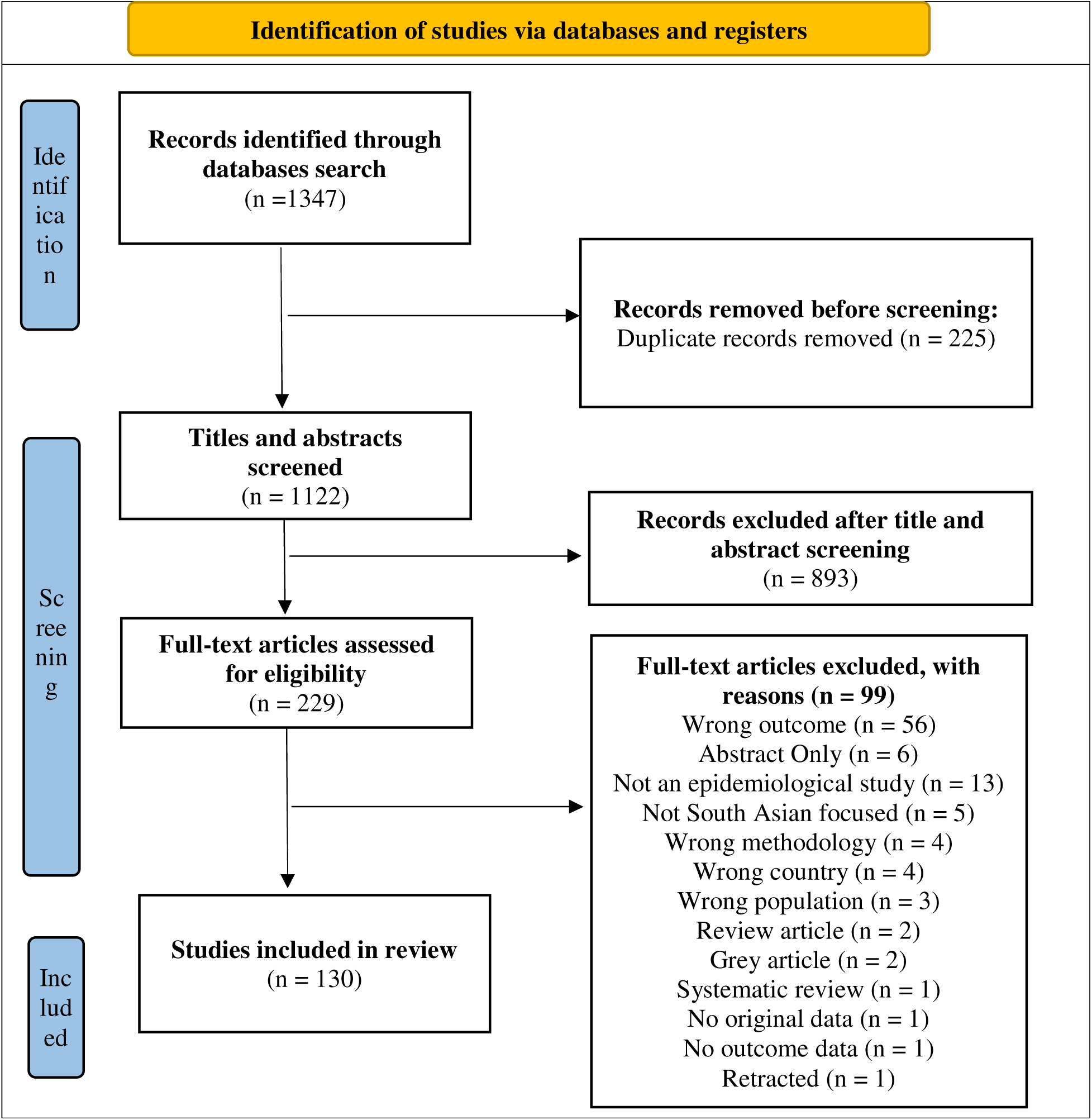
Preferred Reporting Items for Systematic Reviews and Meta-Analyses (PRISMA) flowchart of the study selection process

**Table 1.**
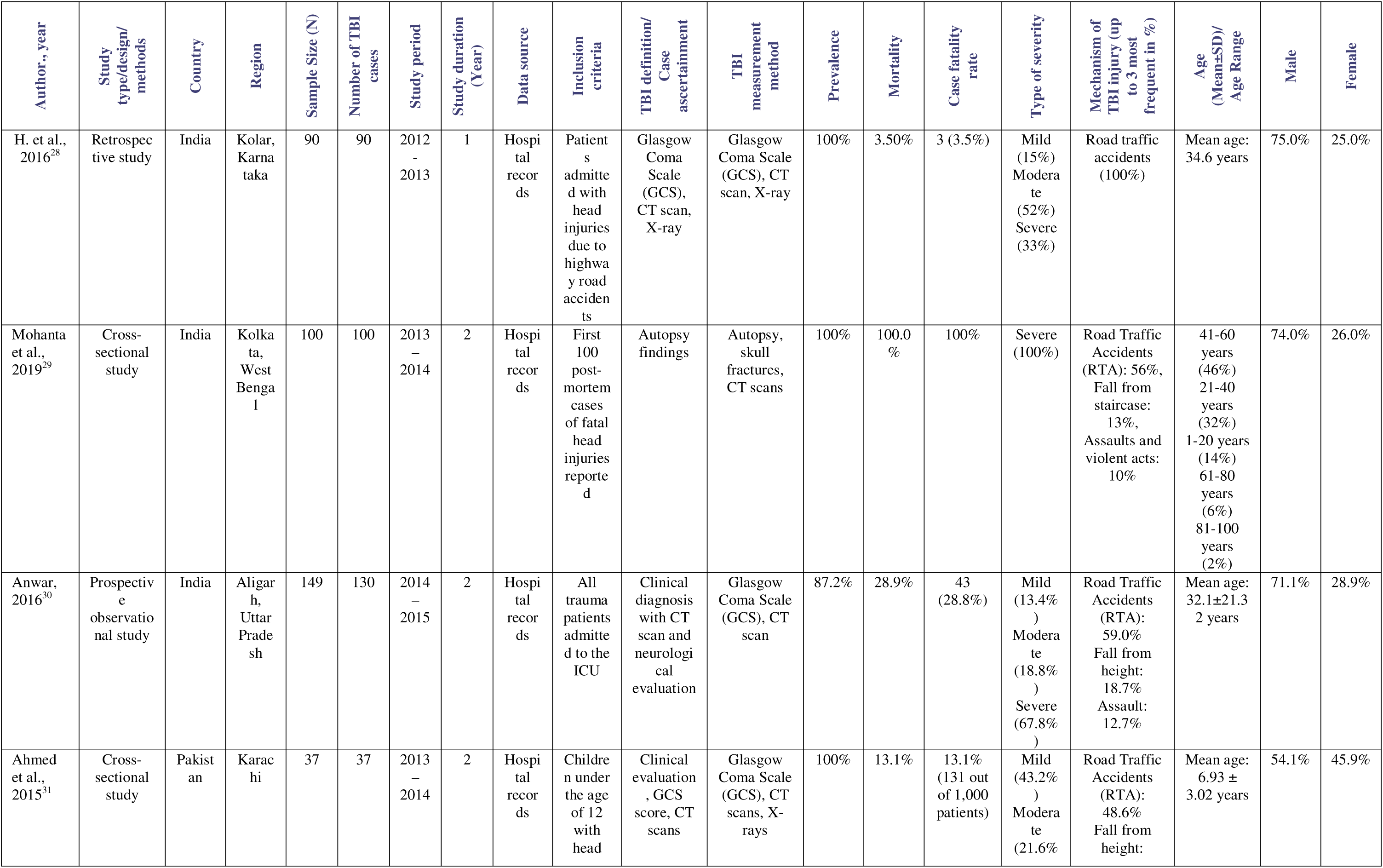
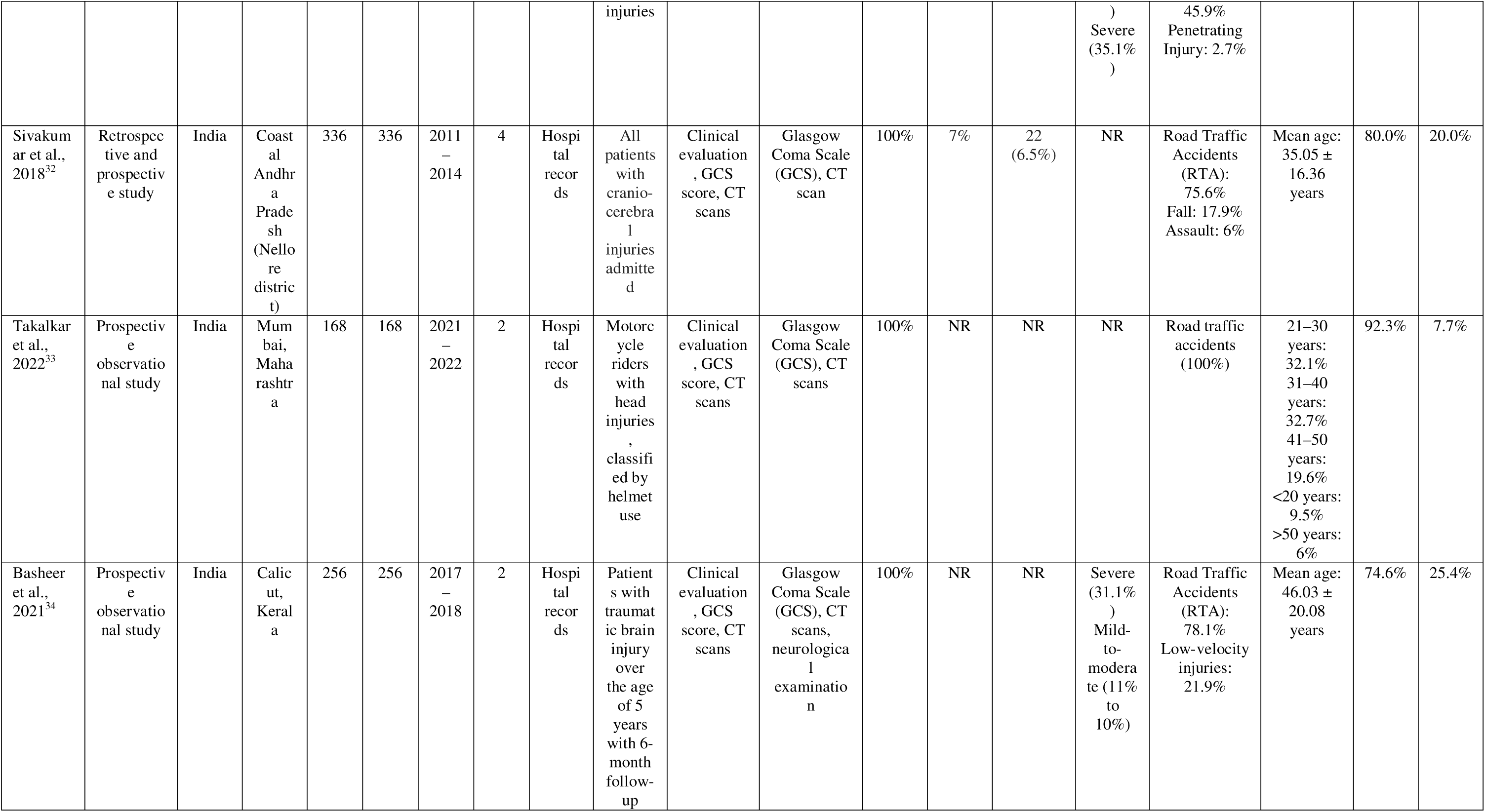
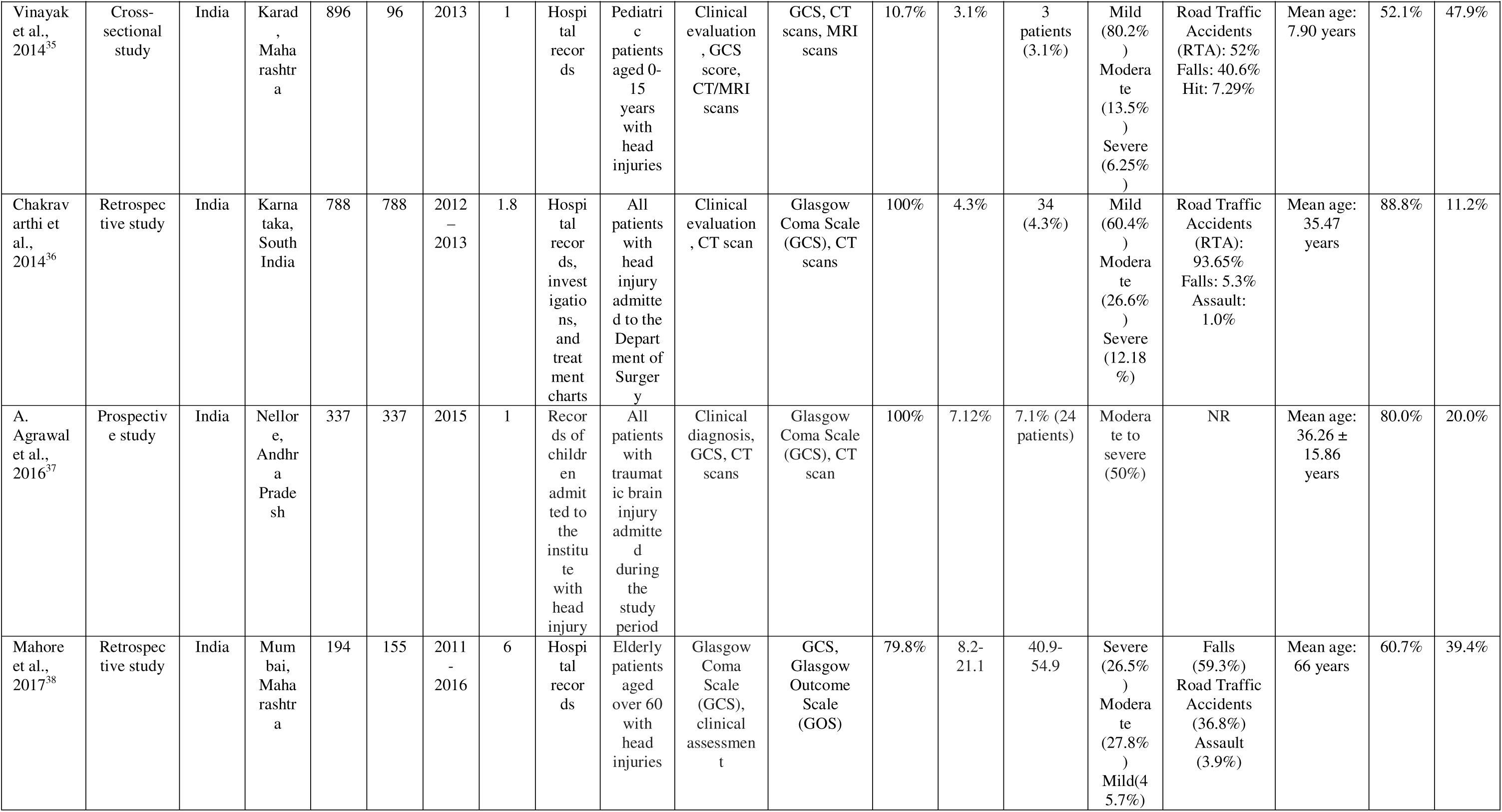
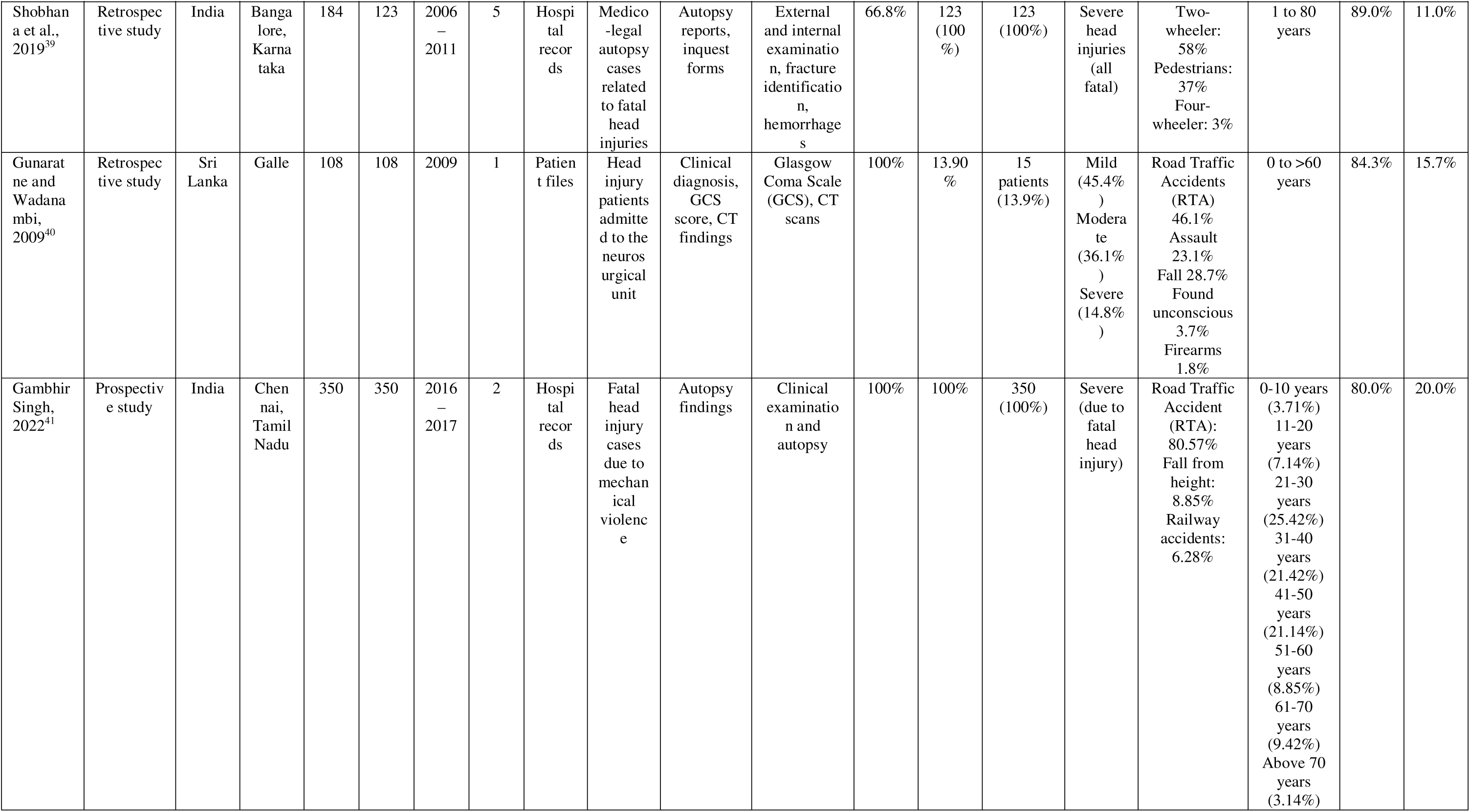
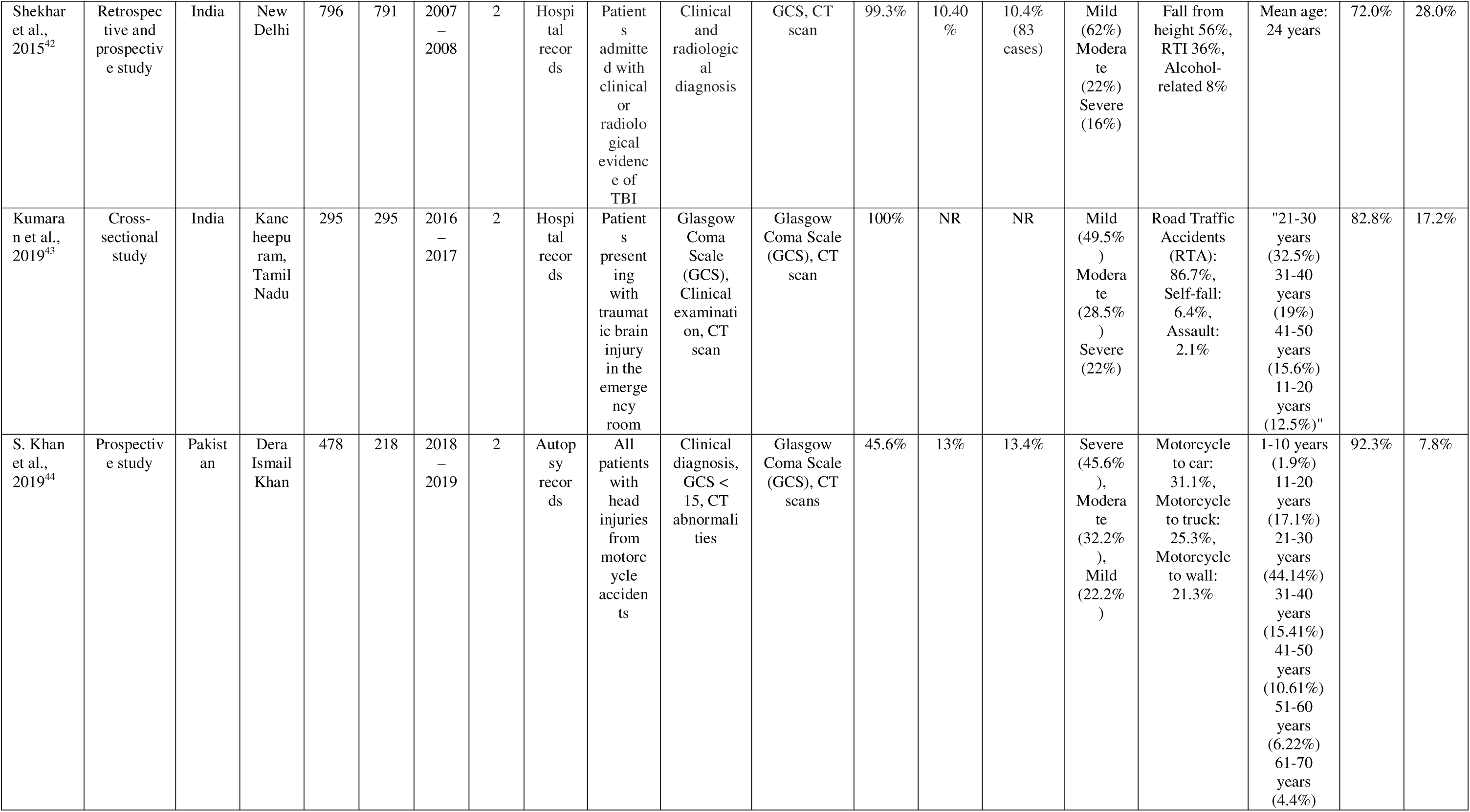
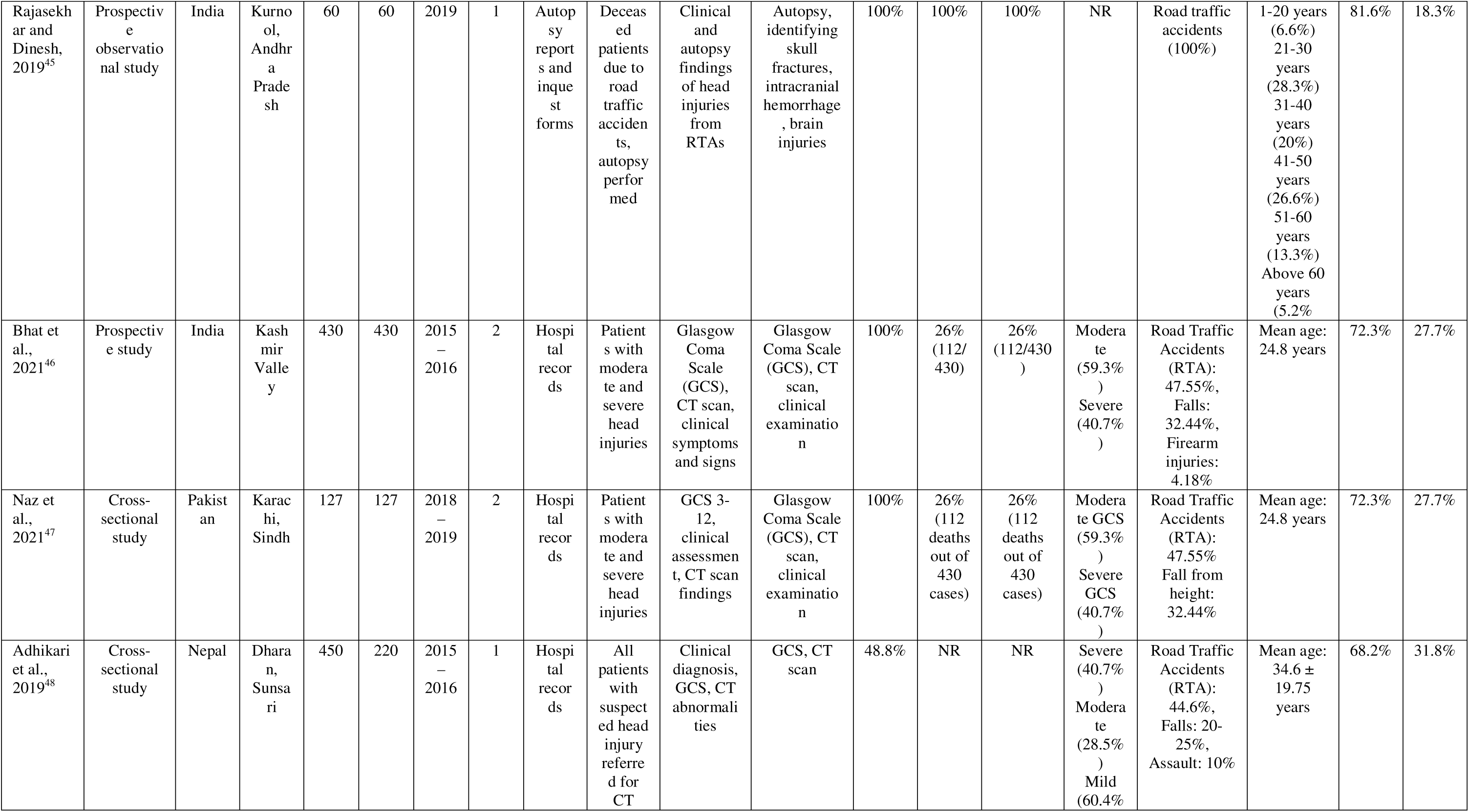
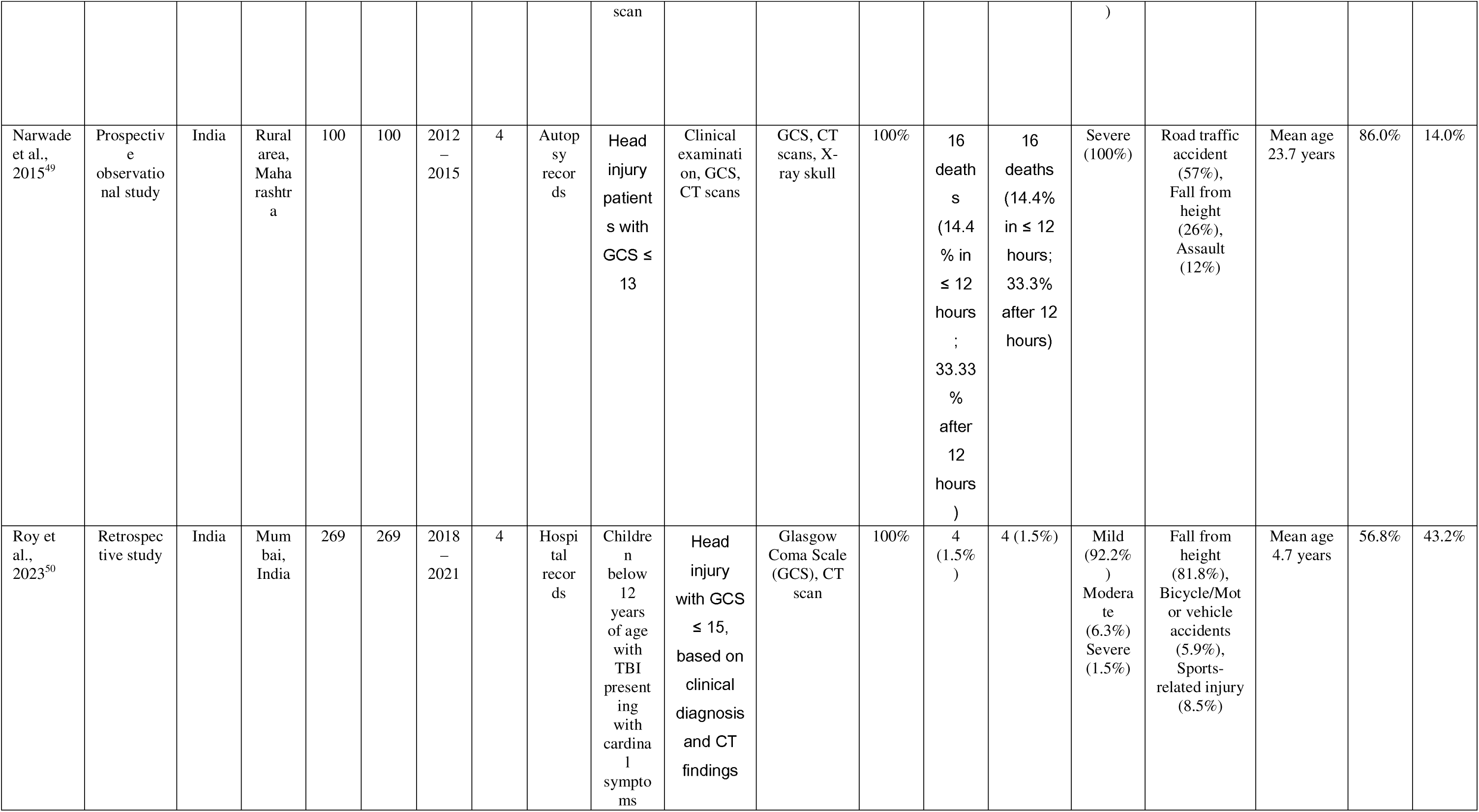
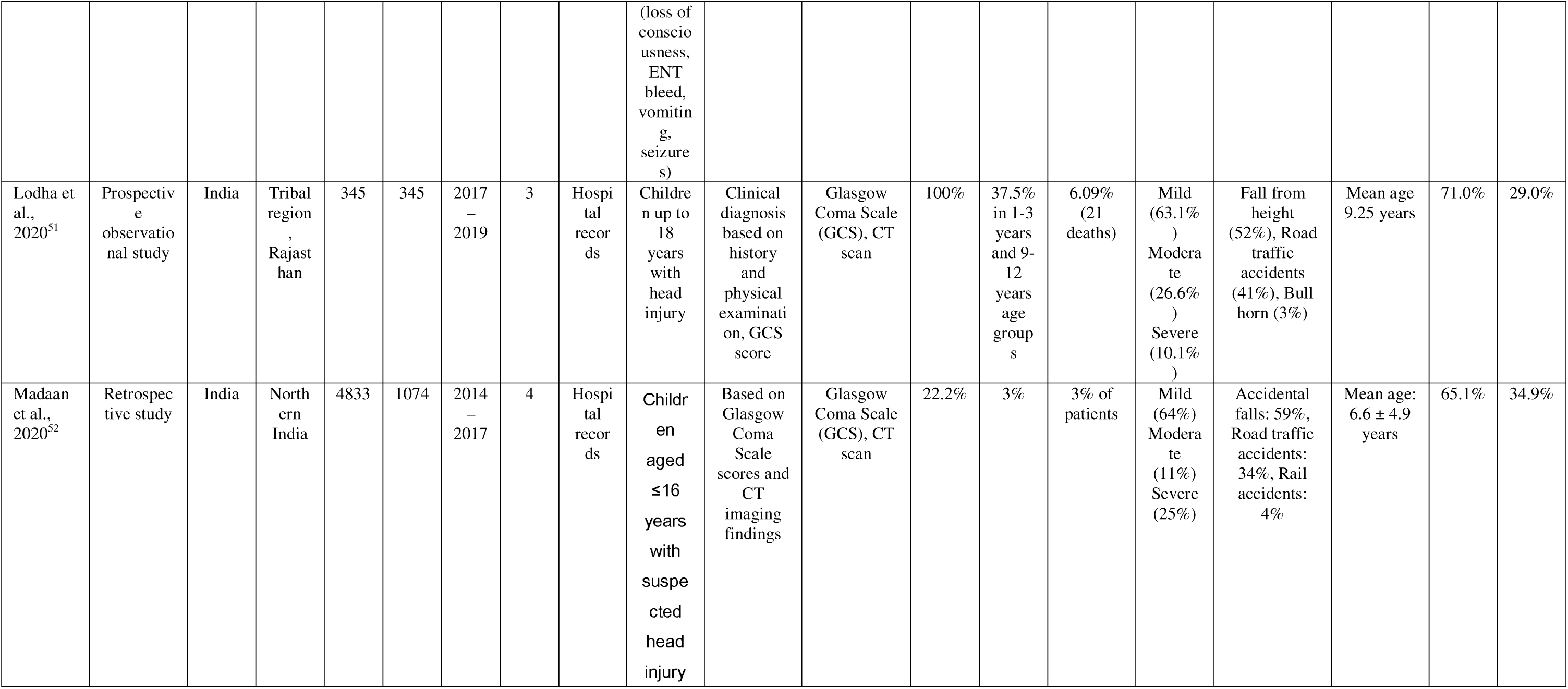
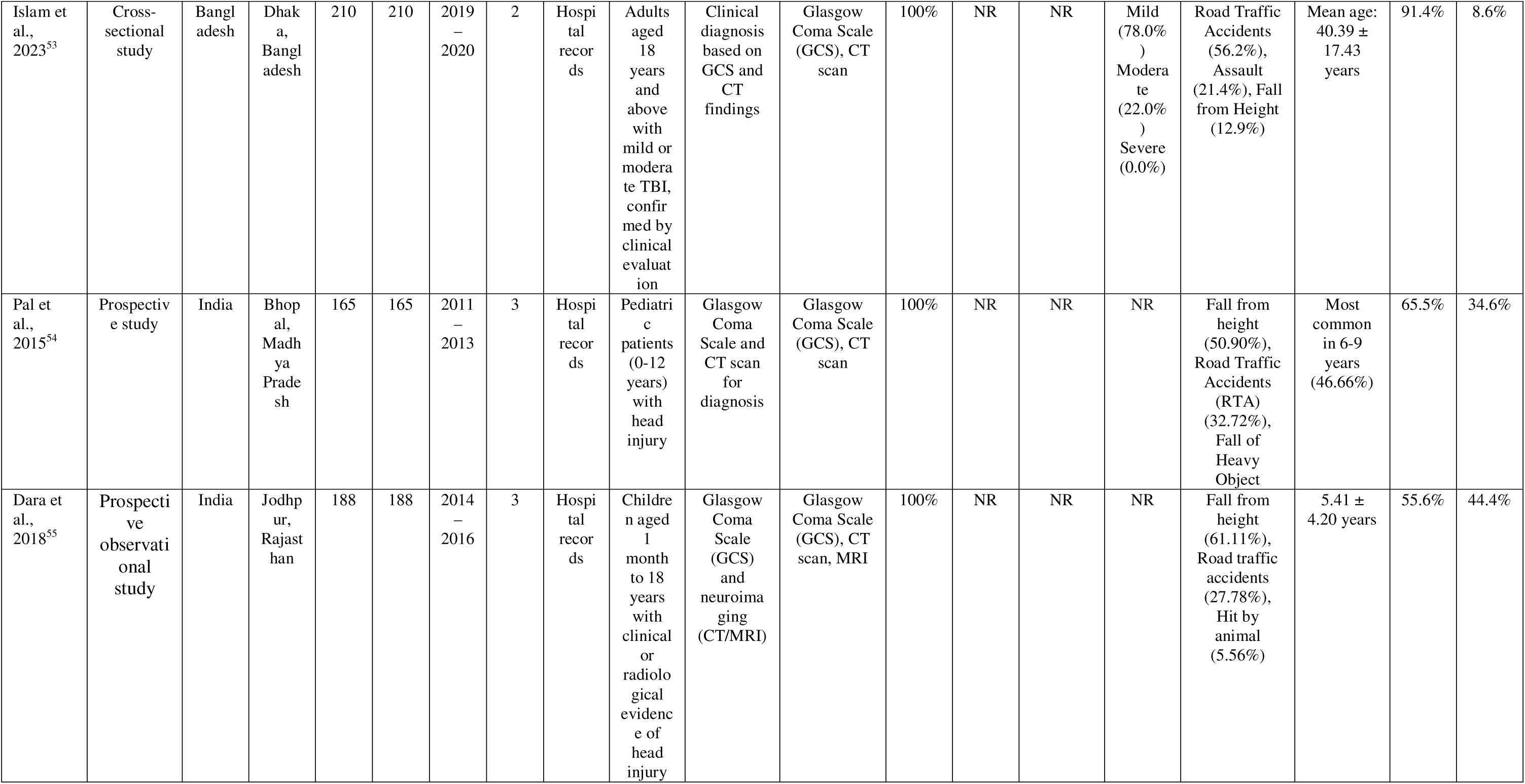
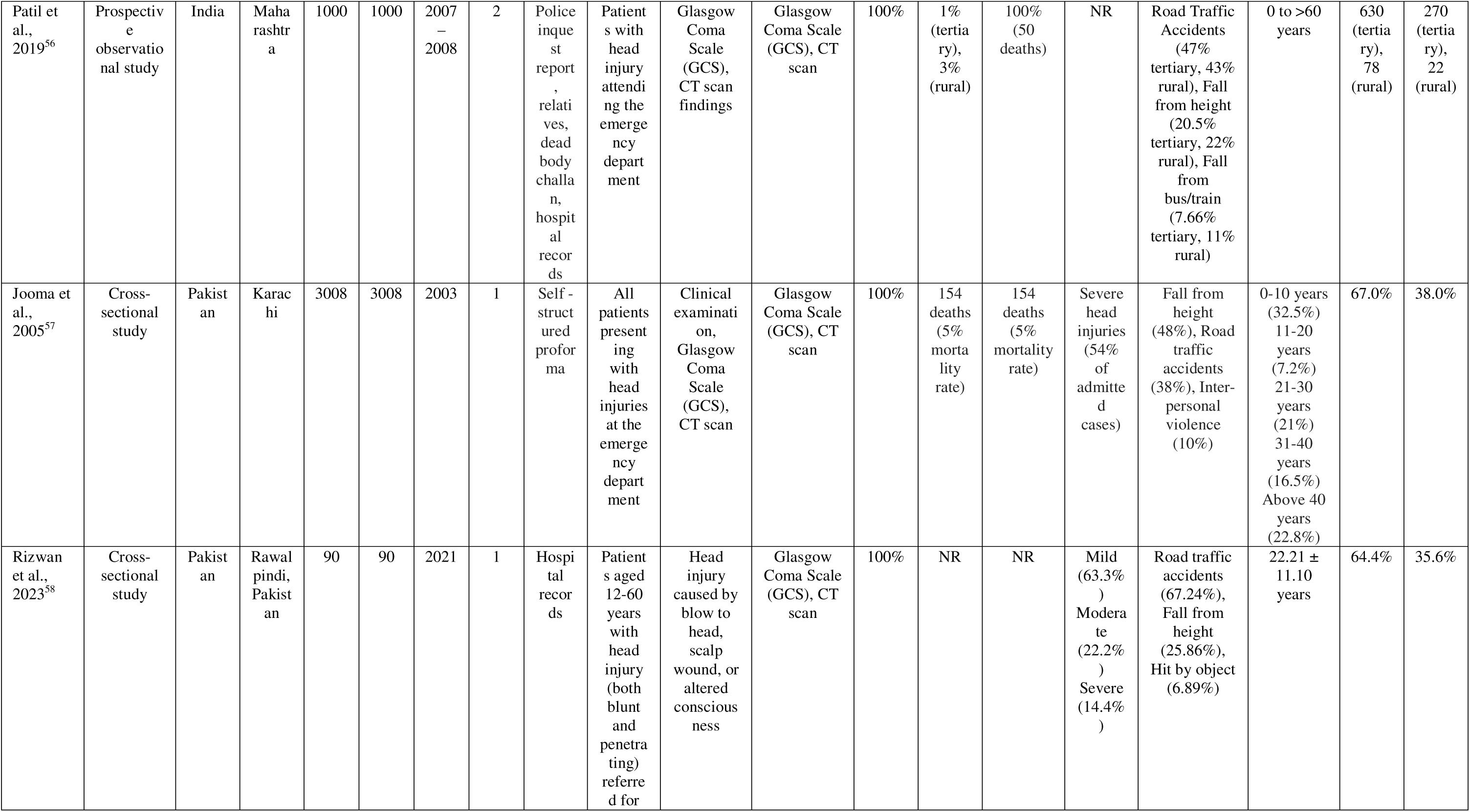
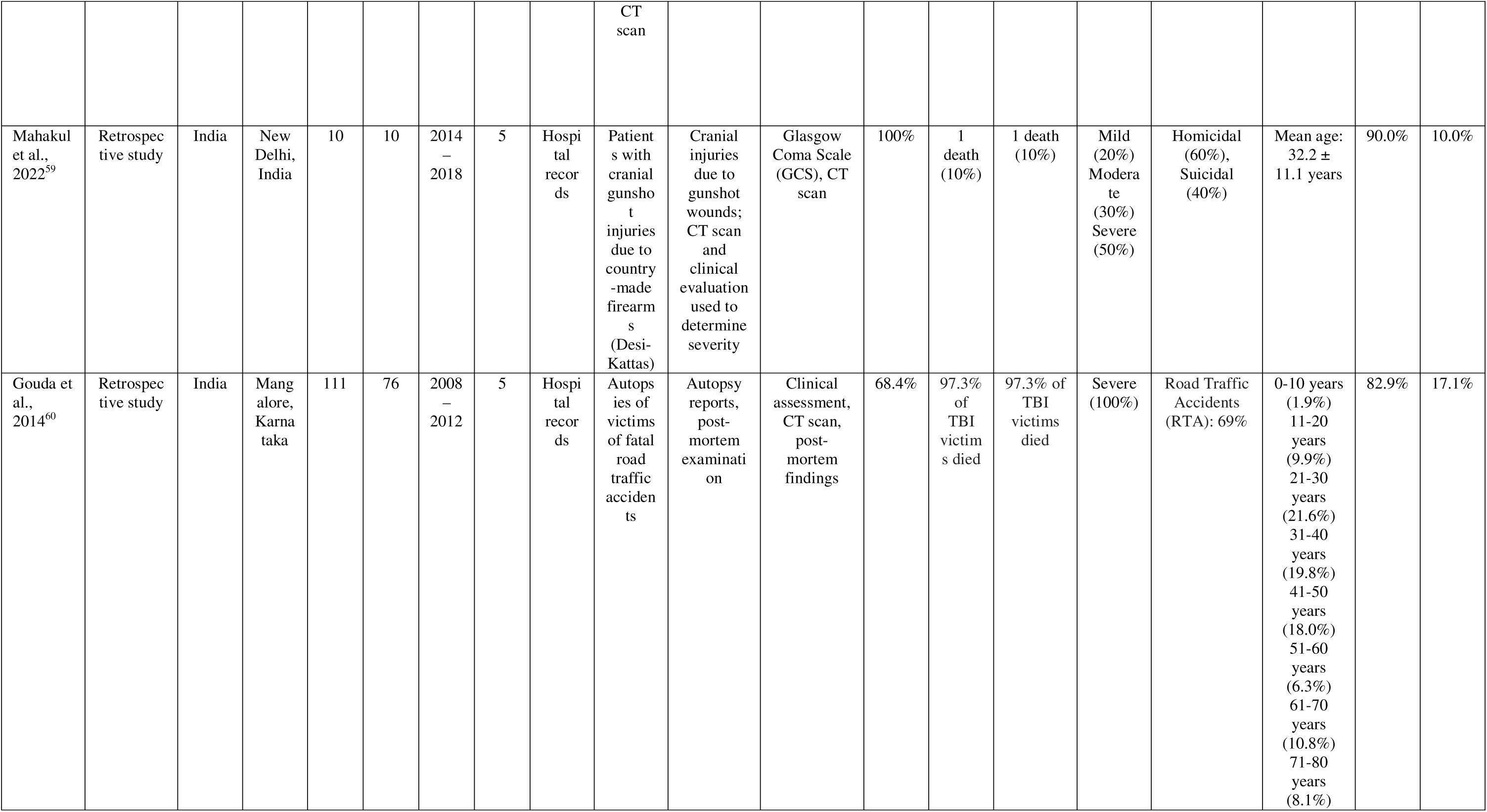
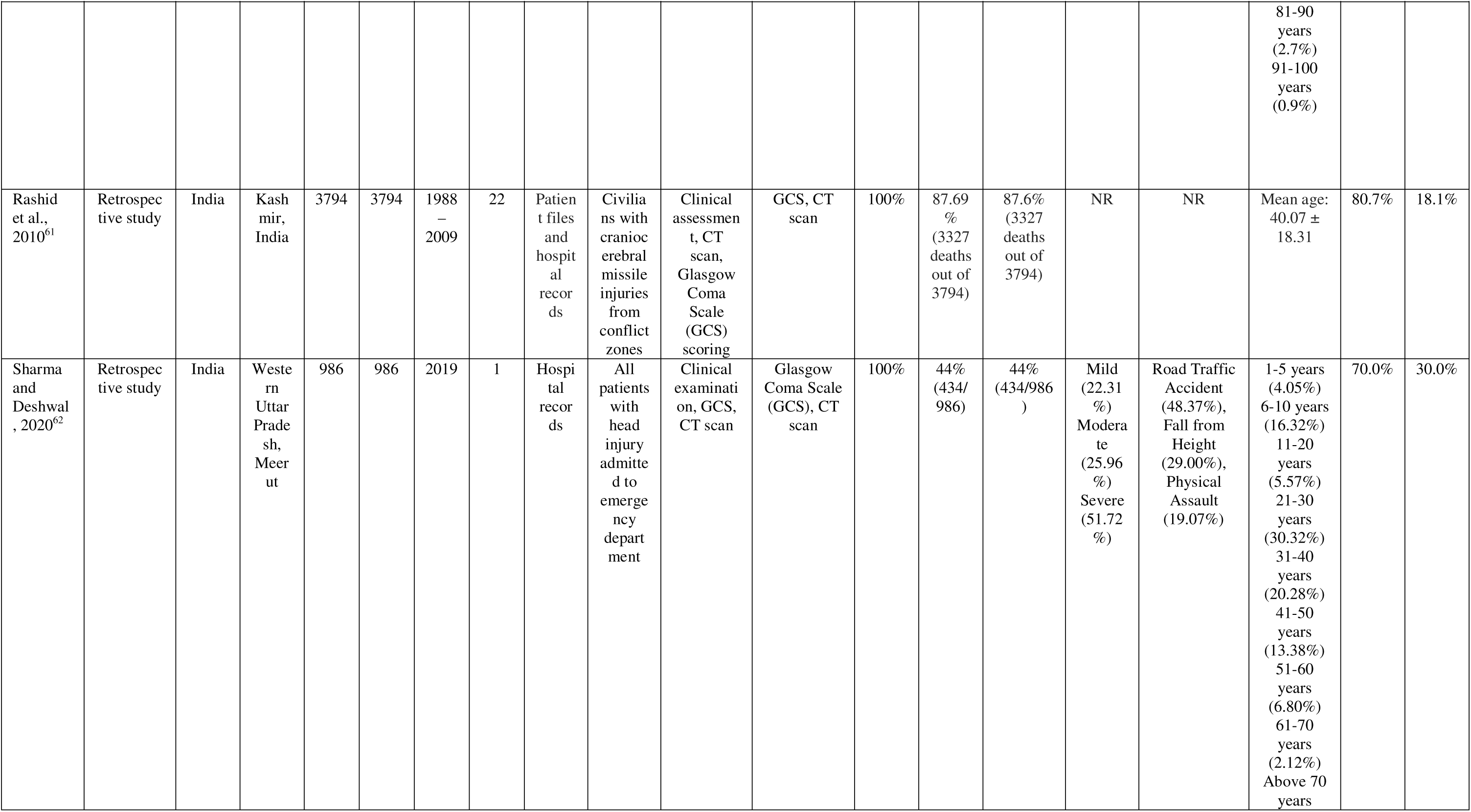
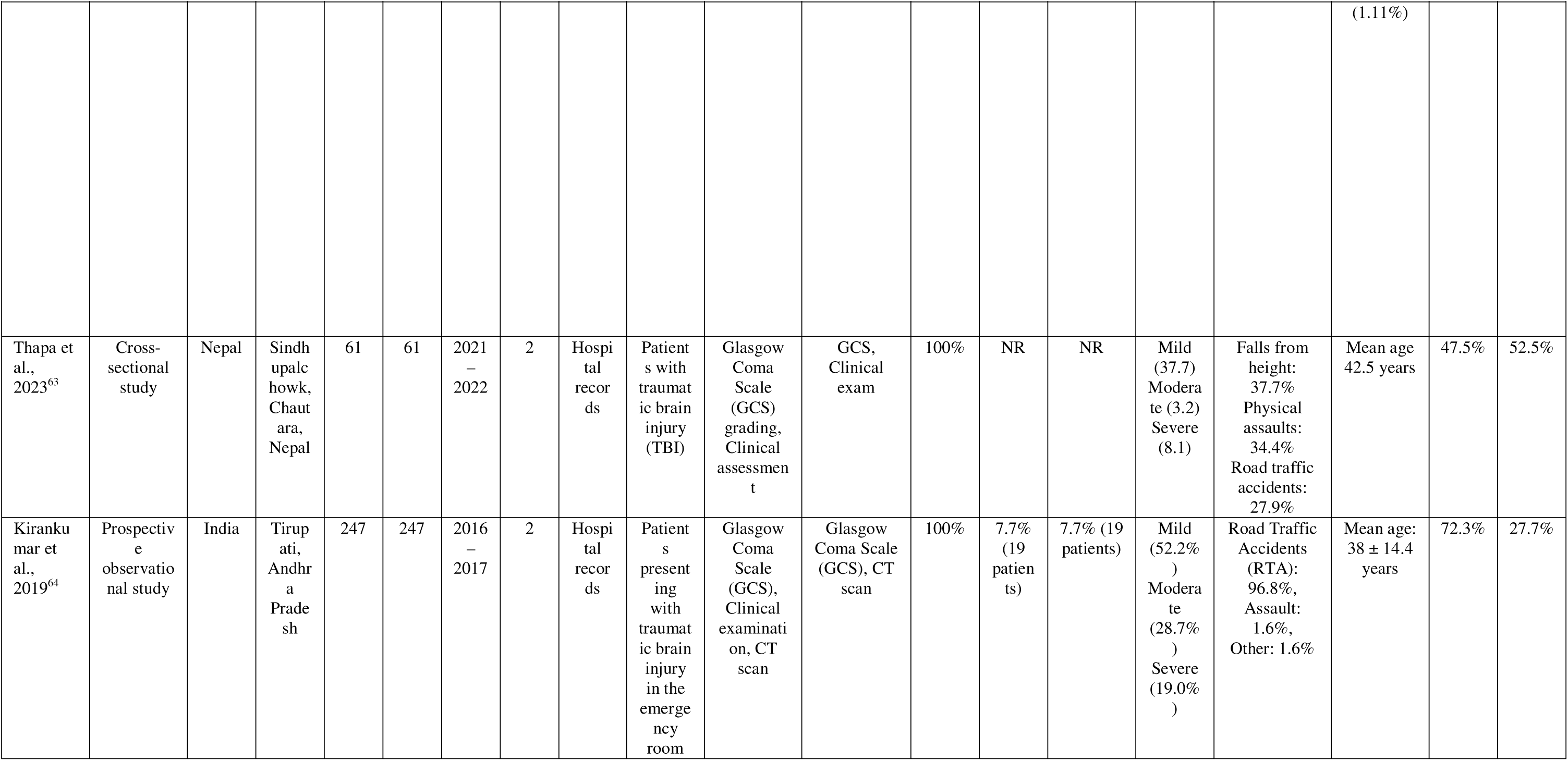
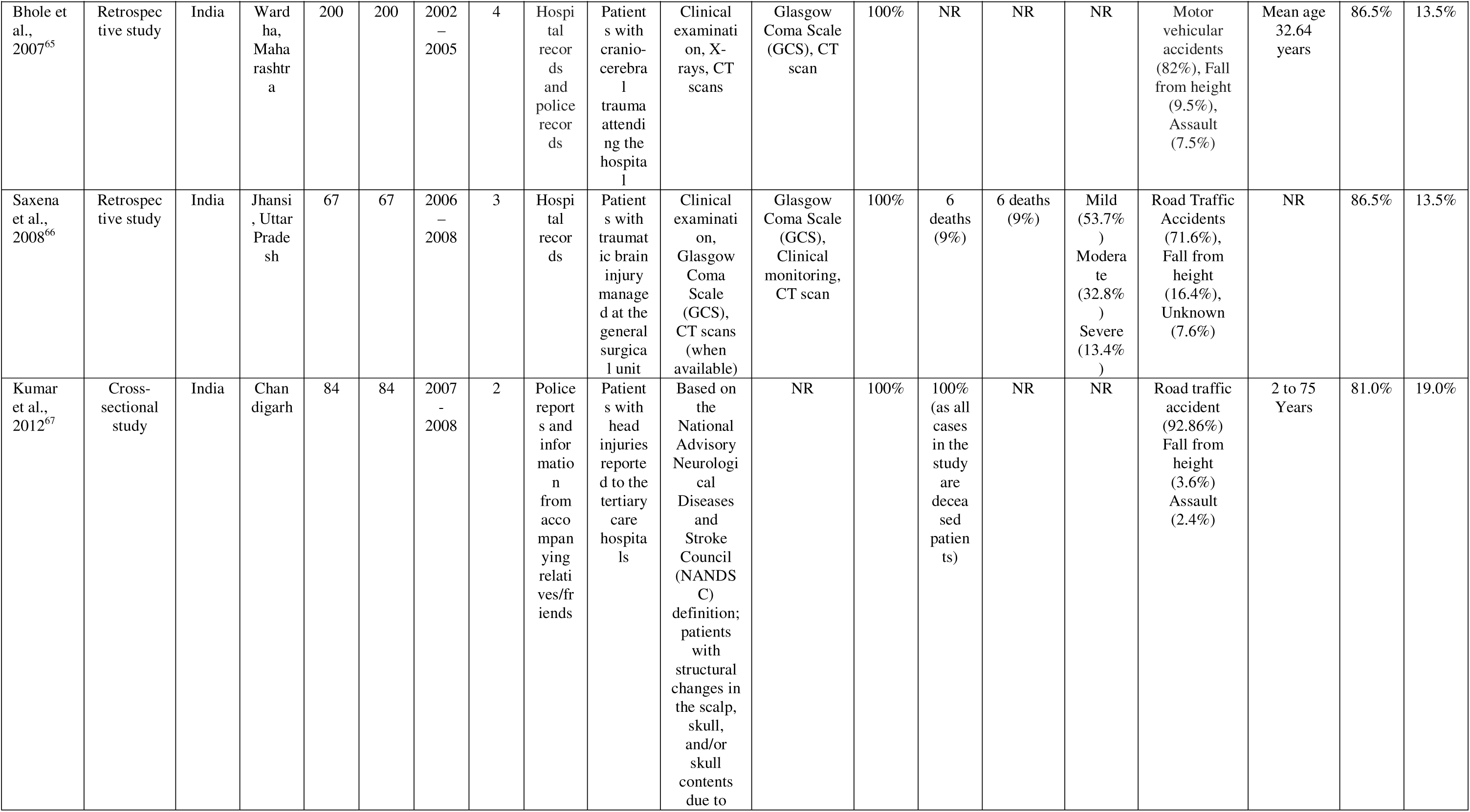
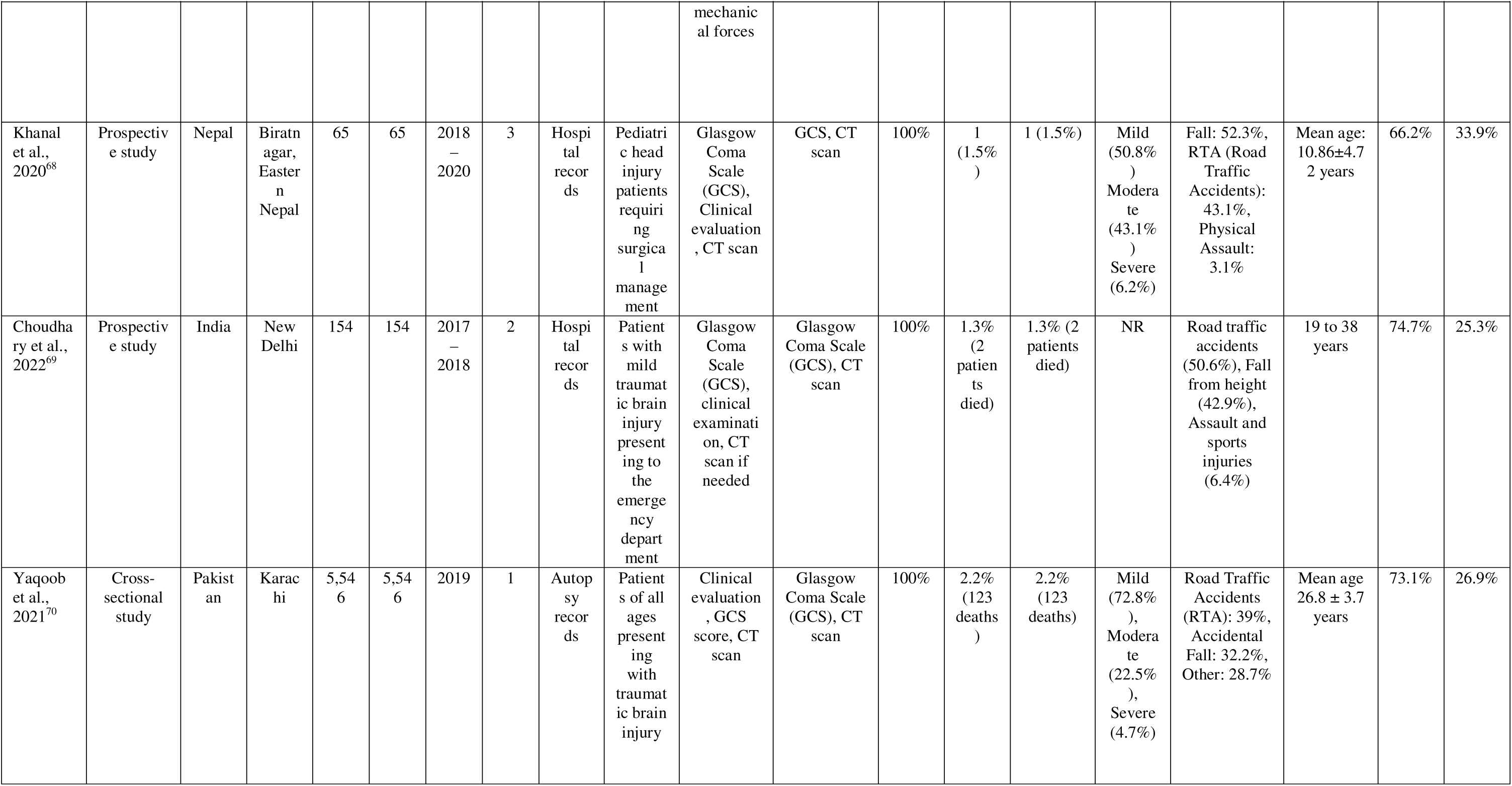
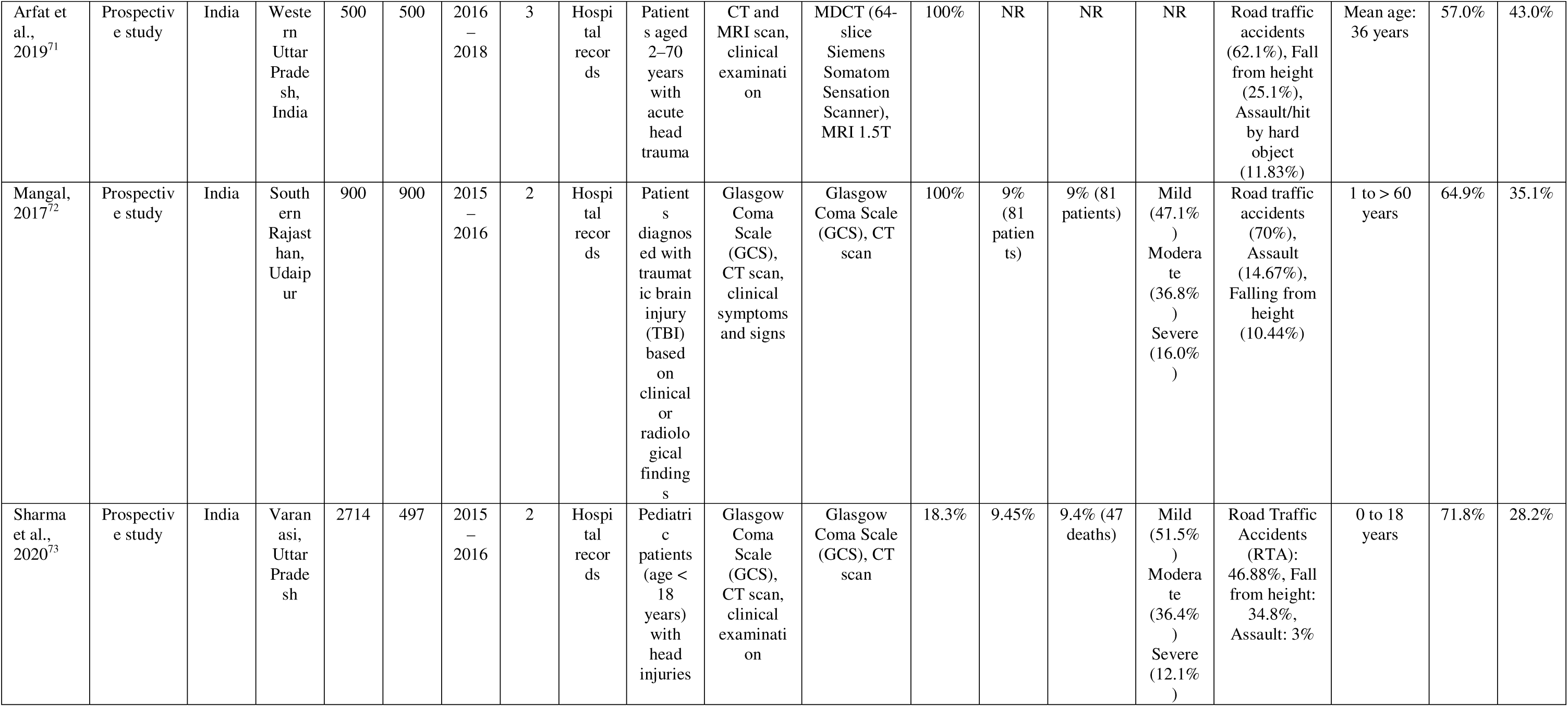
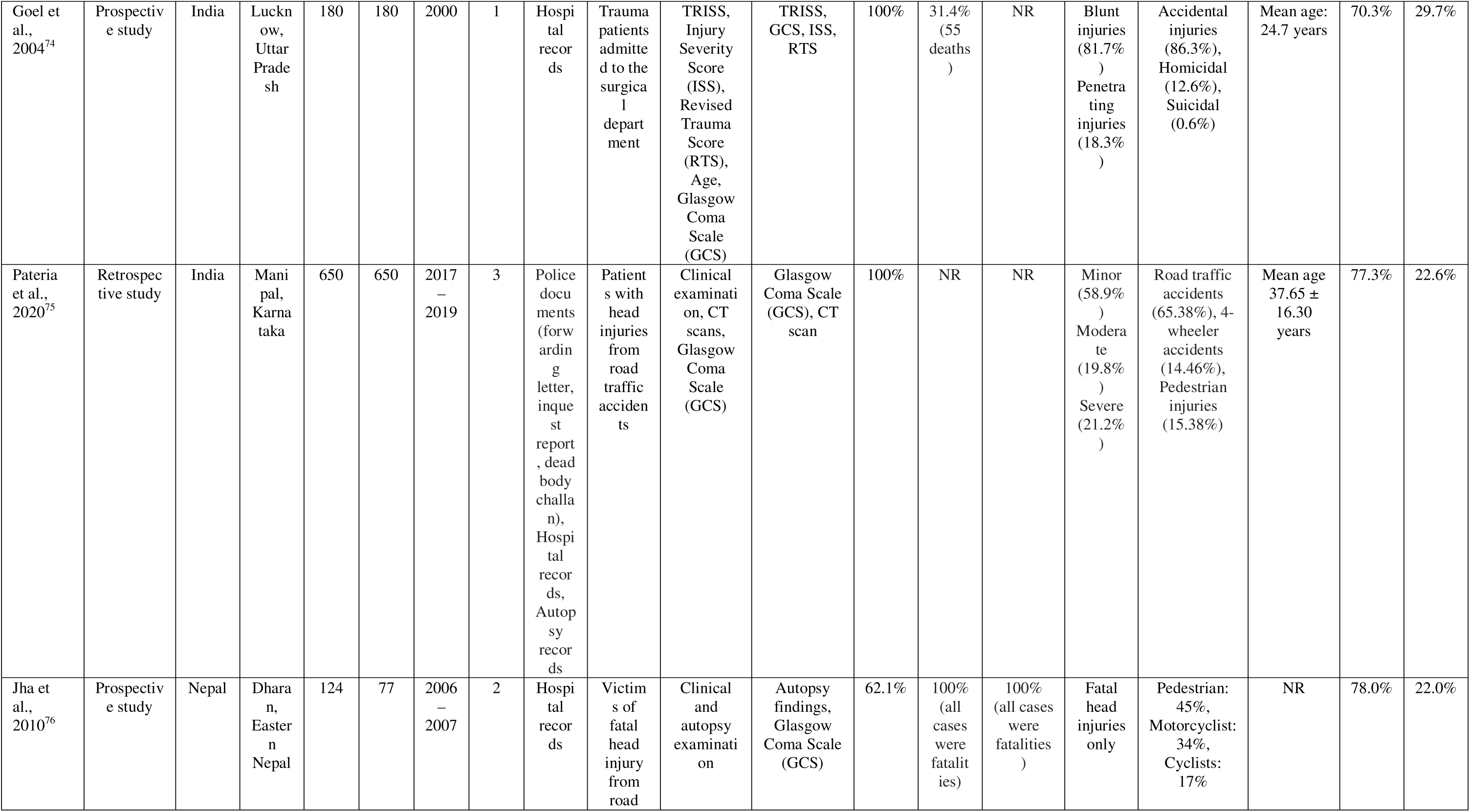
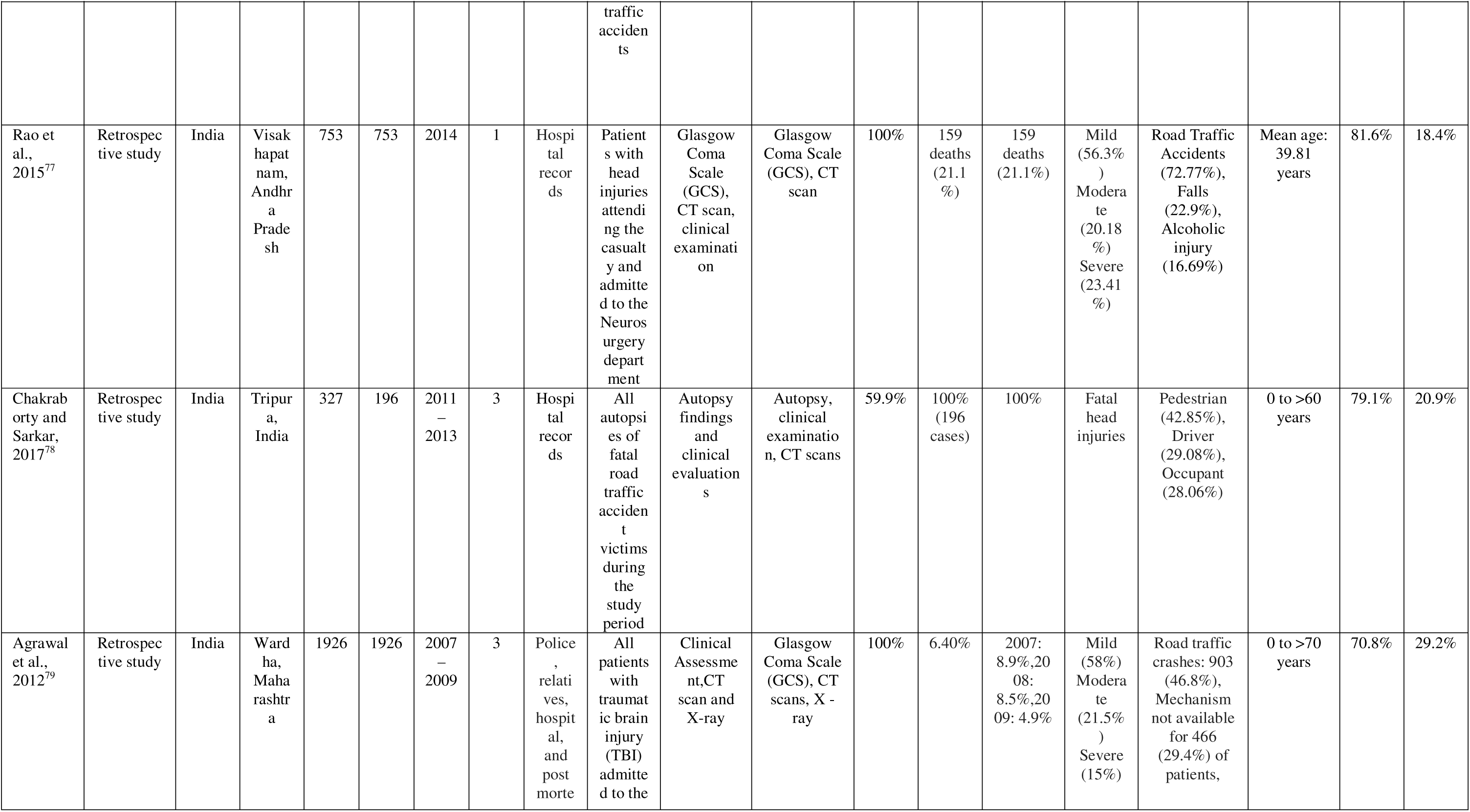
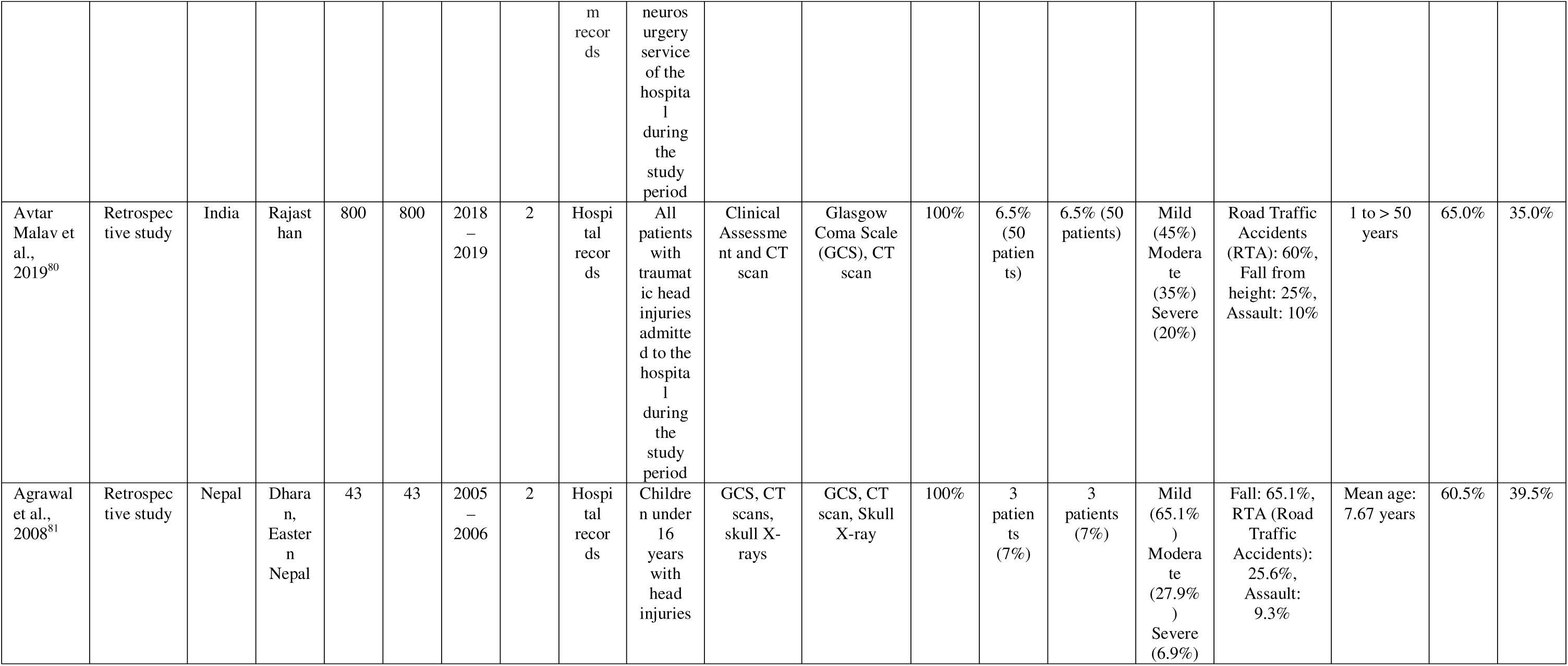
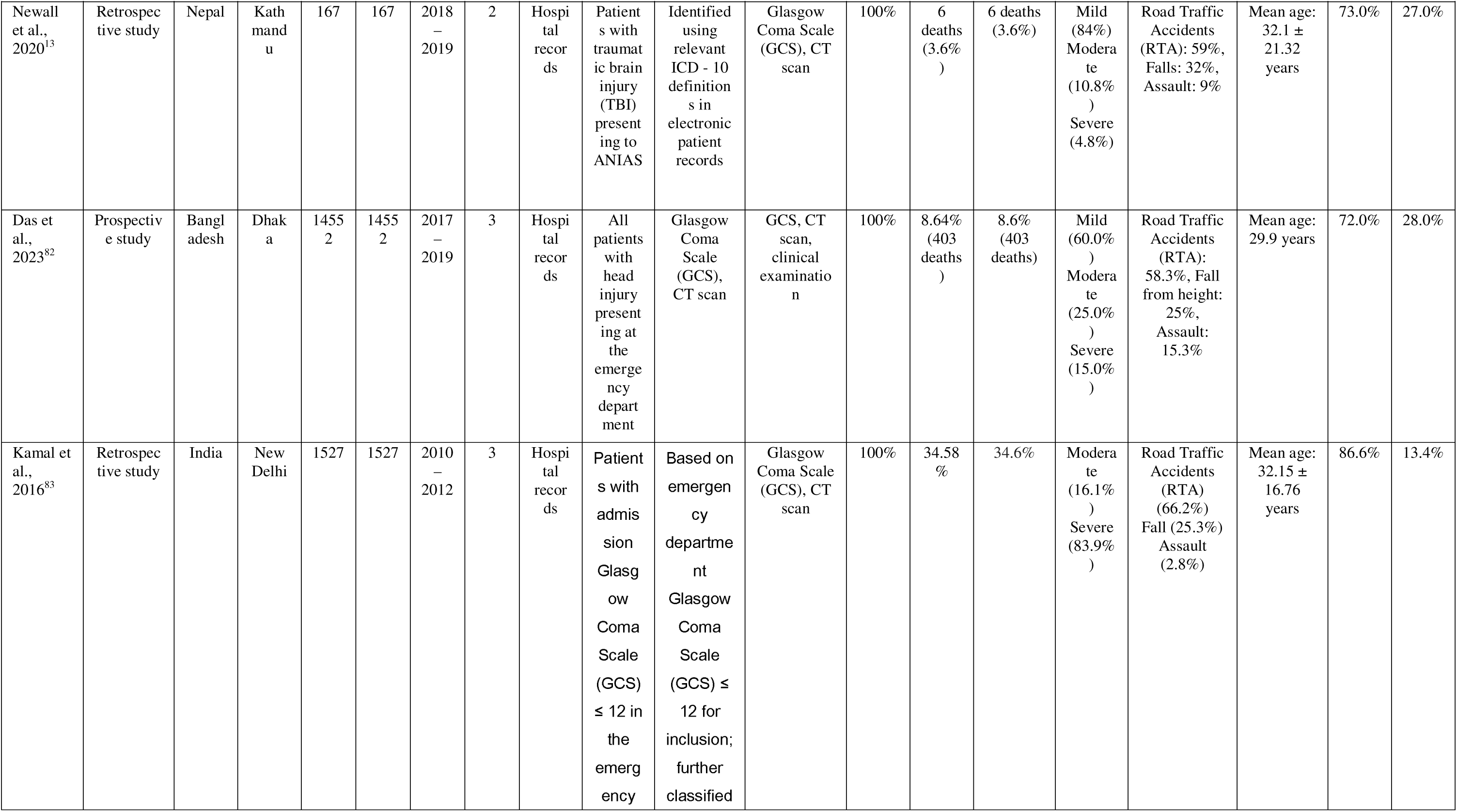
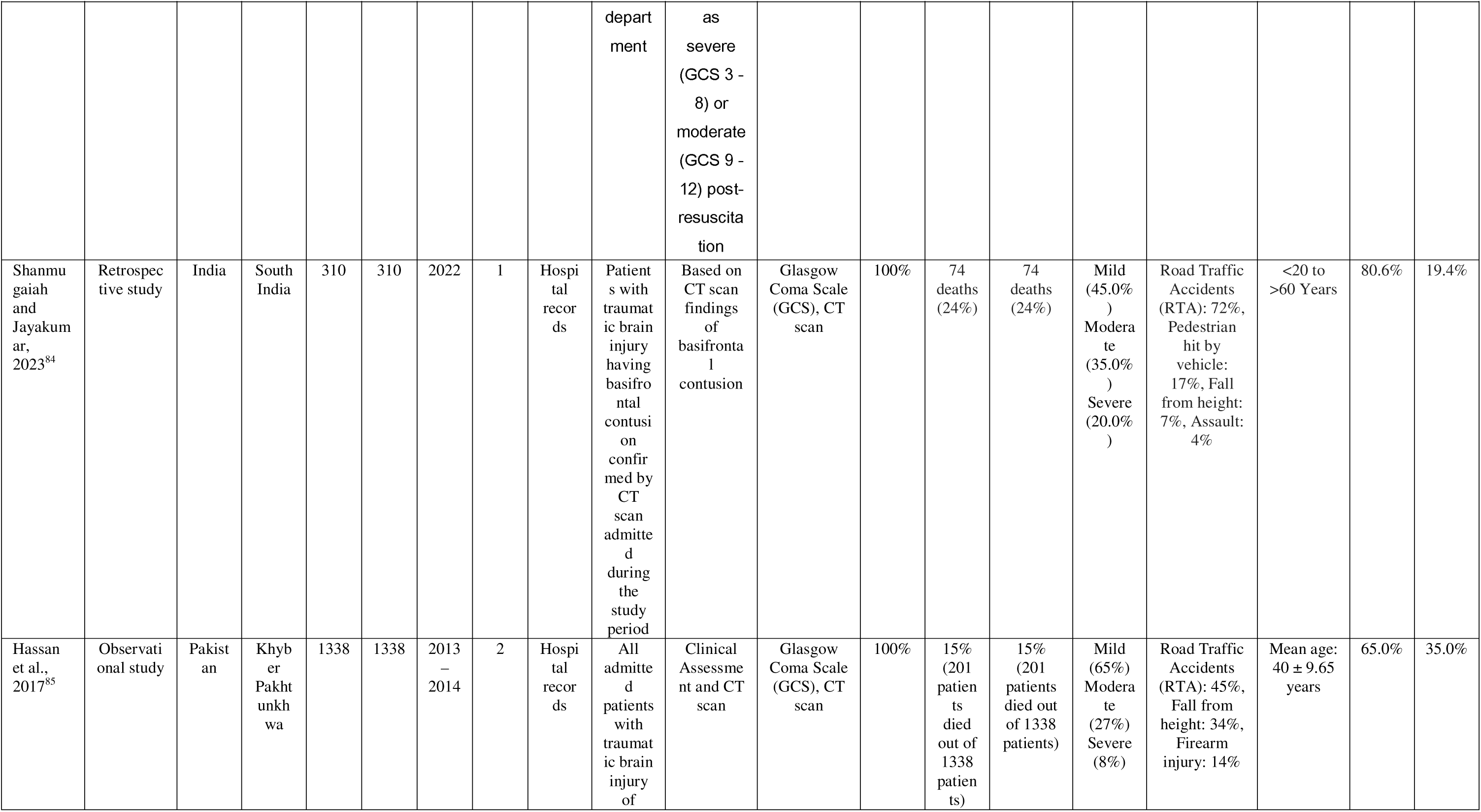
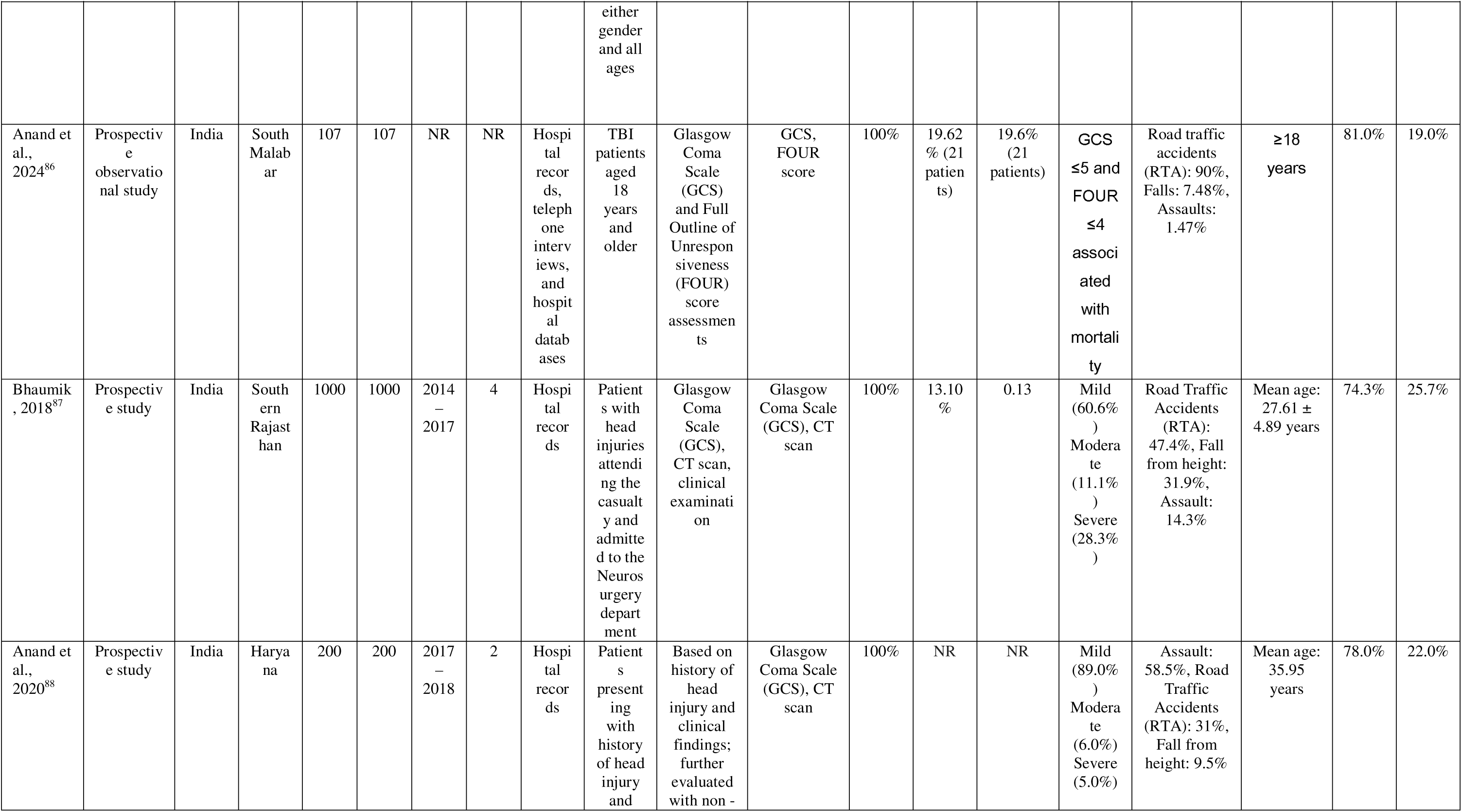
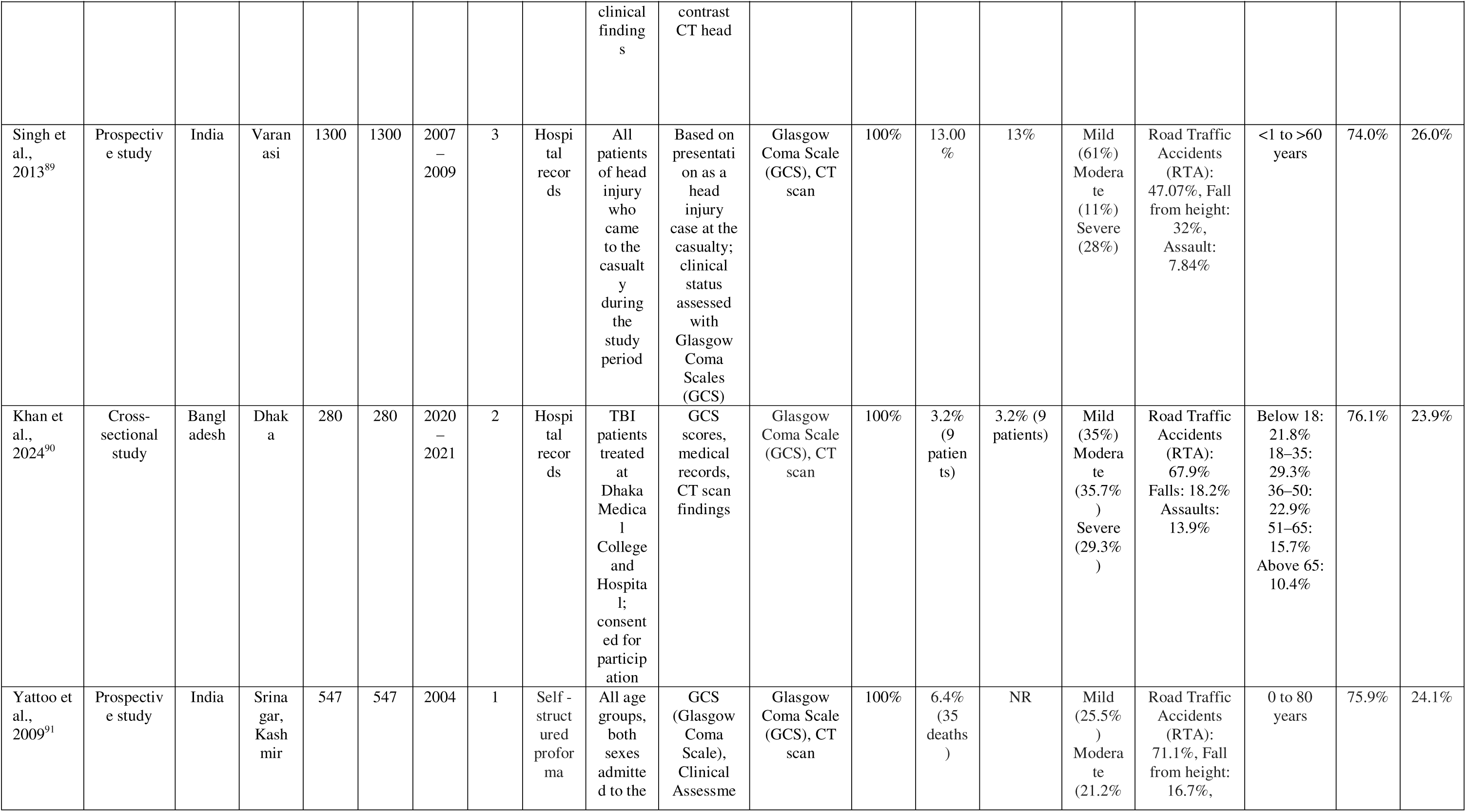
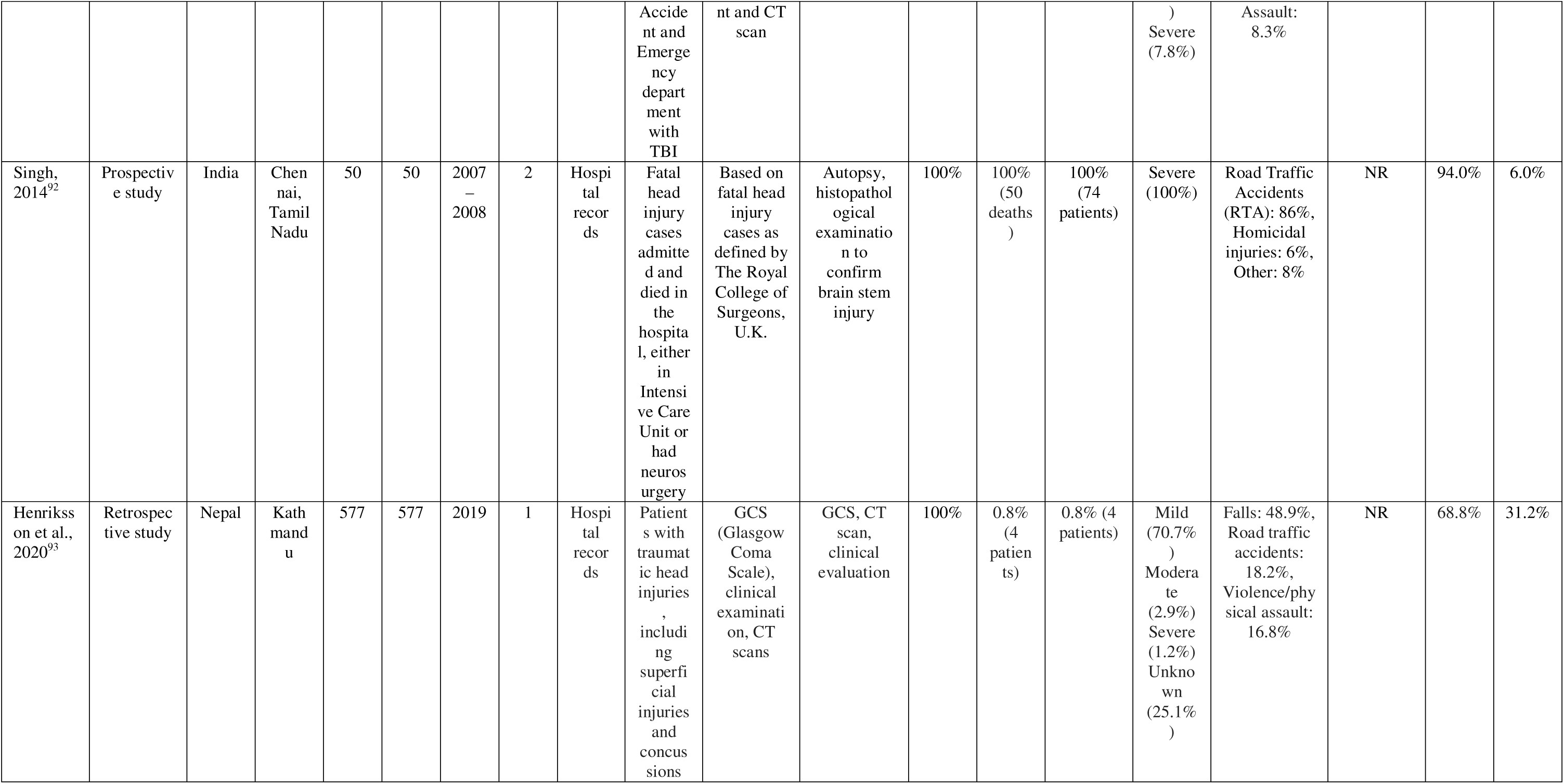
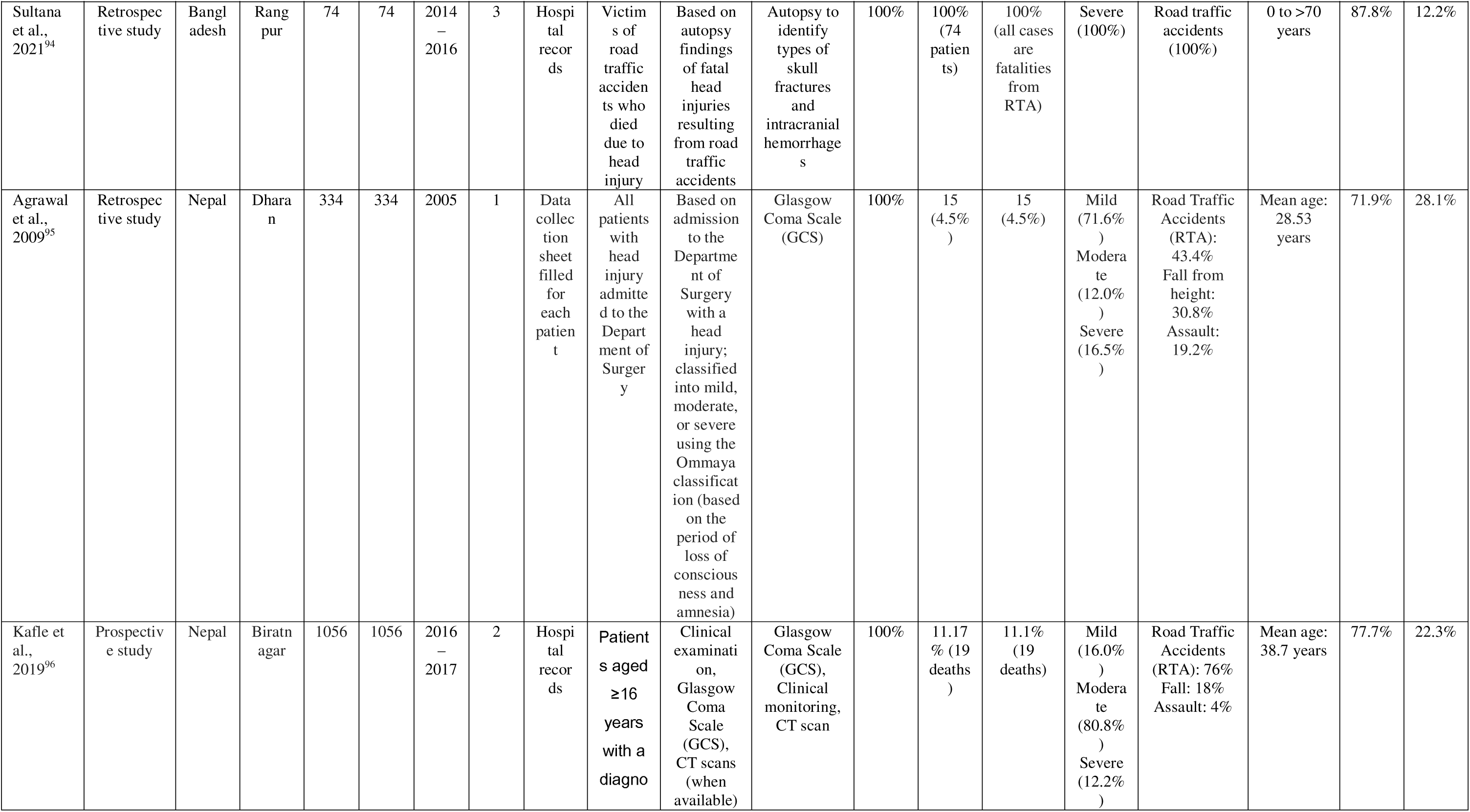
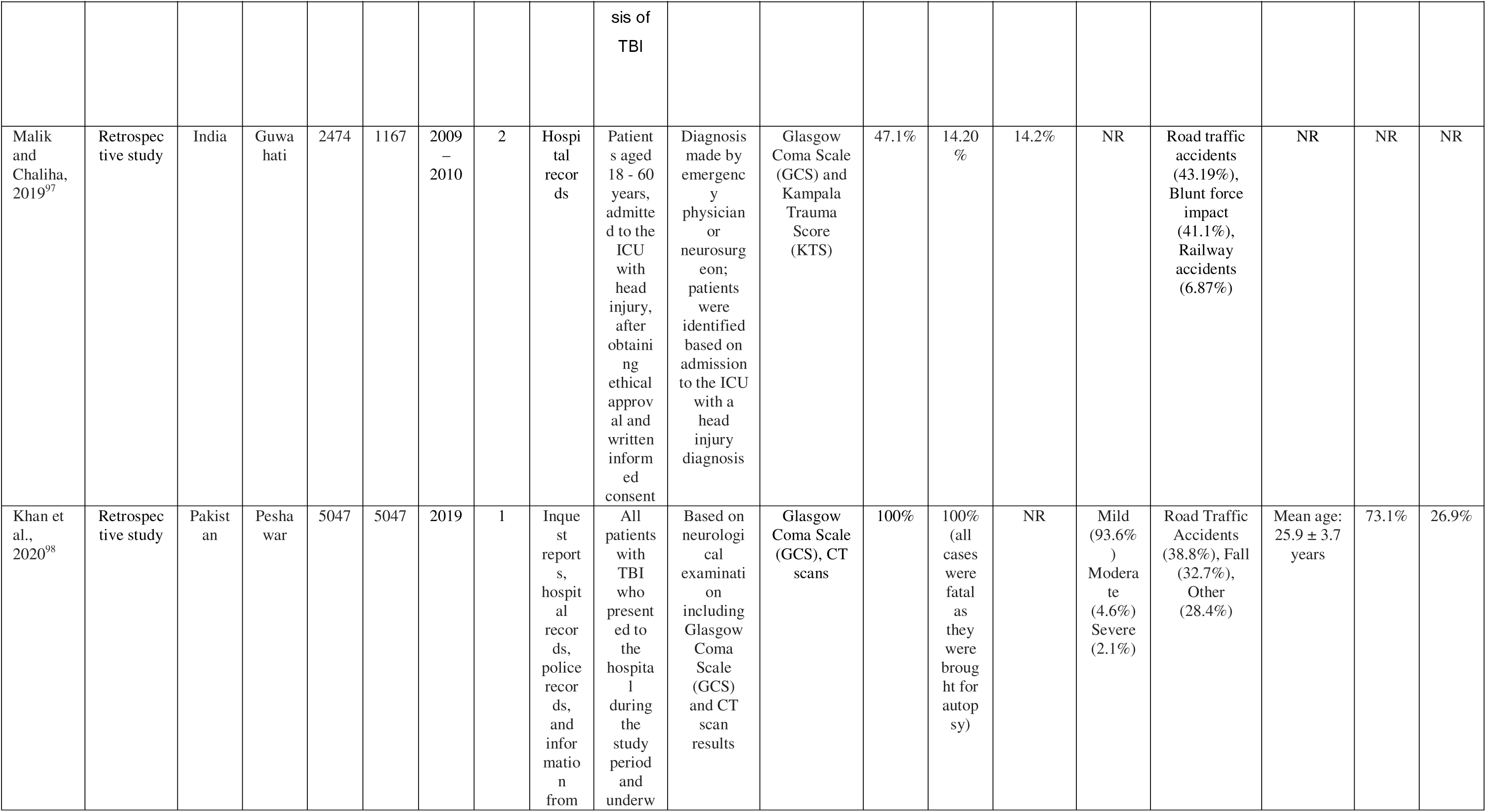
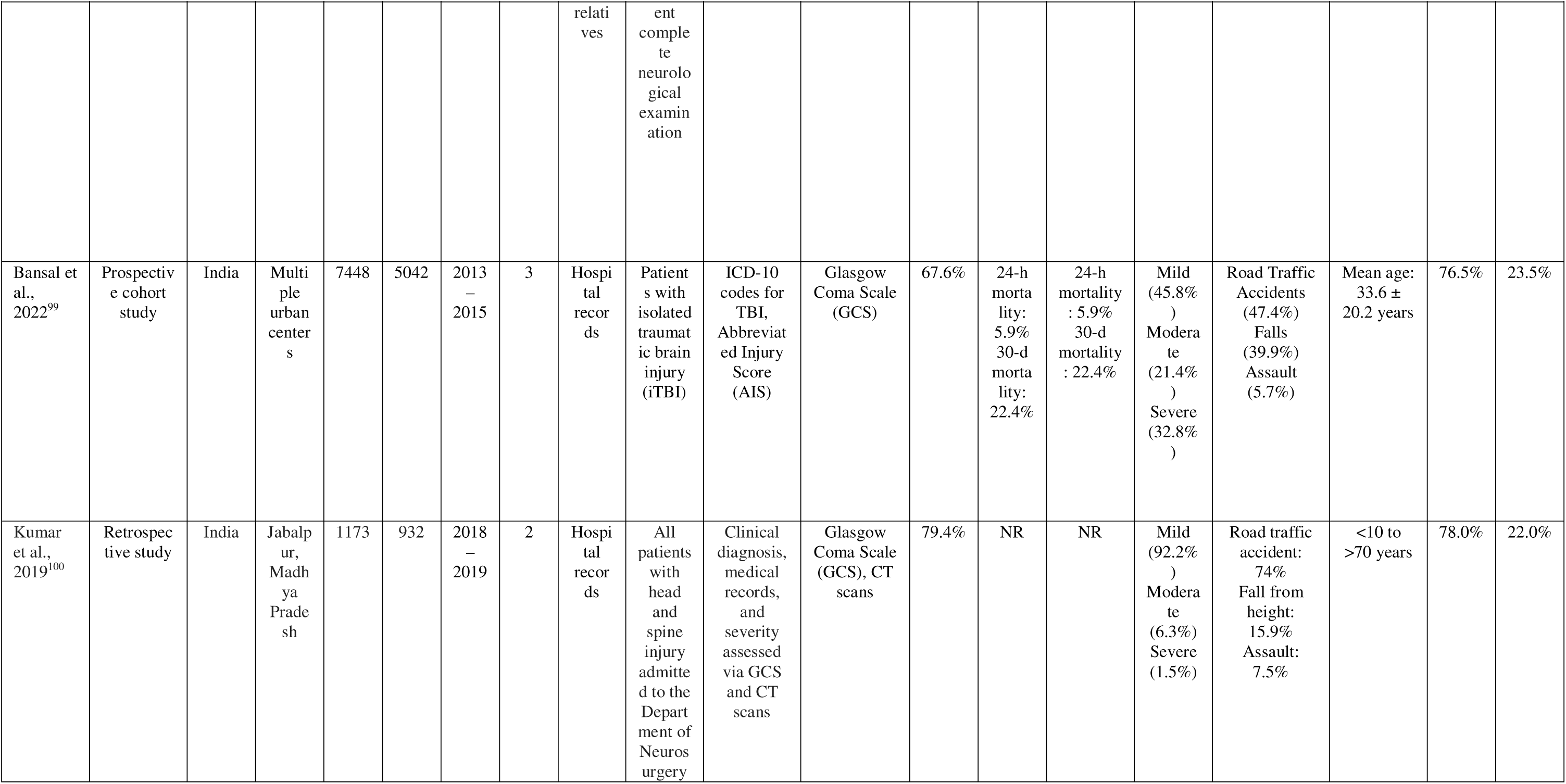
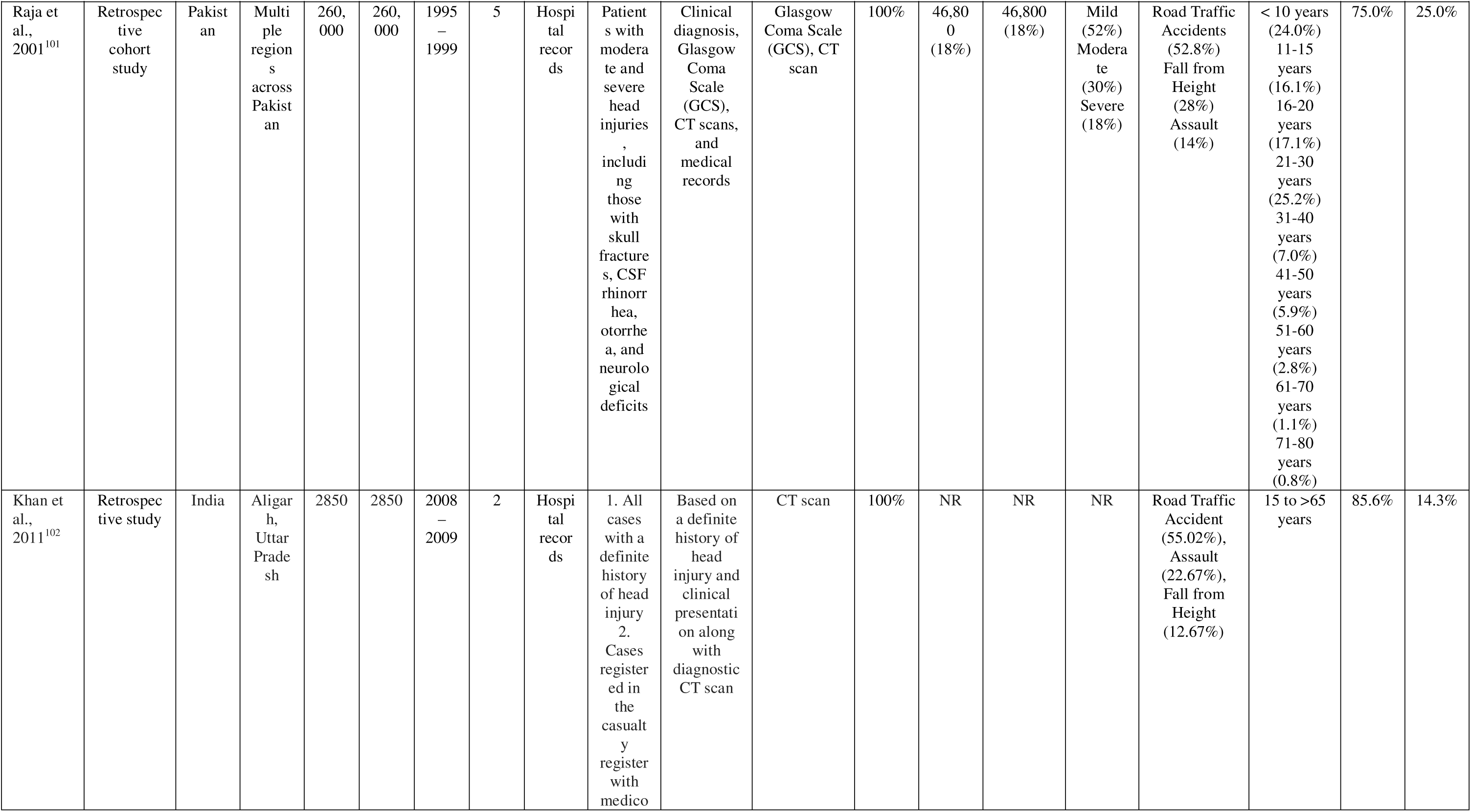
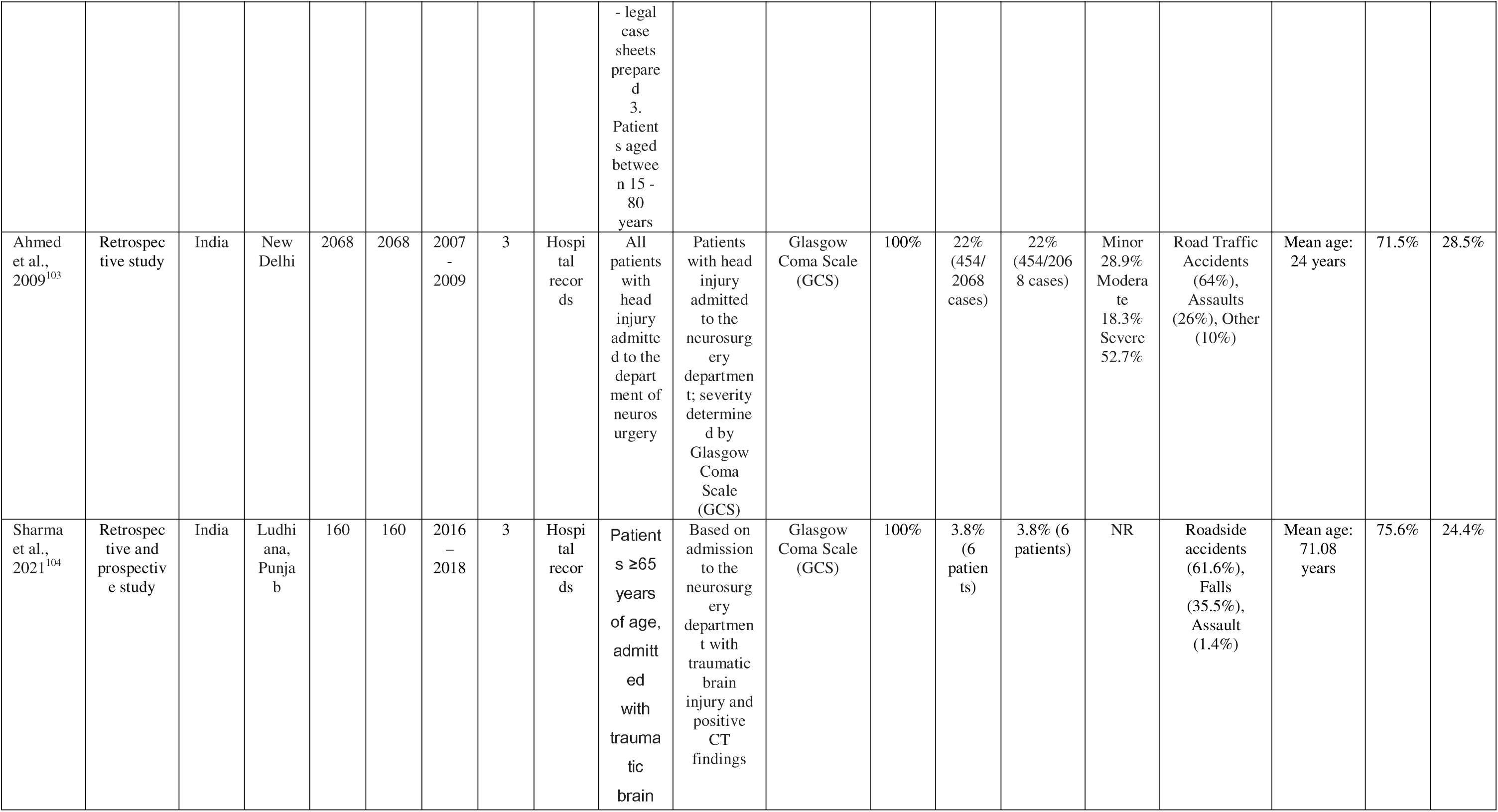
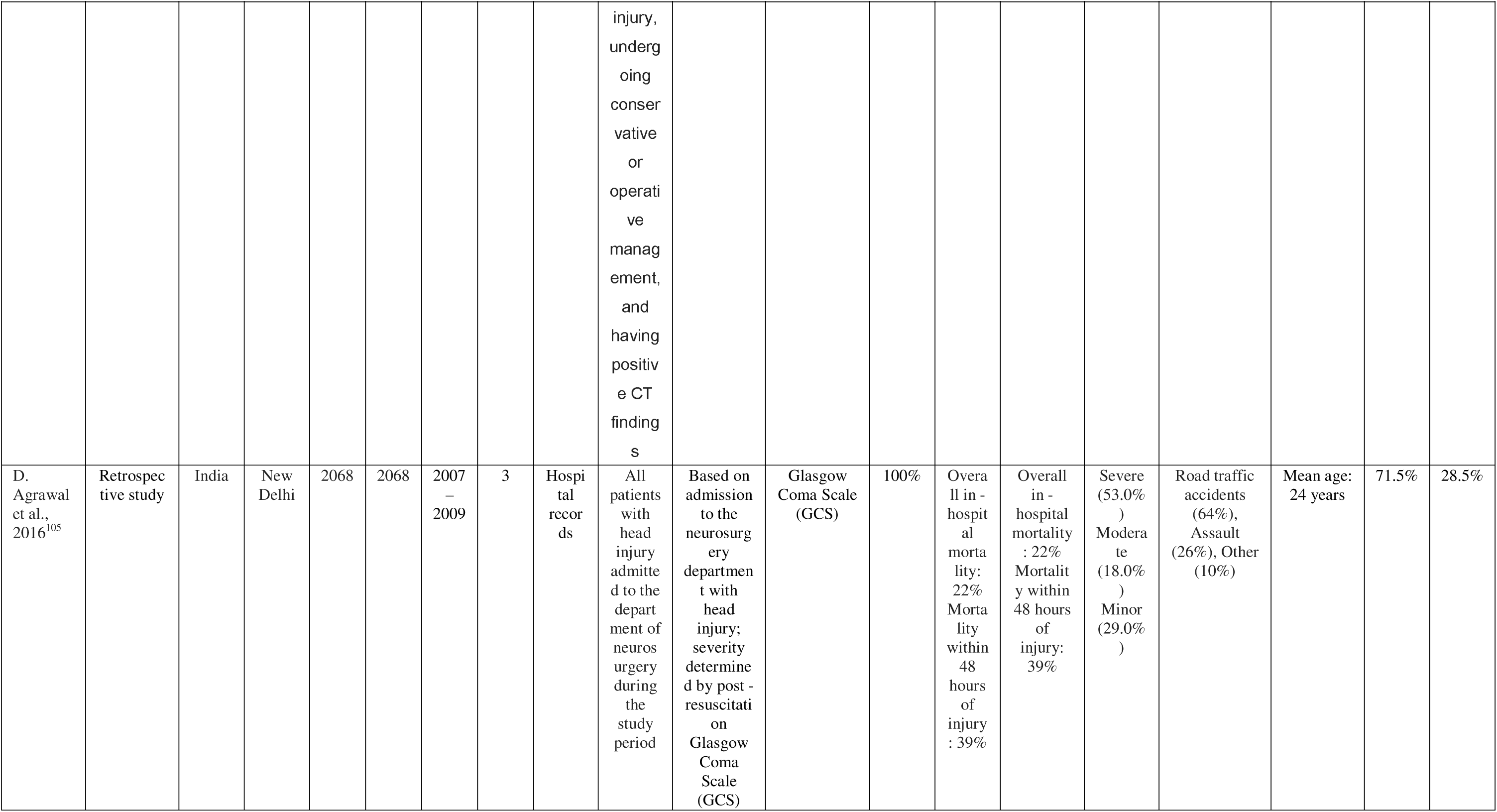
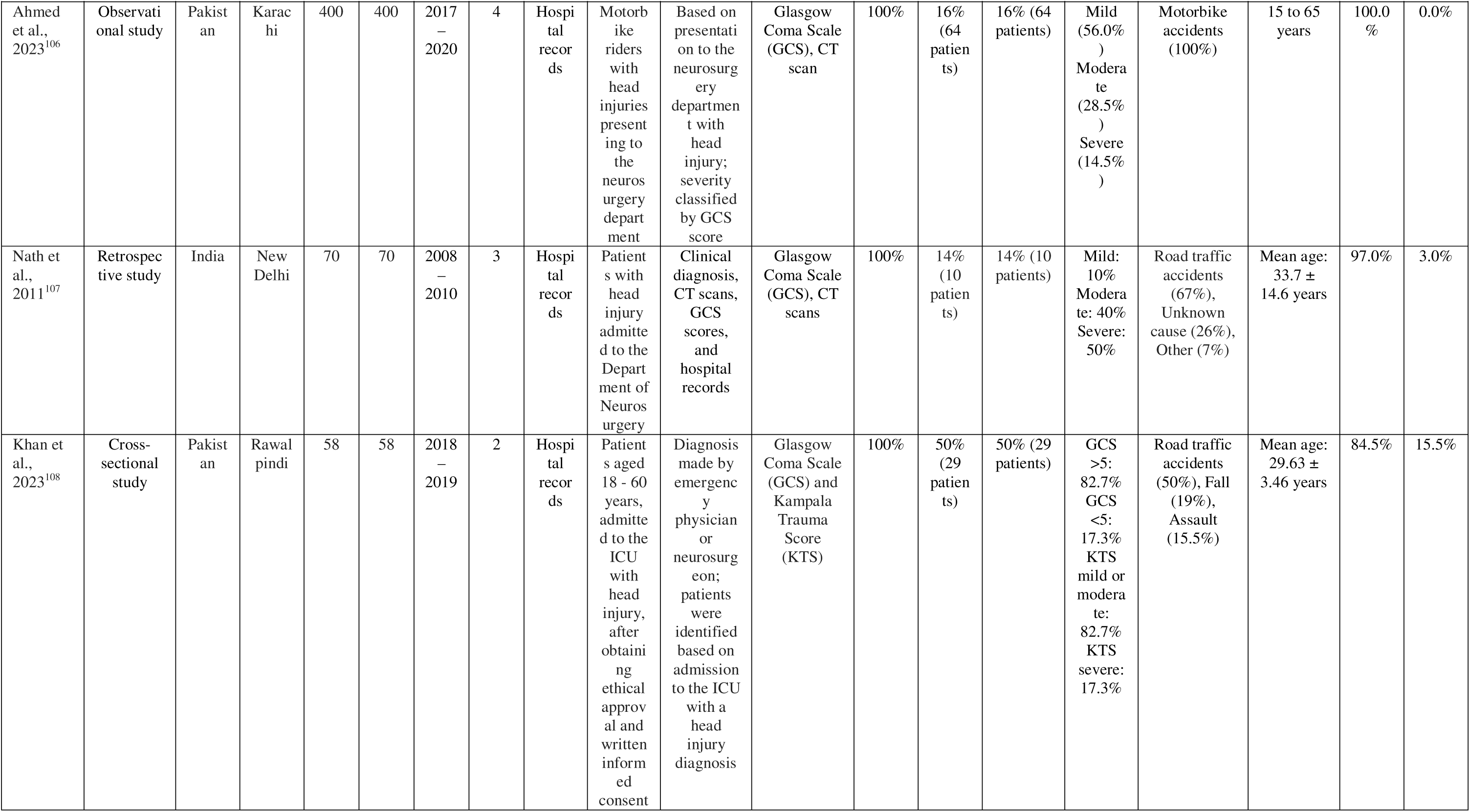
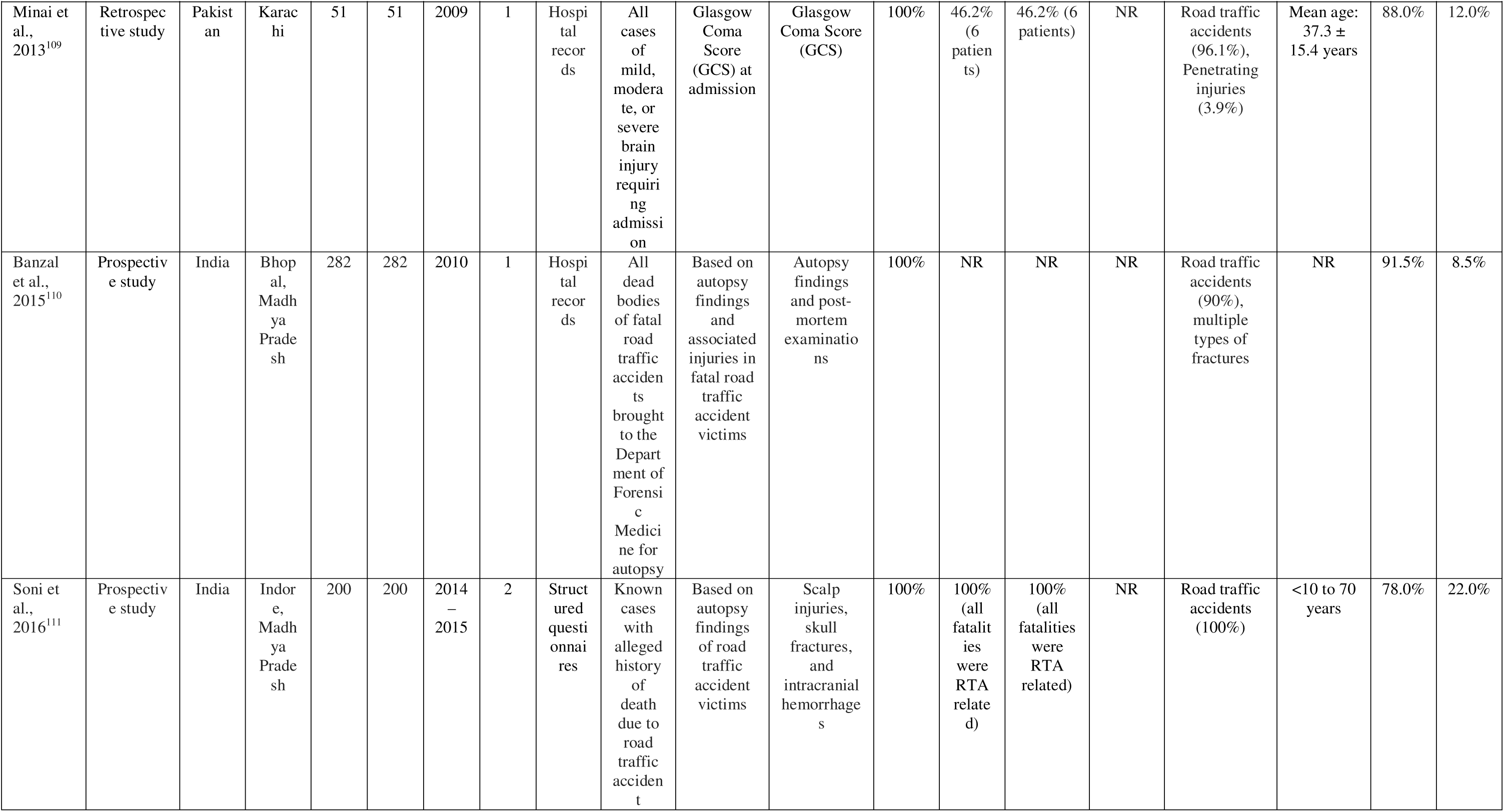
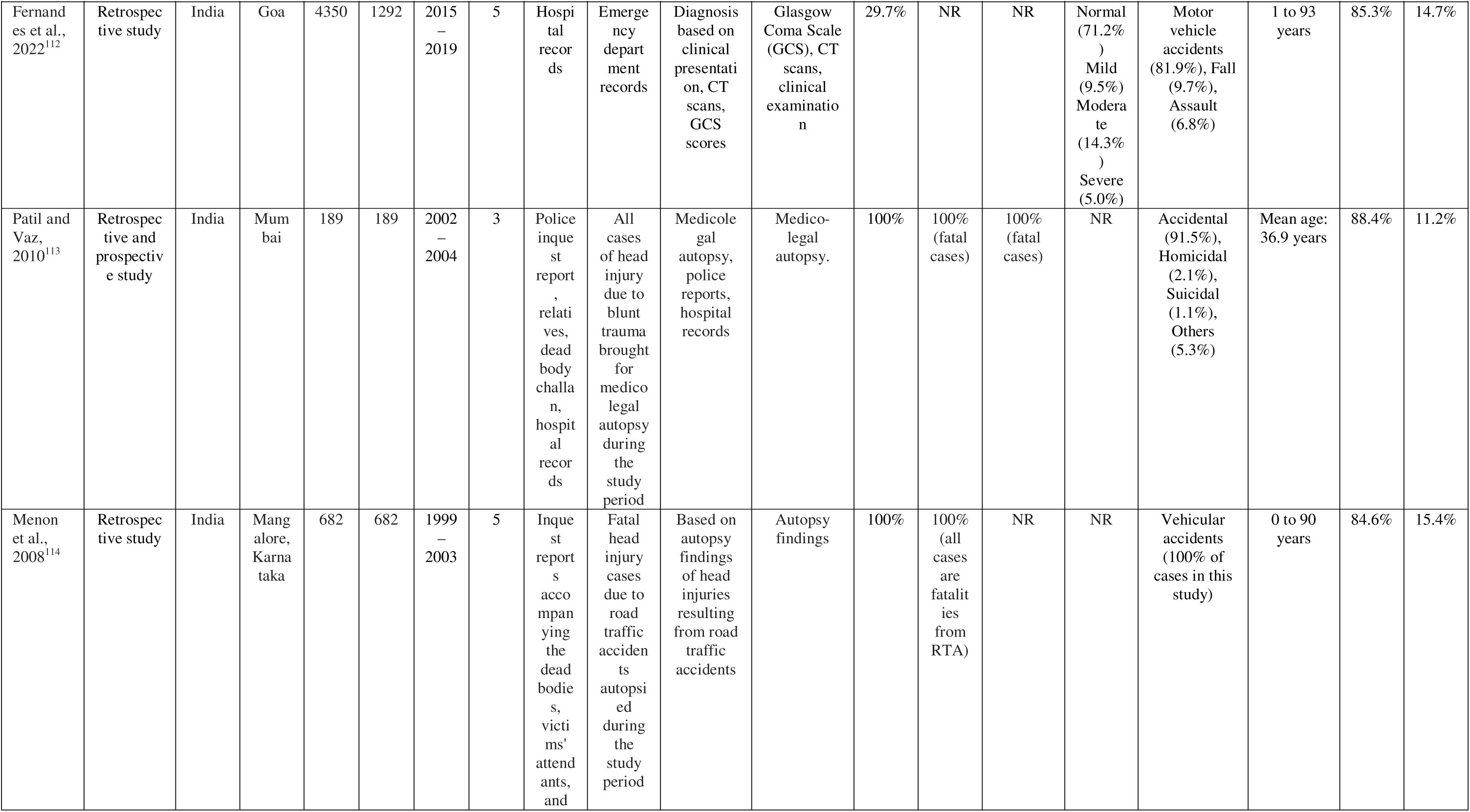
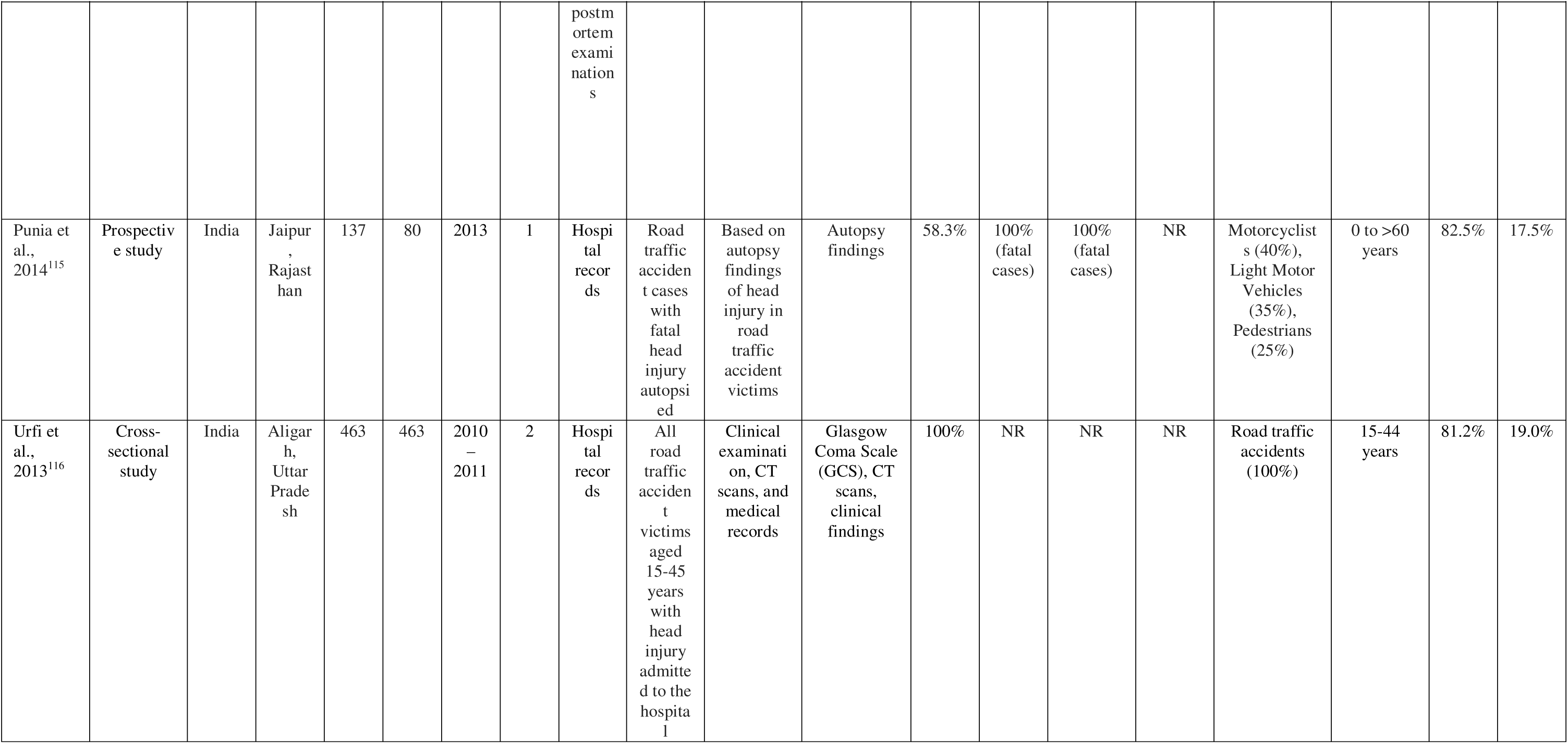
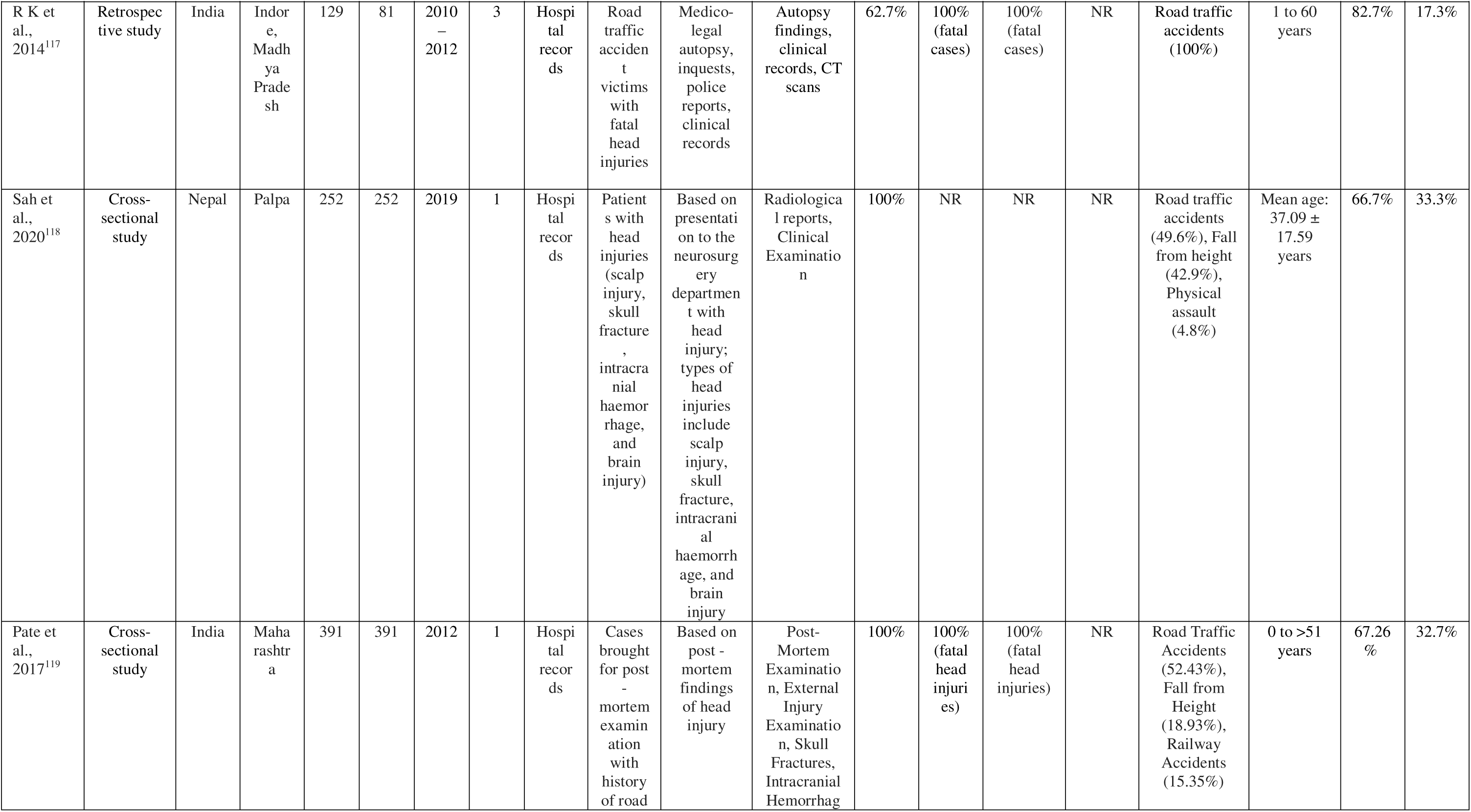
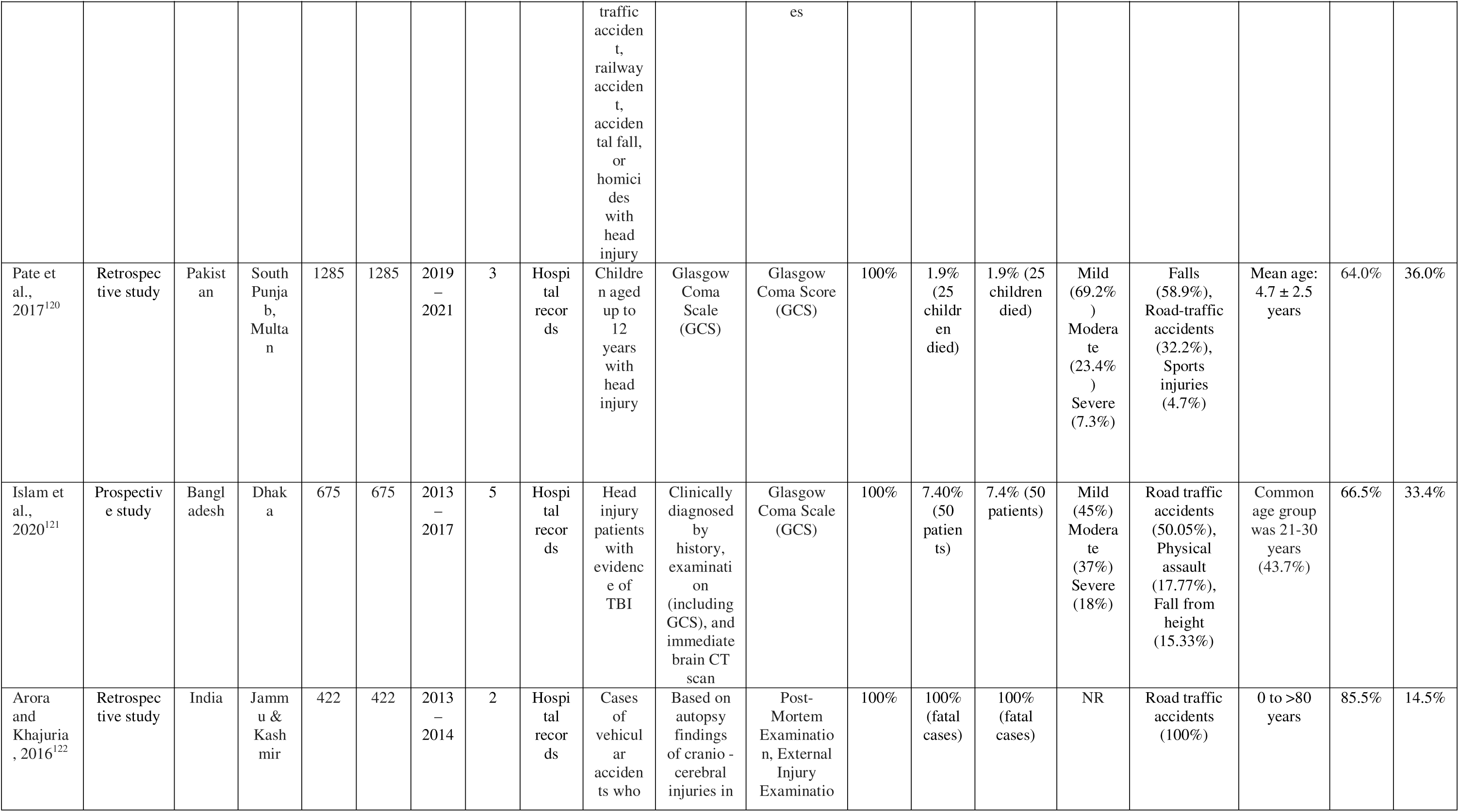
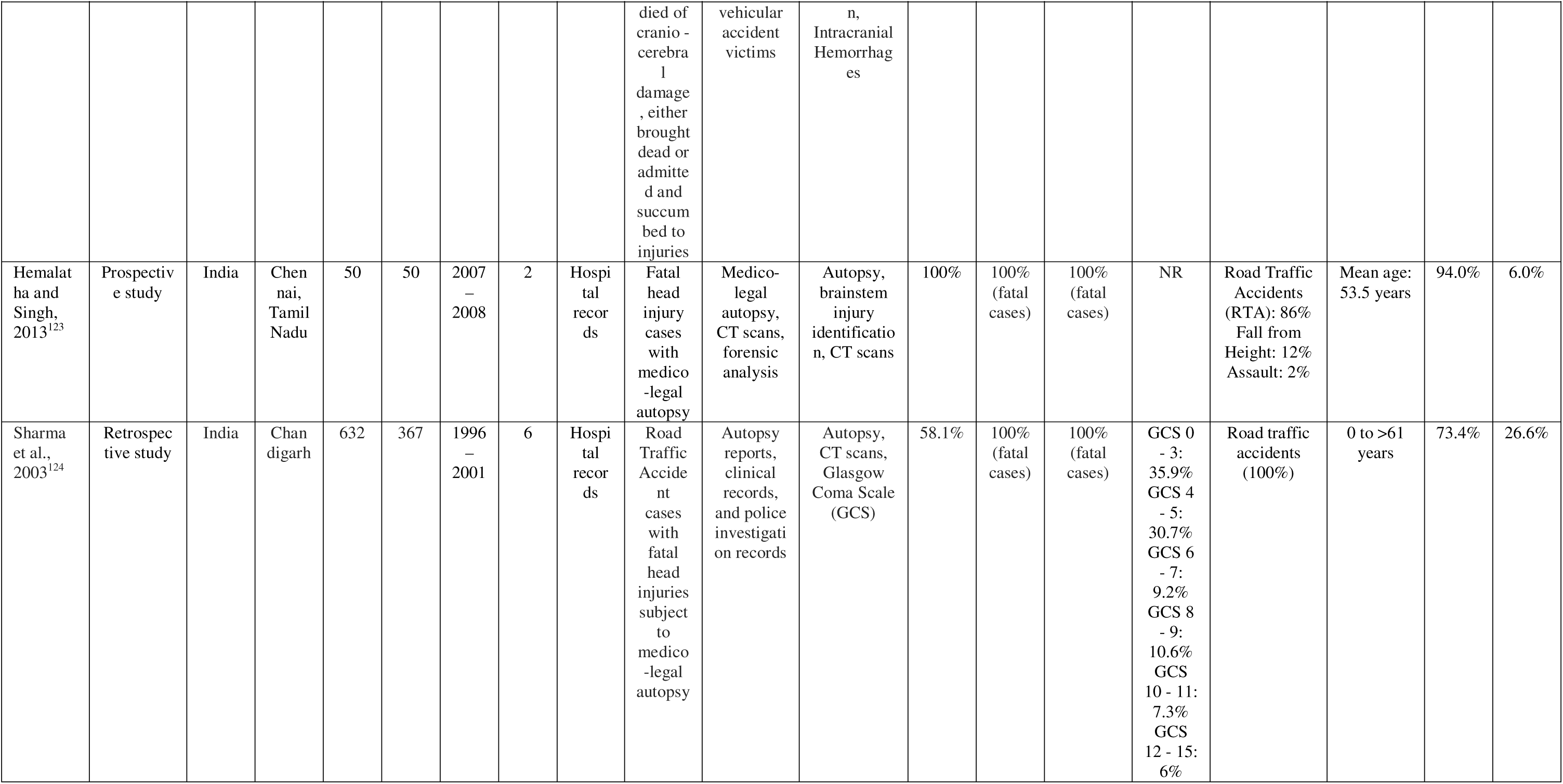
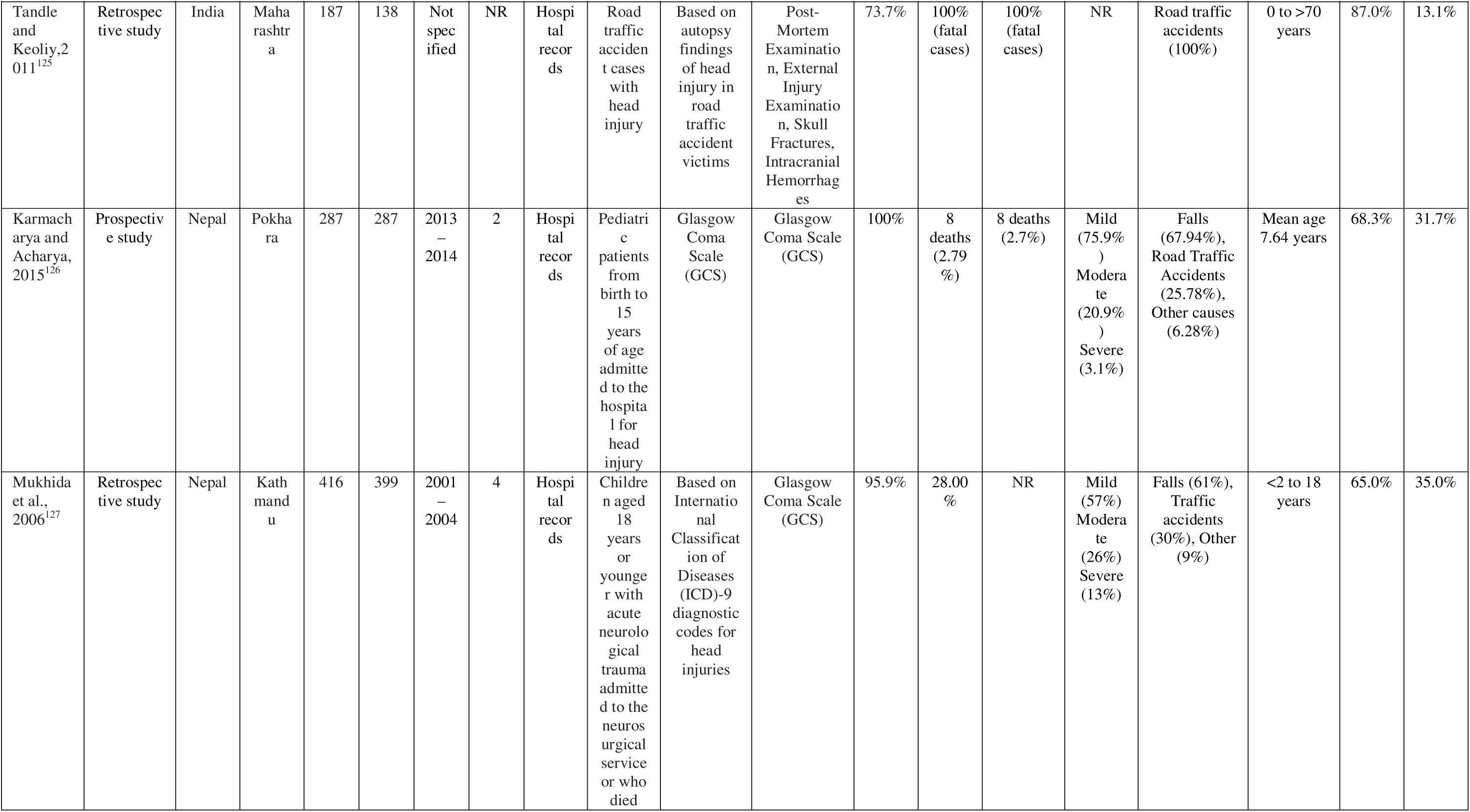
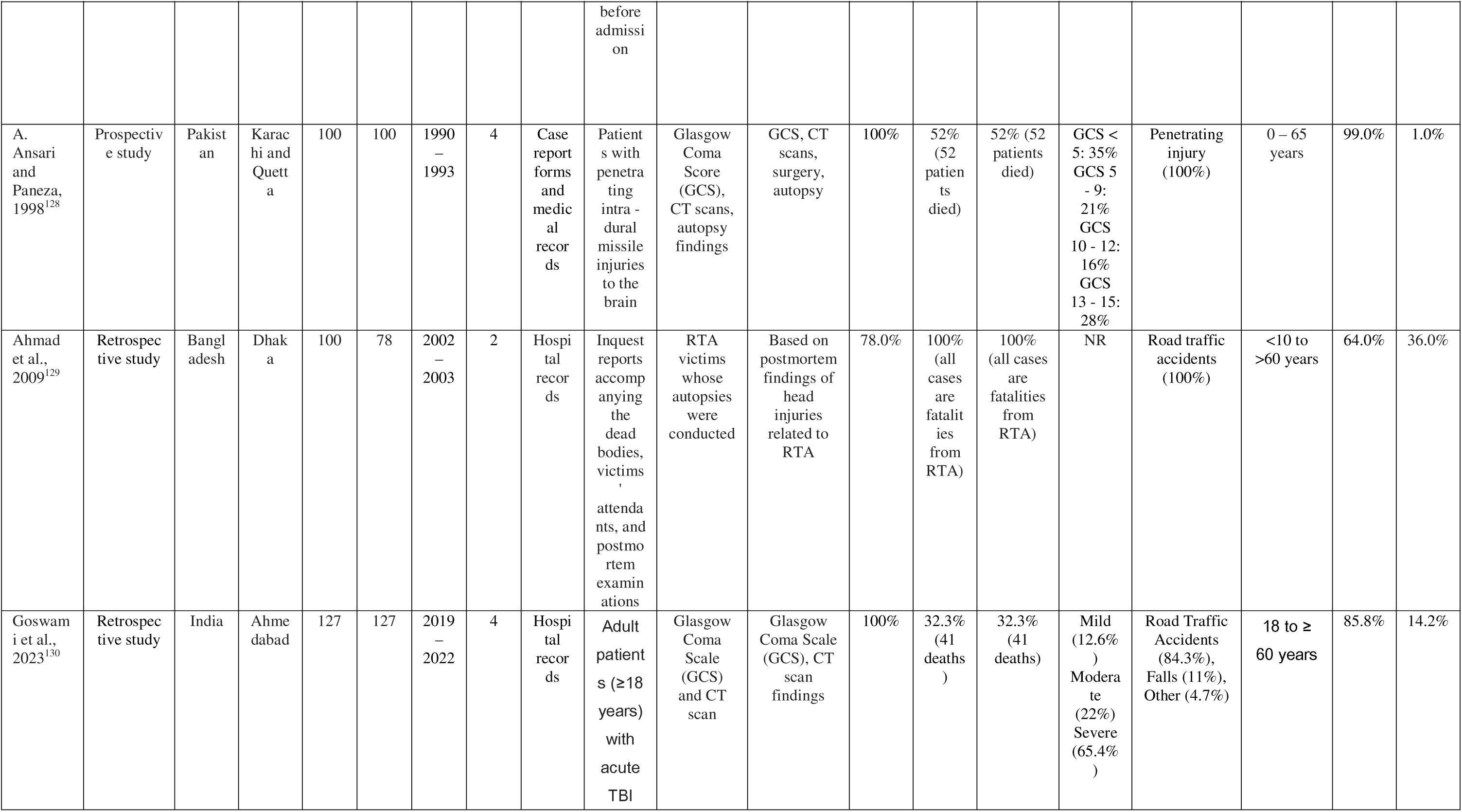
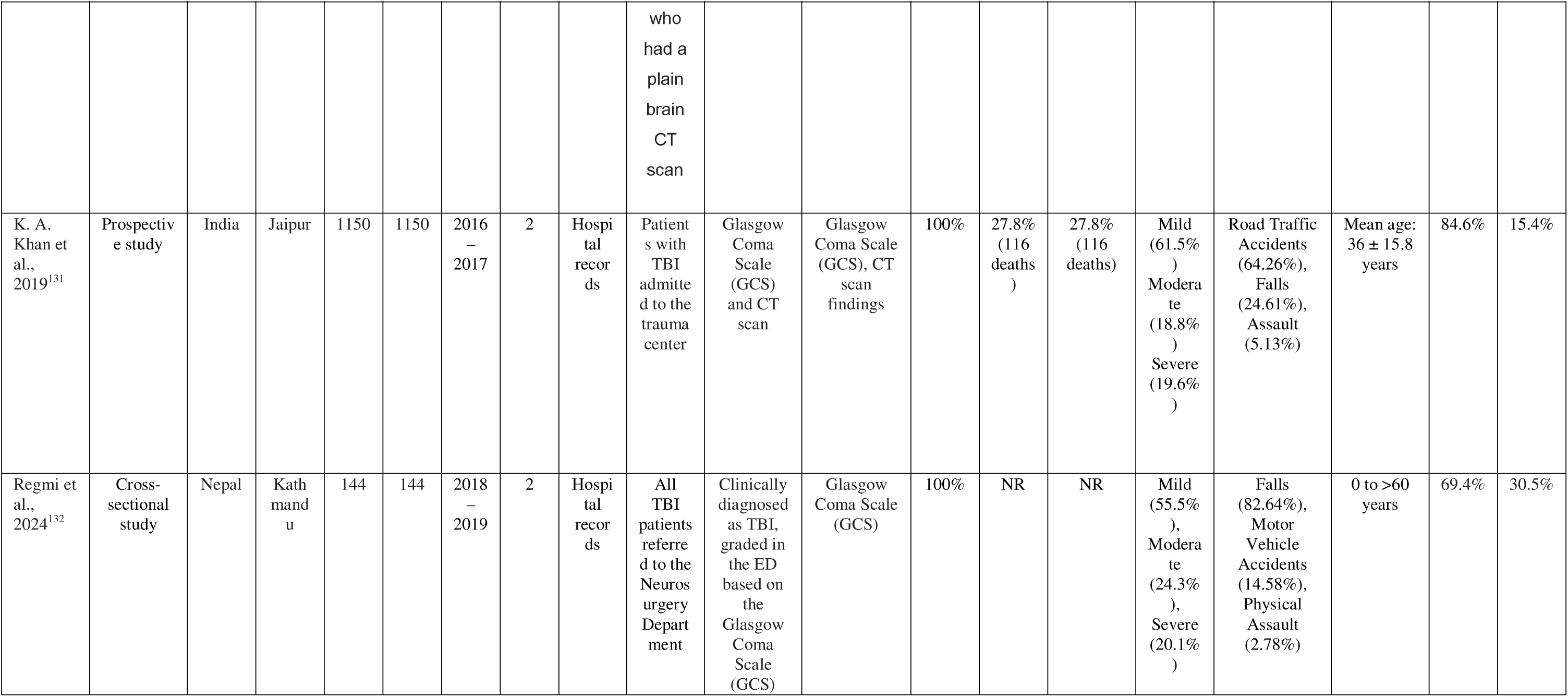
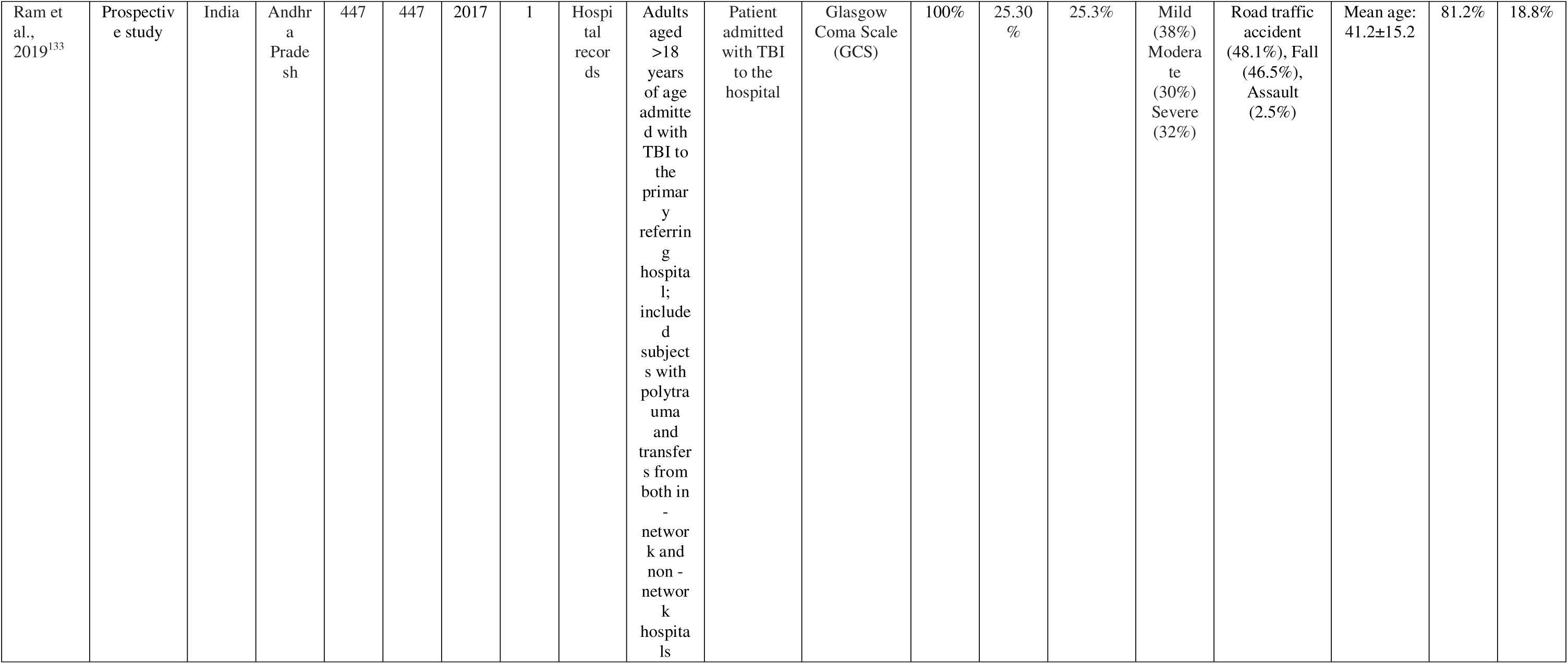
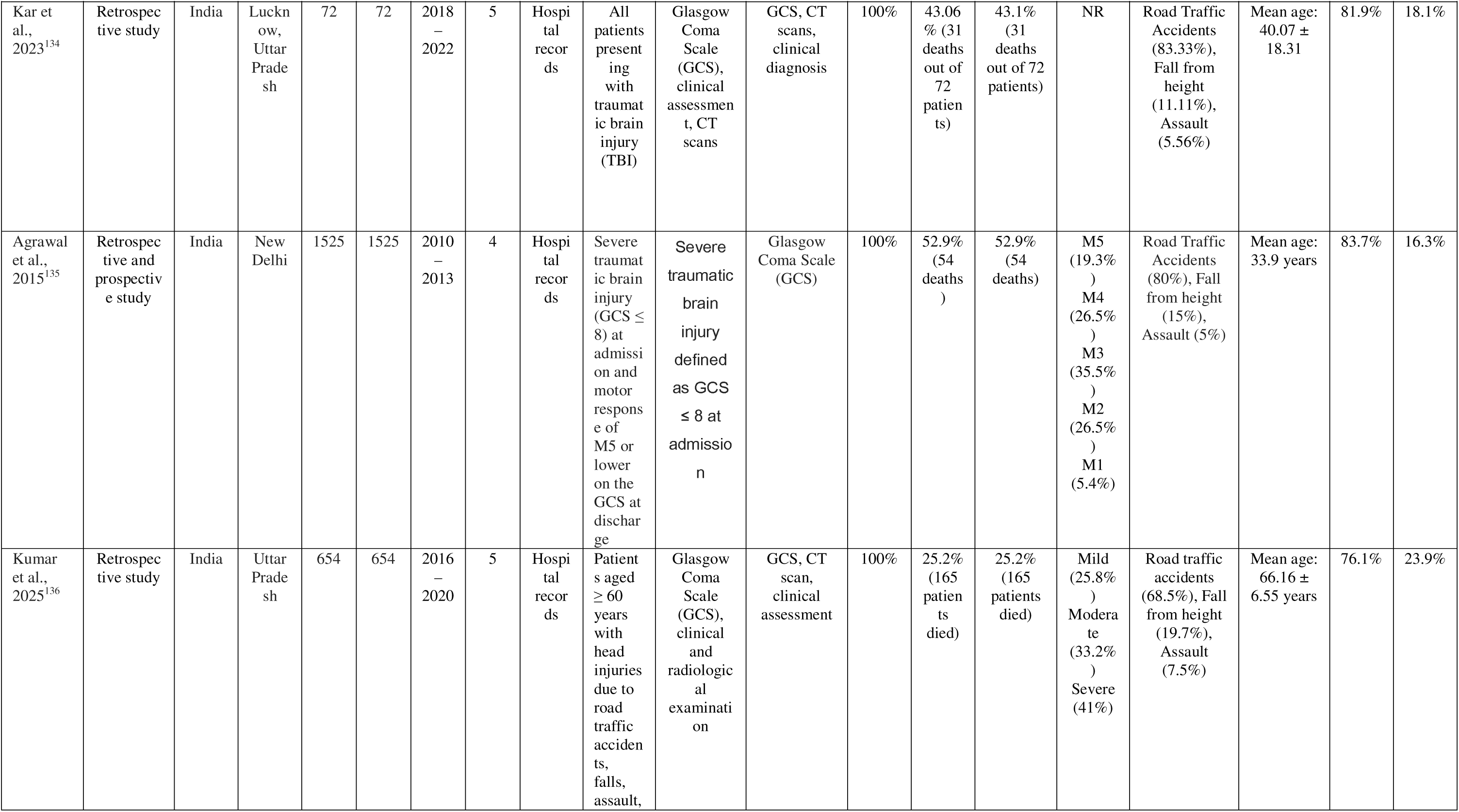
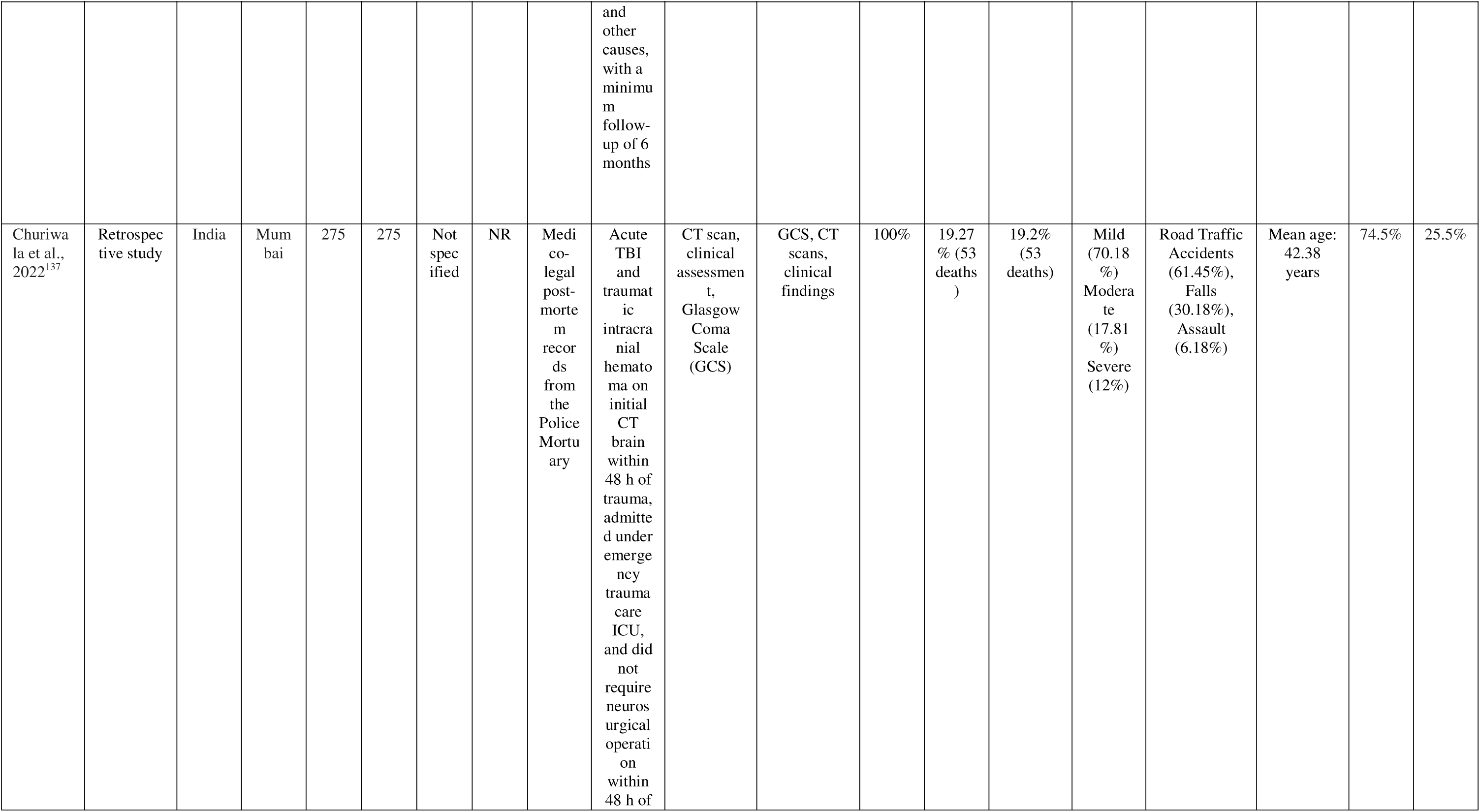
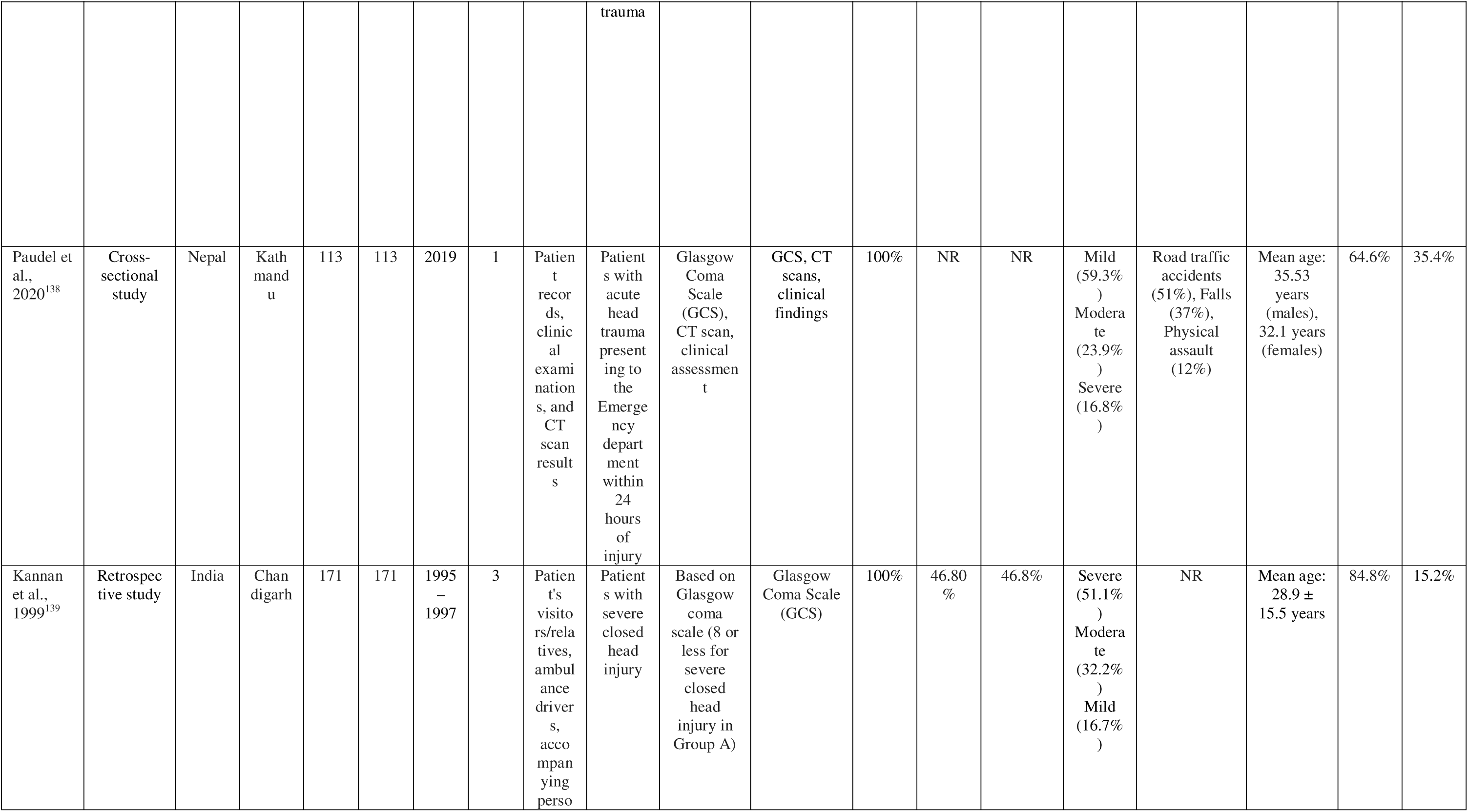
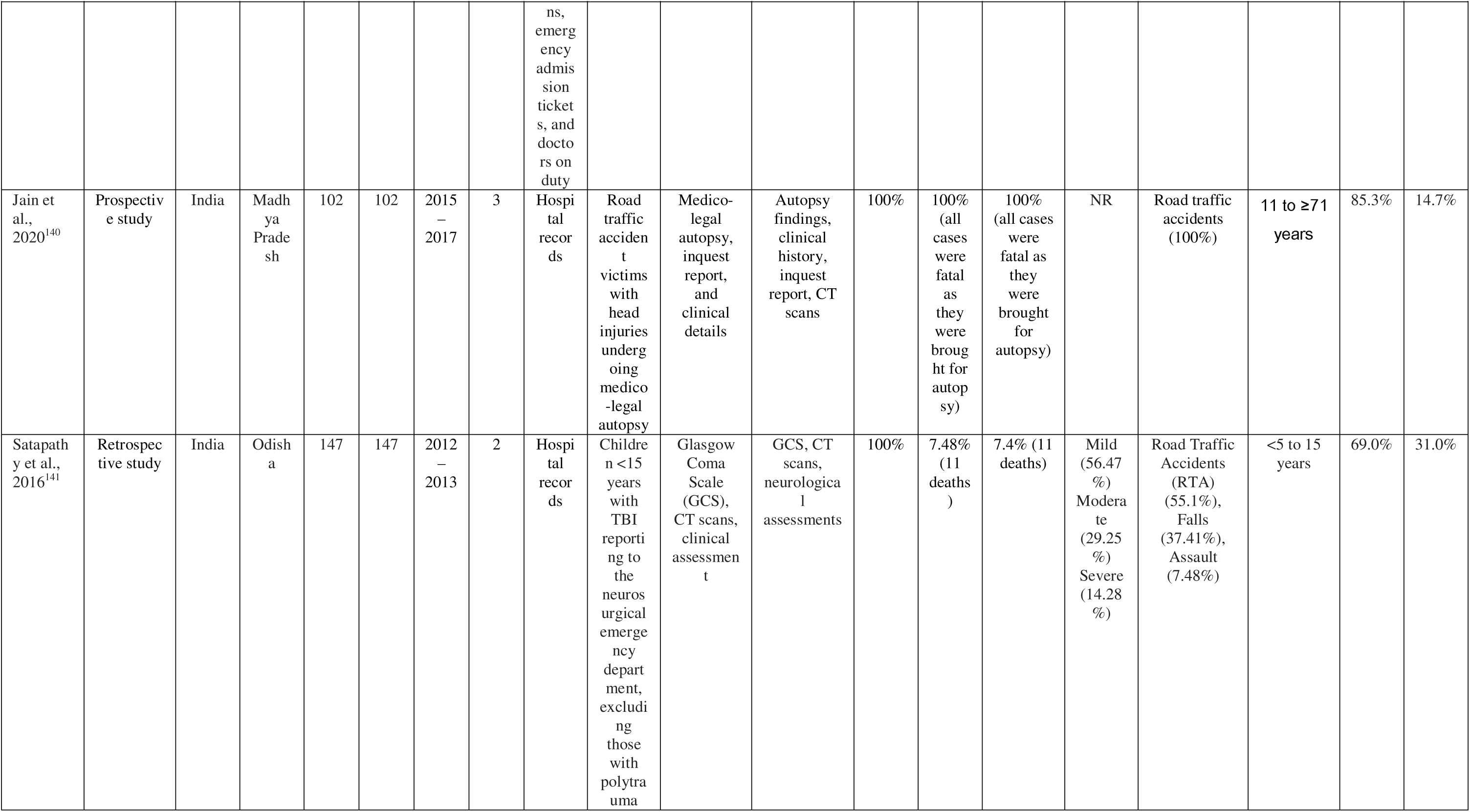
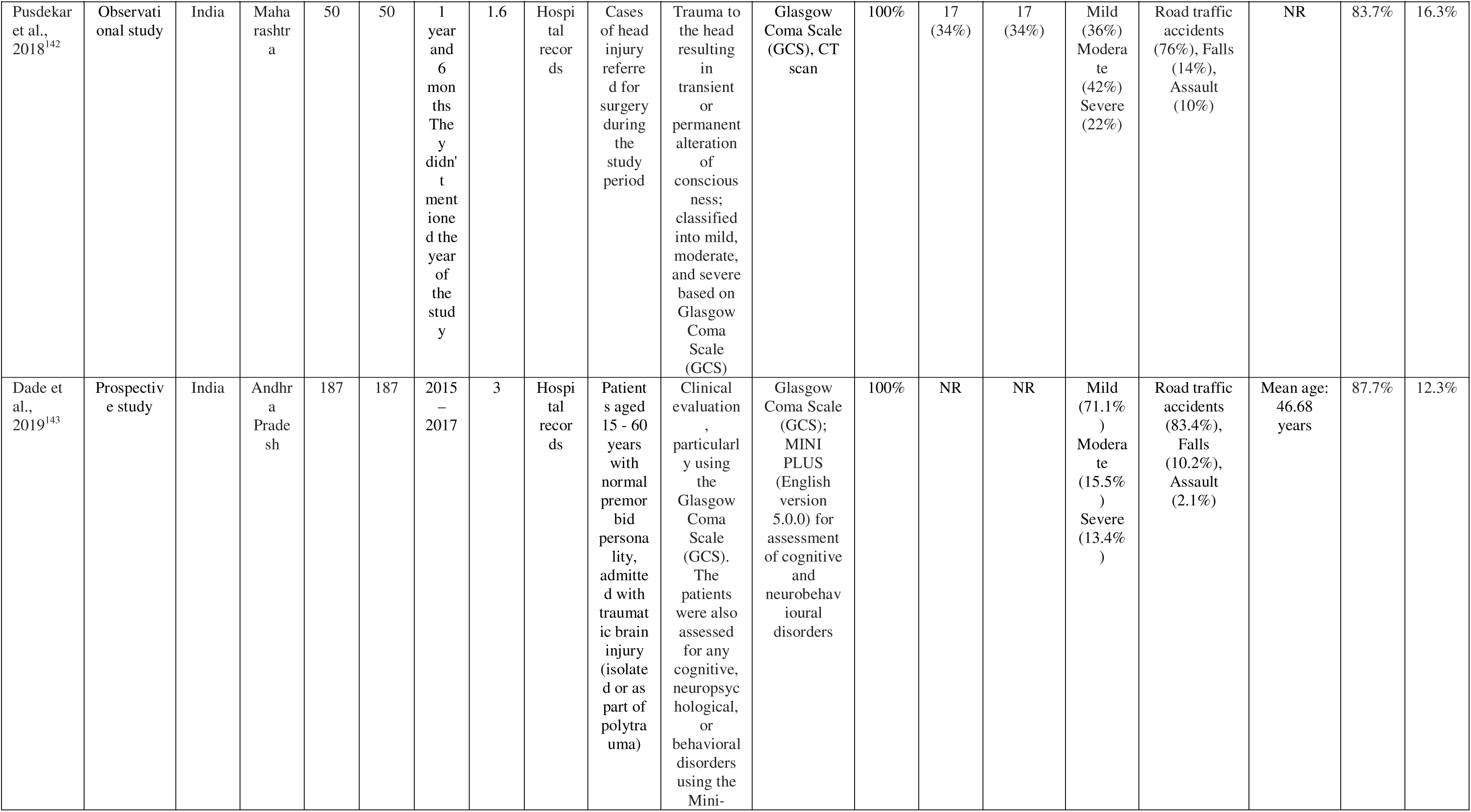
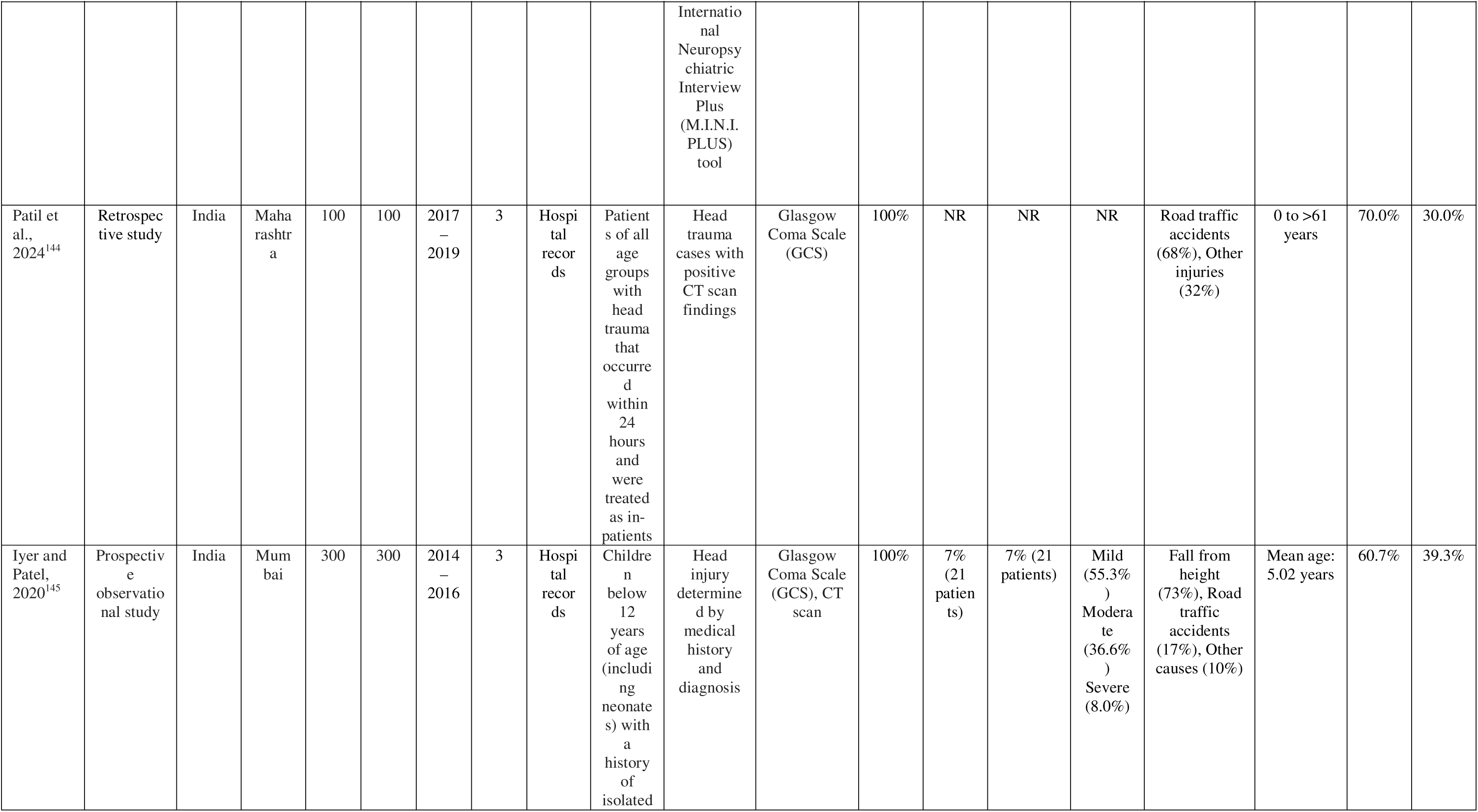
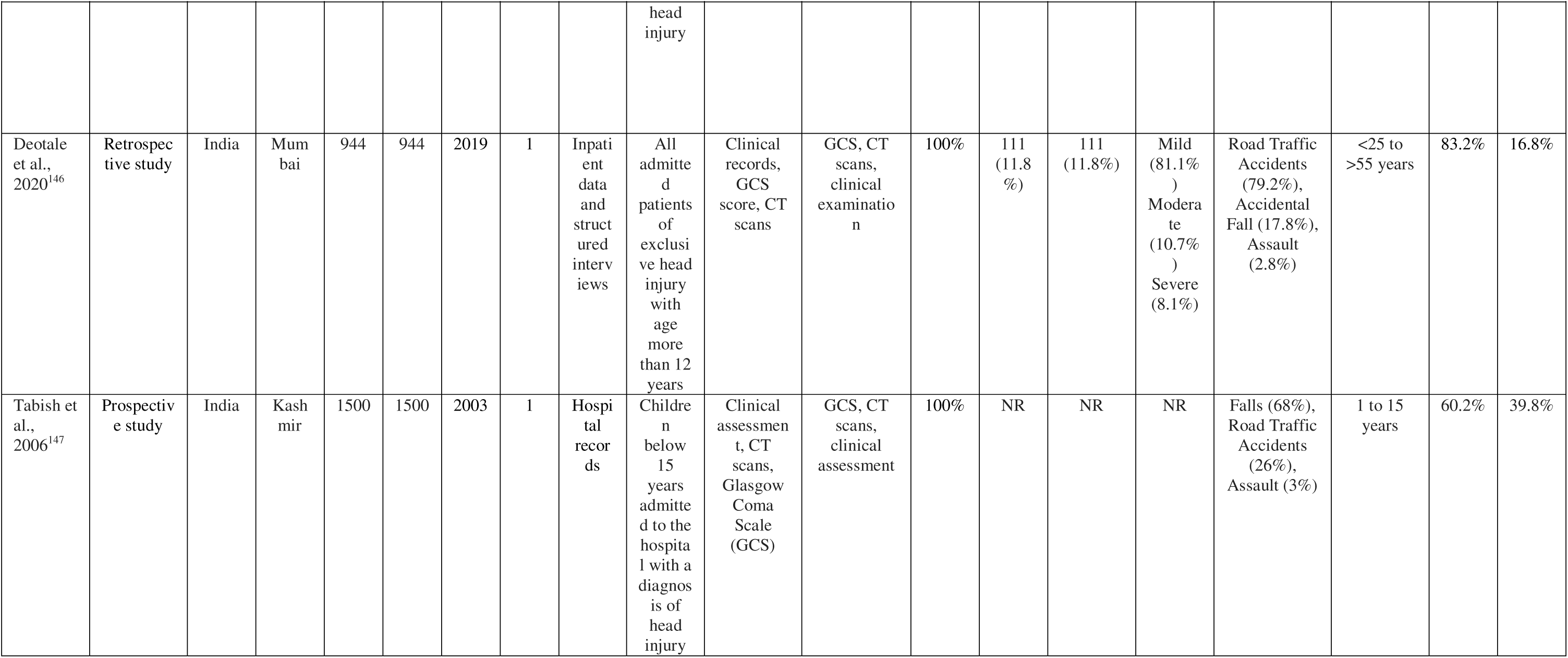
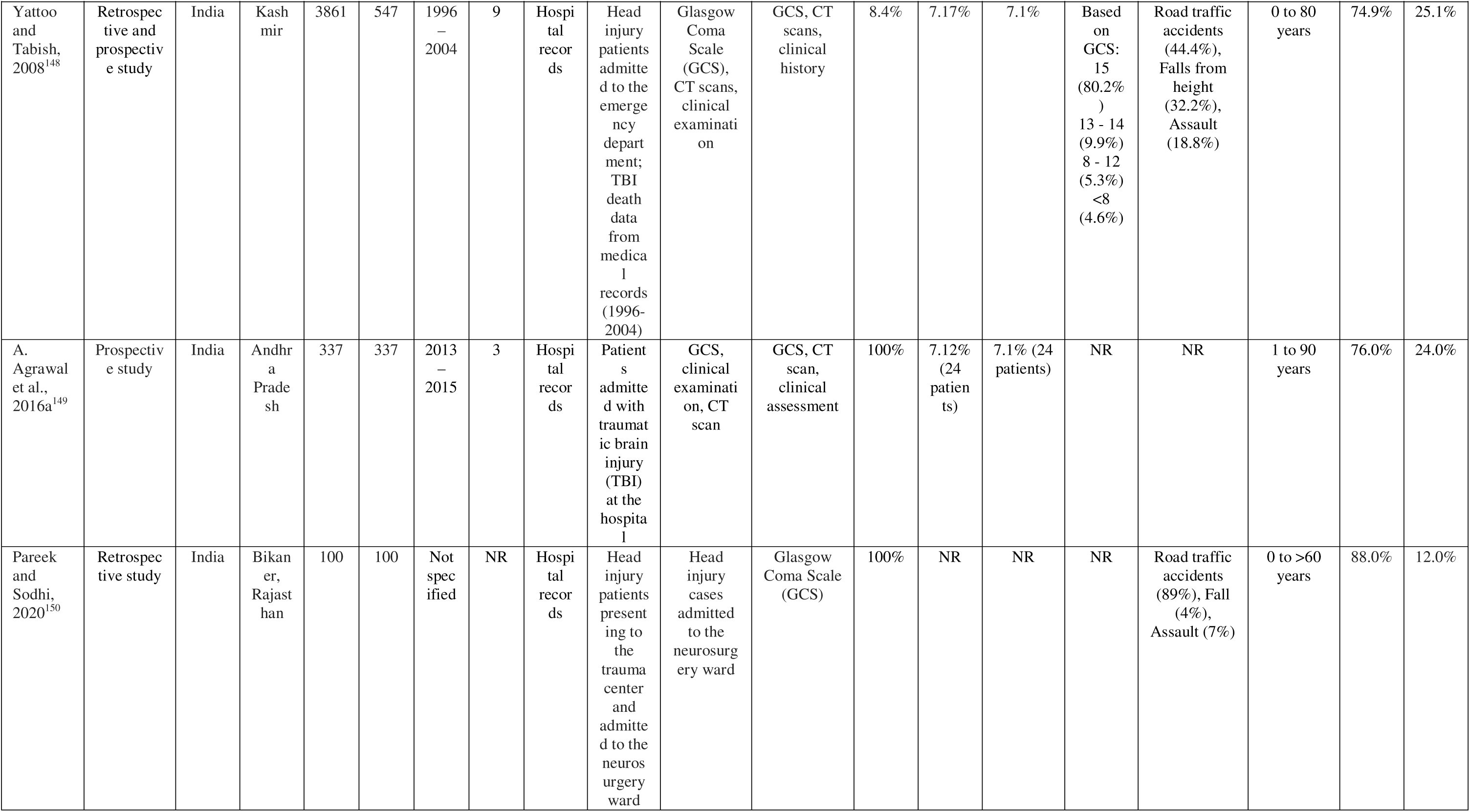
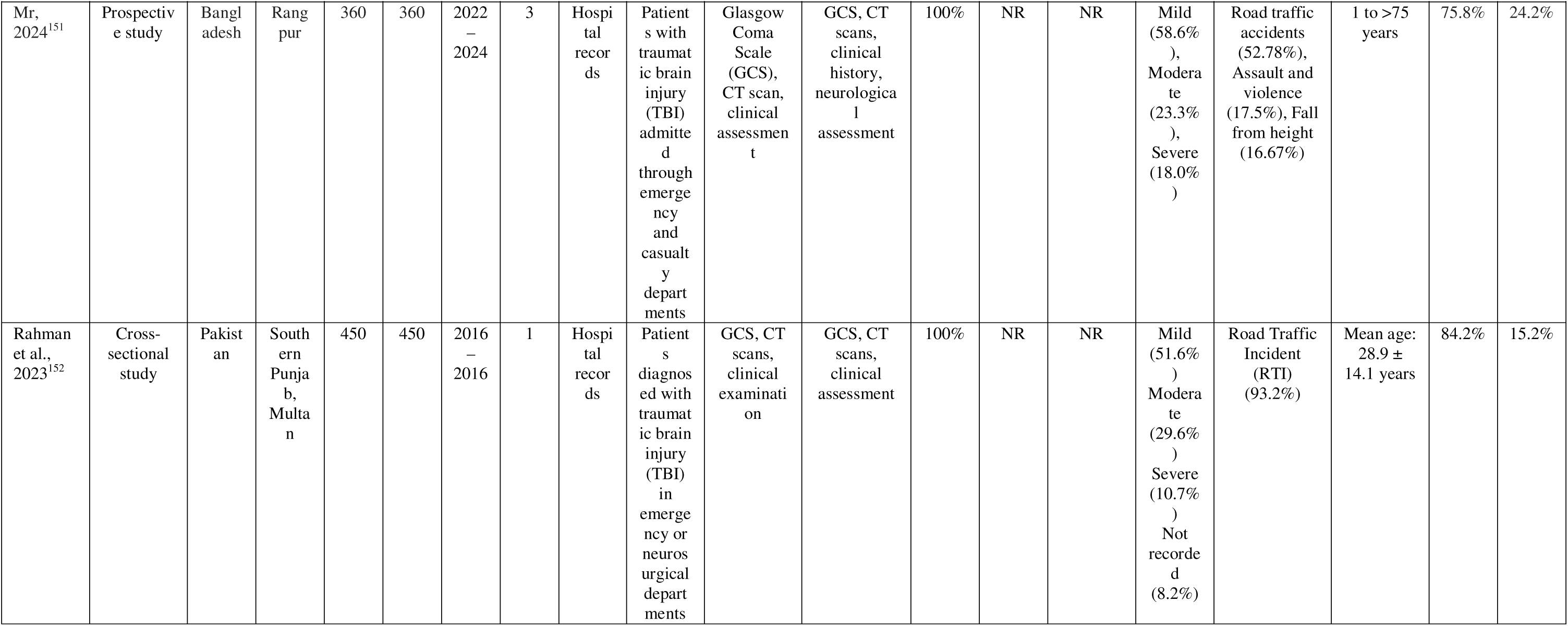
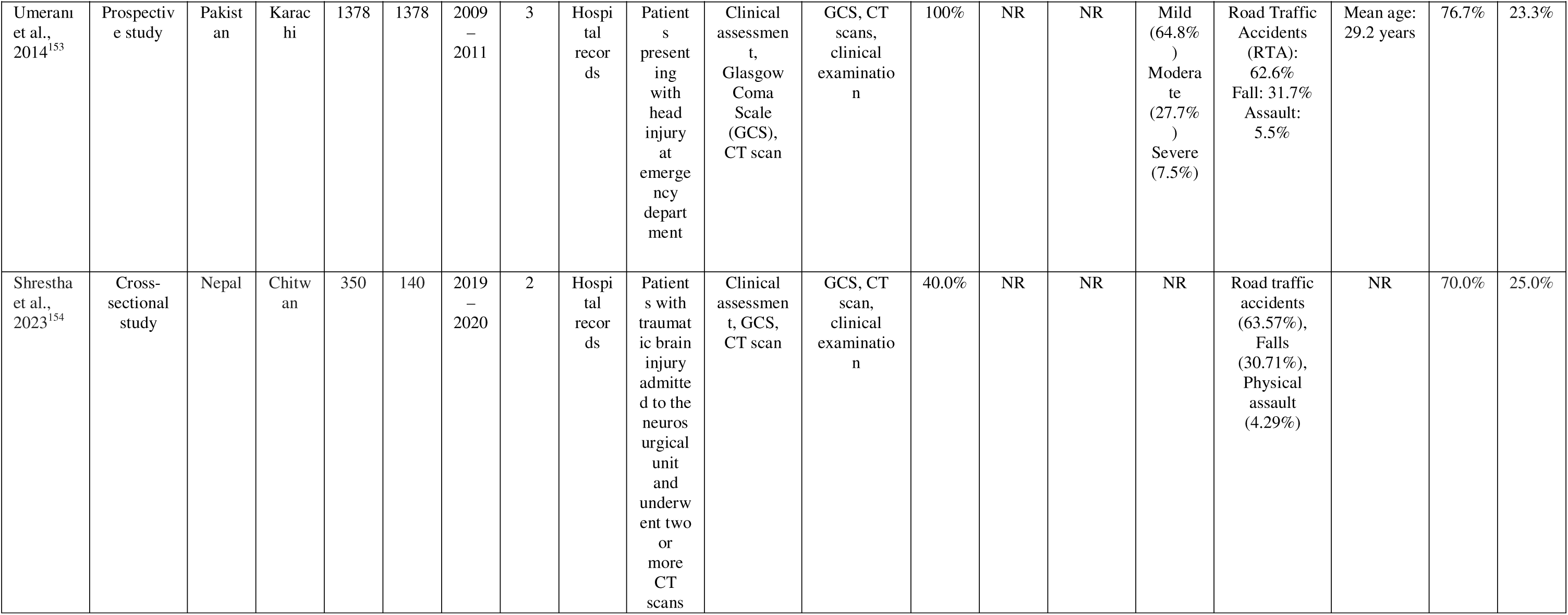
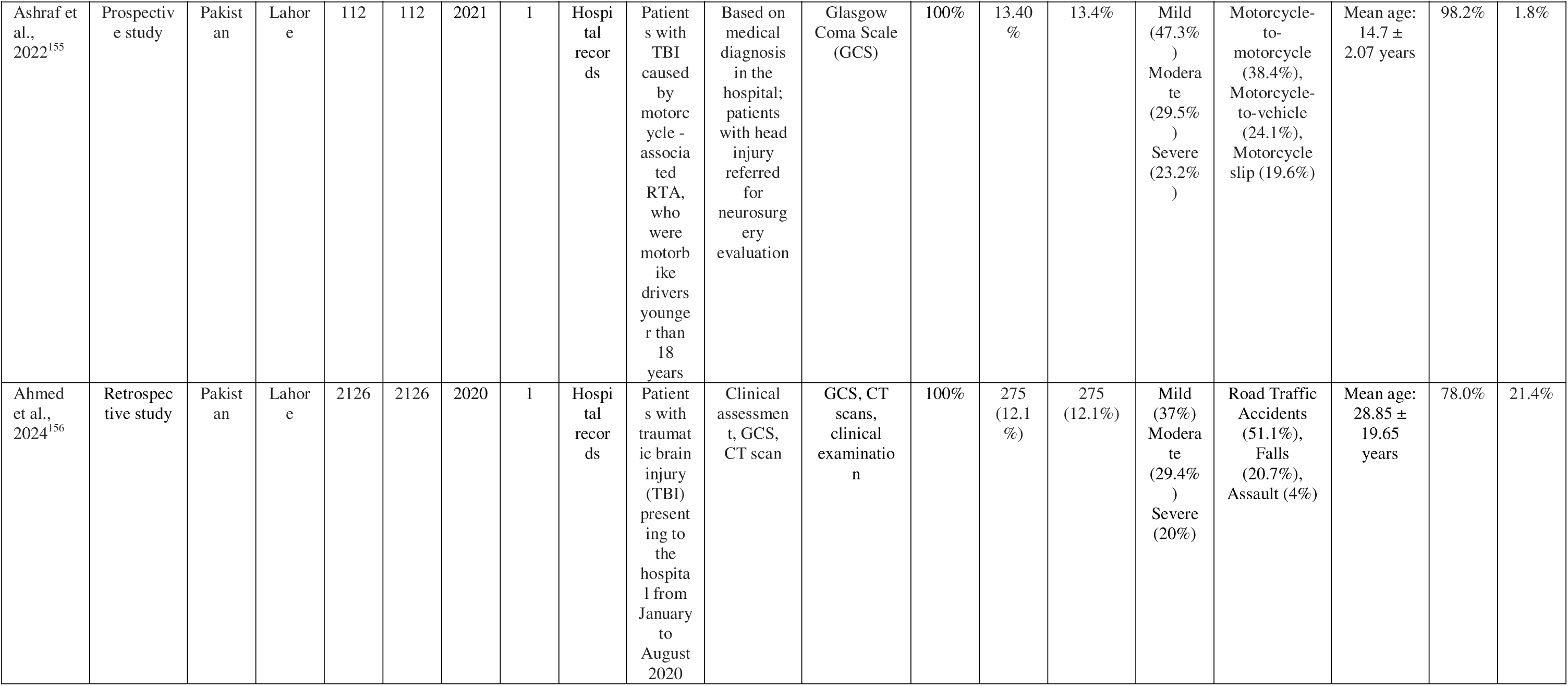
Characteristics of the included studies.

### Country Variation

The revies included studies from Bangladesh, India, Nepal, Pakistan and Sri Lanka. Studies from three more countries in South Asia, Afghanistan, Bhutan, and Maldives, were not part of this research. Though they were under consideration for inclusion, no articles from these countries met predetermined inclusion criteria. These countries thus did not represent in this final list of included studies for review.

### Sample Size

The extensive range in sample size (from 10 up to 260,000) indicates heterogeneity in research scope on a large scale.

### Study period

A total of 126 out of 130 studies in this research reported the duration for which data were collected. The average duration study periods were about 2.66 years. The study periods ranged from 1 to 22 years, showing variation in observation periods. Most studies focused on a relatively brief observation period. With regard to frequency, 1–3 year durations were most common. Among them 42 studies reported 2 years, 29 studies reported 1 year and remaining 29 studies had 3-year duration. These three durations contributed more than 78% of studies with valid duration data and demonstrate a strong tendency towards short-term investigations. Conversely, extended-term studies were rare. Only 11 studies reached 4 years, 9 studies reached 5 years, 2 studies reached 6 years and only 2 studies extended to 9 and 22 years, constituting an unusual instance of lengthy surveillance.

### Data sources

Data for most came from hospital records, including medical histories, patient records, and treatment charts. Autopsy records and reports came into play to evaluate fatal cases in many instances. Police records in the form of inquest reports and dead body challans added to the clinical data. Structured questionnaires, self-structured proformas, clinical examination, and CT scan reports were among other data sources. Telephone interviewing was also included by some studies.

### Inclusion Criteria

This study illustrates considerable heterogeneity among the studies, mirroring diversity in patient selection for TBI cases based on age, injury severity, clinical setting, and injury mechanism. Although some studies confined themselves to moderate to severe TBI alone, others included mild head injury cases as well as cases from all severities. Inclusion criteria differed by age groups as well, with some studies focusing on pediatric populations (<18 years), while others targeted adult and older populations (>60 years). These age-based subgroup distinctions help elucidate how age plays a role in susceptibility to as well as recovery from TBI and are important when interpreting age-specific risk factors and outcome patterns. Furthermore, numerous studies select based on clinical admission category, such as those admitted to ICUs or neurosurgical departments. Mechanisms involving road traffic accidents (RTAs) and falls were readily listed as inclusion criteria. These are common injury mechanisms in South Asia. The time since injury was another inclusion criterion with considerable heterogeneity. While most studies included acute TBI cases within 24 to 48 hours post-injury, others included patients with a more prolonged duration of injury from any hour to days post-injury and even included surgical cases. Such broad variation in inclusion could affect generalizability and comparability among studies.

### TBI definition/ Case ascertainment

The severity of TBI was assessed primarily through Glasgow Coma Scale (GCS) and supplemented regularly by CT scans, X-rays, and MRI scans. Clinical examination and autopsies were included as part of many studies to determine injuries such as skull fractures and intracranial hemorrhages. Furthermore, tools including Glasgow Outcome Scale (GOS), Kampala Trauma Score (KTS), TRISS, and FOUR score were used to quantify severity and outcome.

### Methodological quality

Based on the MORE checklist, all included studies described their aims clearly (99.2%) and used proper study designs (100%), while ethical approval and conflict of interest were reported well (91.5% and 84.6%, respectively). Funding information reporting was poor (96.2%), while sampling exhibited significant flaws with 98.5% receiving minor flaws ratings. Definitions for cases and classifications for TBI severities were good (>93%), but management of bias was a critical flaw and 99.2% exhibited major flaws. Data sources and subject flow were reported well (97.7%). Reliability of estimates and reporting of mortality were less good with more than 60.8% and 24.5% exhibiting flaws, respectively.

**Table 2.**
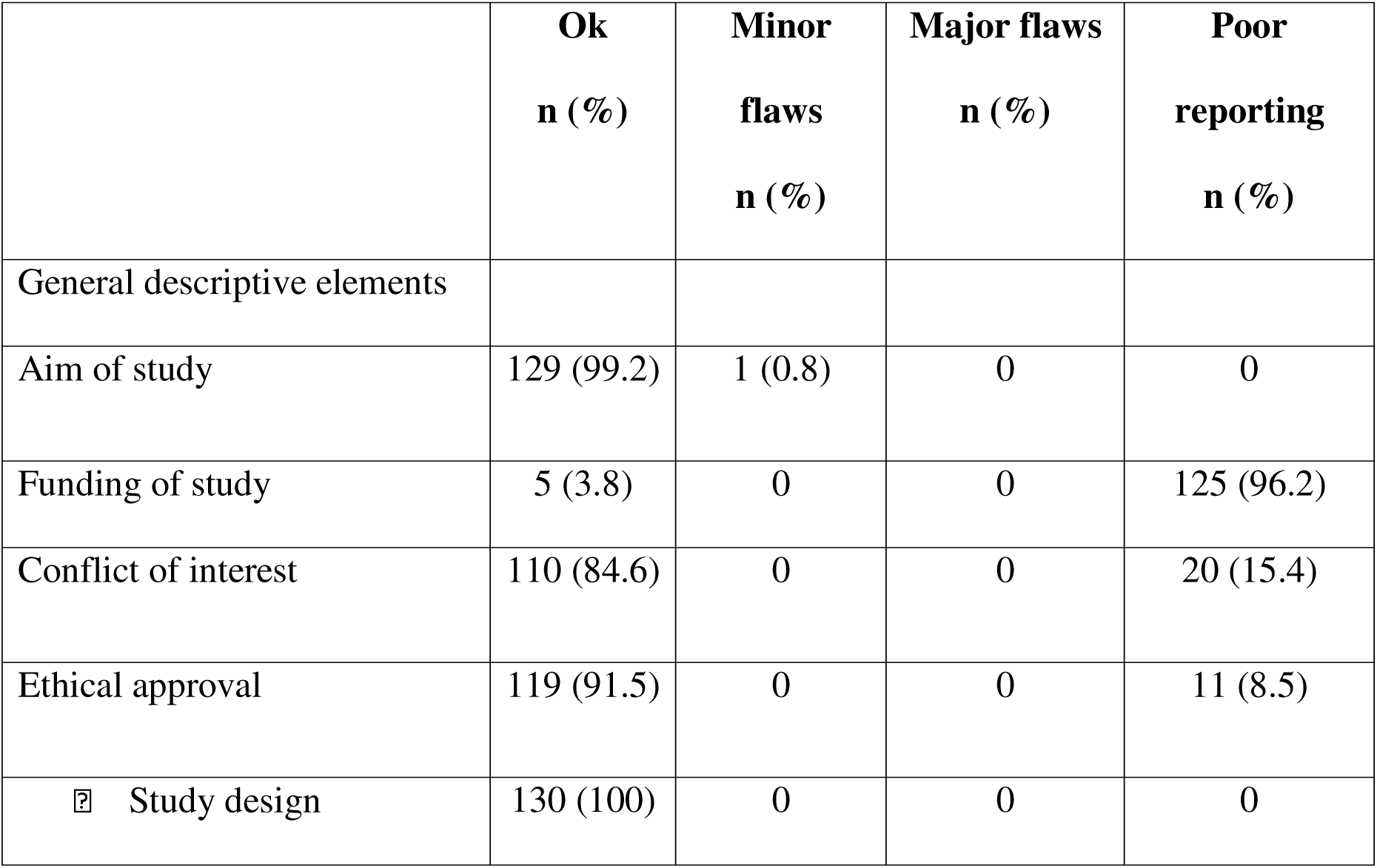
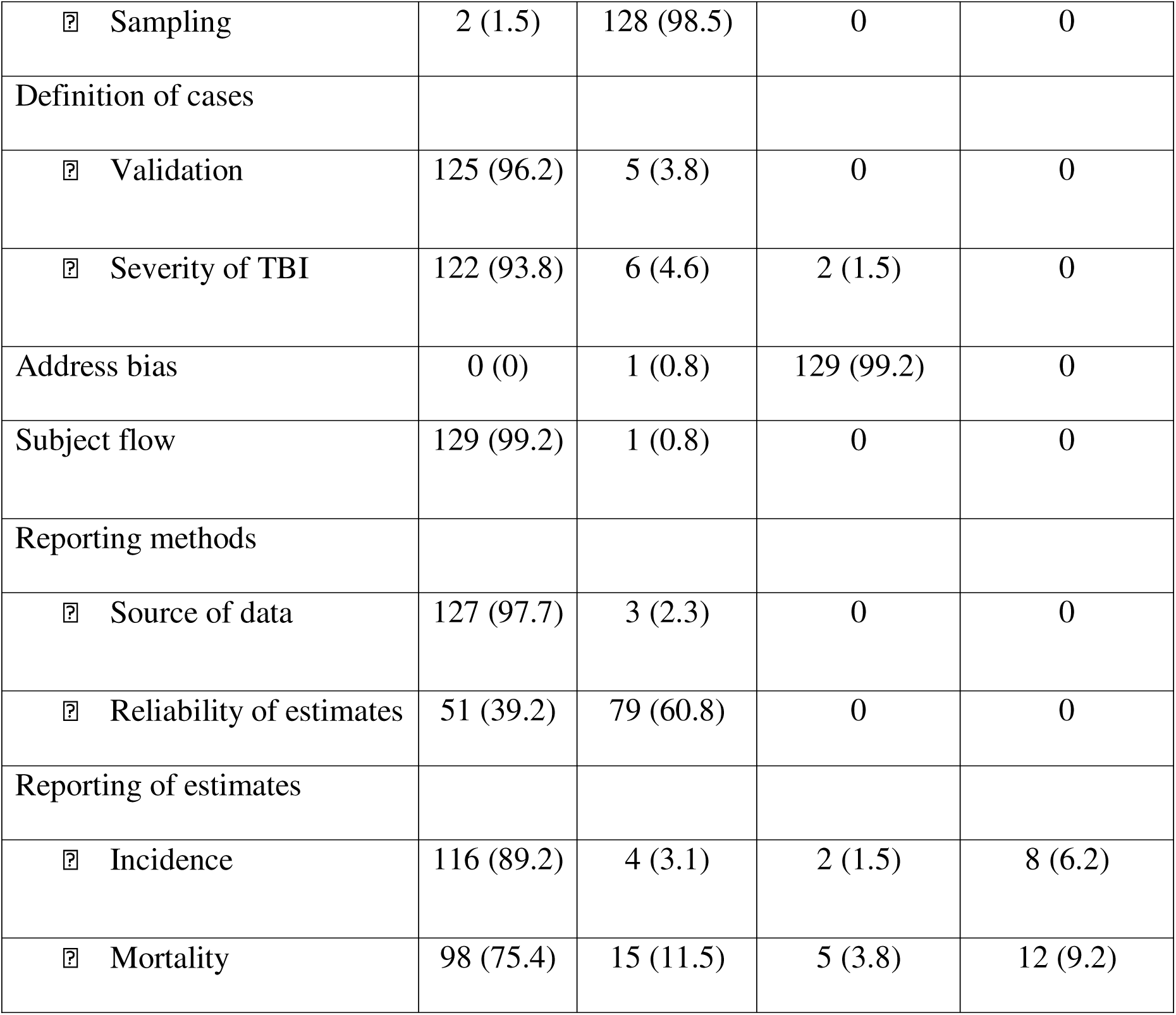
Summary results of quality assessment of included studies using Methodological Evaluation of Observational Research checklist (MORE) checklist.

|  | **Ok**<br>**n (%)** | **Minor**<br>**flaws**<br>**n (%)** | **Major flaws**<br>**n (%)** | **Poor**<br>**reporting**<br>**n (%)** |
| --- | --- | --- | --- | --- |
| General descriptive elements |  |  |  |  |
| Aim of study | 129 (99.2) | 1 (0.8) | 0 | 0 |
| Funding of study | 5 (3.8) | 0 | 0 | 125 (96.2) |
| Conflict of interest | 110 (84.6) | 0 | 0 | 20 (15.4) |
| Ethical approval | 119 (91.5) | 0 | 0 | 11 (8.5) |
| □ Study design | 130 (100) | 0 | 0 | 0 |
| ☐ Sampling | 2 (1.5) | 128 (98.5) | 0 | 0 |
| Definition of cases |  |  |  |  |
| ☐ Validation | 125 (96.2) | 5 (3.8) | 0 | 0 |
| ☐ Severity of TBI | 122 (93.8) | 6 (4.6) | 2 (1.5) | 0 |
| Address bias | 0 (0) | 1 (0.8) | 129 (99.2) | 0 |
| Subject flow | 129 (99.2) | 1 (0.8) | 0 | 0 |
| Reporting methods |  |  |  |  |
| ☐ Source of data | 127 (97.7) | 3 (2.3) | 0 | 0 |
| ☐ Reliability of estimates | 51 (39.2) | 79 (60.8) | 0 | 0 |
| Reporting of estimates |  |  |  |  |
| ☐ Incidence | 116 (89.2) | 4 (3.1) | 2 (1.5) | 8 (6.2) |
| ☐ Mortality | 98 (75.4) | 15 (11.5) | 5 (3.8) | 12 (9.2) |

### Prevalence

In this systematic review, 130 selected articles presented different prevalence rates for traumatic brain injury (TBI). While most study reported 100% prevalence on the selected study population, each representing uniform occurrence among the study populations, others reported prevalence rates ranging from 8.4% to 95.9%.

### Mortality and case fatality rates

A total of 101 studies reported mortality rates. Mortality in studies included in this study varied significantly, as expected given differences in study design, target populations, and case severity being studied. Total 29 studies did not report mortality data.

### Type of severity

The severity distribution of traumatic brain injuries (TBI) across the studies shows considerable variation. A total 95 studies reported severity data with detailed categorizations. The most frequent distribution shows a focus on mild, moderate, and severe injuries, with varying proportions in different studies.

### Mechanism of TBI Injury

The injury mechanism for TBI is dominated strongly by road traffic accidents (RTAs) as the predominant source of injury in South Asia. There were 12 studies, and they reported 100% of cases attributed to RTAs,^33,45,94,111,116,117,122,124,125,129,140,157^ confirming road traffic injury’s high burden in the region. Aside from RTAs, some studies included fall from height as a major source of injury. Violence or assault as a factor was reported by some studies but less commonly included.

### Age and sex

The average age of participants in studies included in this systematic review is highly variable and indicates diverse populations under investigation. The age in studies ranges from pediatric populations with average ages as low as 4.7 years to elderly populations with average ages as high as 71.08 years. The majority of studies target younger populations with ages in their 20s and 30s as indicated by commonest average age values of 24 and 28.9 years. Nevertheless, a few studies target adolescents and children with average ages of 15 and 4.7 years as an indication that pediatric TBI is their focus. The gender split throughout the studies is dominated by male participants, and most studies report 70% and above male participants. For instance, 78% is the most frequent male proportion, and most studies reveal male proportions ranging from 65% to 88%. Female proportions tend to be a smaller percentage and usually 22% or below.

## Discussion

This systematic review fully explored the epidemiology of traumatic brain injury (TBI) in South Asia. Despite significant methodological limitations in the primary studies, suggests that TBI is a major public health concernin the region as indicated by high rates of prevalence among young adults. Road traffic accidents (RTAs) were found to be the primary source of TBI in countries in South Asia as a whole, including Bangladesh, India, and Nepal which recorded especially high rates involving traffic injuries leading to TBI. Mortality differed, however, the burden of TBI as indicated by case fatality and disability over a prolonged period after injury was high. Our systematic review further highlighted that severity ranges of TBI in the region were mostly found to be mild to moderate while a large number of cases also presented with severe injuries.

This systematic review indicated that TBI was a considerable public health problem in South Asia with variable prevalence rates among countries. The prevalence of TBI within the region was significantly high, particularly among countries with inadequate infrastructure and poor road safety policies.^158^ The rapid urbanization and motorization in South Asia further fueled the cases of TBI and road traffic accidents (RTAs) were predominant causes in most studies.^155^ TBI is more common in other regions, such as United States. In a systematic meta-analysis including a general adult population, about 18.2% (95% CI 14.4-22.7%) adults reported a lifetime history of TBI with loss of consciousness.^159^

The case and mortality rates due to TBI in South Asia were very high. There was wide variation in these rates in the systematic review, with severe cases of TBI being a major driving factor for mortality. Traumatic brain injury is also a major cause of death and disability worldwide despite accurate worldwide mortality rates being highly variable according to geographical region, healthcare system, and reporting policies. Mortality due to TBI is calculated in a systemic review to be 20-30% worldwide.^160^ The crude mortality rates in Europe ranged from 9 to 28.1 per 100,000 population per year.^161^ A study discovered that TBI now stands as the leading cause of mortality in instances of polytrauma as evidence indicates a change in patterns among trauma-related deaths.^162^ They reported 66% (70/106) among all trauma-related mortality.^162^

In terms of their severity, most cases of TBI among South Asians were mild or moderate but a large percentage also included severe injuries. The distribution according to severity differed among countries and regions and an area might record a greater percentage of severe injuries. This inconsistency might have been due to variations in case ascertainment approaches that involved use of Glasgow Coma Scale (GCS) scores and high-tech imaging modalities such as CT and MRI scans. The injury severity in TBI was found to be directly associated with outcome, and the results indicated more severe injury in most instances leading to an extended duration of stay in the hospital, higher healthcare expenditure, and prolonged disabilities.

The major mode of TBI throughout South Asia was road traffic accidents (RTAs), followed by falls and assaults. RTAs specifically contributed to more than half of the cases with motorcycles being the leading vehicle implicated in TBI. These study results coincide with other studies. A meta-analysis LMICs reported 39% pediatric TBI cases due to RTAs.^19^ In Ethiopia, 21% of TBI cases were due to RTAs.^163^ A bibliometric study involving research on TBI indicated RTAs as the most common source of TBI over falls among older populations in line with the evidence.^160^ The predominance of RTAs in South Asia was driven by inadequate road infrastructure,^164–166^ poor protective precautions such as helmet.^167^

TBI affected all ages but varied significantly depending on age and sex. This systematic review found high incidences among young adults and especially males that align with results in previous studies. One study found that young adults and especially men and older people are high-risk groups for TBI.^168^ In line with a systematic review and meta-analysis involving all major studies up to 2023, males account for about 74.3% of all cases of TBI and create a male-to-female ratio close to 3:1.^169^ One meta-analysis found that prevalence among genders differed and that males recorded a higher percentage at 20.8%.^159^ Several studies have discovered age as a major factor in determining TBI outcome. In a recent systematic review and meta-analysis, age is a major determinant with an average age gap of 8.72 years among patients.^170^

This review utilized multiple databases to ensure comprehensive coverage of studies from various countries from the region of South Asia. It also had specific research aims centred on important TBI epidemiologic outcomes so a meaningful synthesis could be achieved. This review utilized the PRISMA approach and also employed a rigorous data extraction method for ensuring methodological quality. By considering different study designs like cross-sectional, cohort, and case-control studies, it captured data from a variety of settings from South Asia. The application of the MORE checklist for assessing study quality assured even more rigor and ensured the incorporation of accurate studies.

Major limitations of the present systematic review were the considerable heterogeneity in included studies in relation to study design, data sources, case definitions, TBI measurement method which hampered quantitative synthesis. Dissimilarities in the report of TBI outcomes like prevalence, mortality, severity, and mechanism of TBI injury also hampered comparison. The quality of the studies varied methodologically in relation to sampling and case and data reliability biases. Longitudinal follow-up data were missing from most studies, hence limited the estimation of outcomes like disability or recovery.

TBI in South Asia presented as a severe public health concern with high prevalence, high mortality, and variable severity and was most commonly due to road traffic accidents. This research results highlighted greater need for better infrastructure for trauma care, preventive efforts towards road safety, and recognition towards high-risk groups including young adults, children and older adults. Priority policy interventions towards strengthening road safety, increasing numbers of trauma care facilities, and enhancing data collection were necessary to reduce the burden of TBI in the region. Future studies should incorporate standardized approaches to collecting data on TBI and longitudinal studies to further understand late consequences and outcomes for survivors of TBI.

### Contributors

SH and SCP conceptualized the study. SH, SCP, RH, and MEH performed title, abstract, and full-text screening. FS and KMYA conducted data extraction, and MMM and JAI verified the extracted data. SH, RH, FS, KMYA, MMM, and JAI synthesized the results. SH, RH, MMM and JAI assessed the risk of bias. SH drafted the manuscript, and SCP critically reviewed it for intellectual content. All authors read and approved the final manuscript, jointly decided to submit the paper, and agreed to be accountable for all aspects of the work.

### Data sharing statement

The findings of this study are based entirely on data from previously published studies, all of which are publicly accessible and referenced within this manuscript. No new primary data were collected or analyzed in this systematic review. Additional information or detailed data extraction materials used in this review can be provided upon reasonable request to the corresponding author.

## Supporting information

S1 Appendix: Search String

S2 Appendix: Methodological Evaluation of Observational Research (MORE) Checklist for Quality Assessment of Included Studies

## Data Availability

https://www.crd.york.ac.uk/PROSPERO/view/CRD42022364511

## Acknowledgment

We thank Marzana Afrooj Ria, our information specialist, for her assistance with the search strategy.

## Appendices

### S1 Appendix: Search String

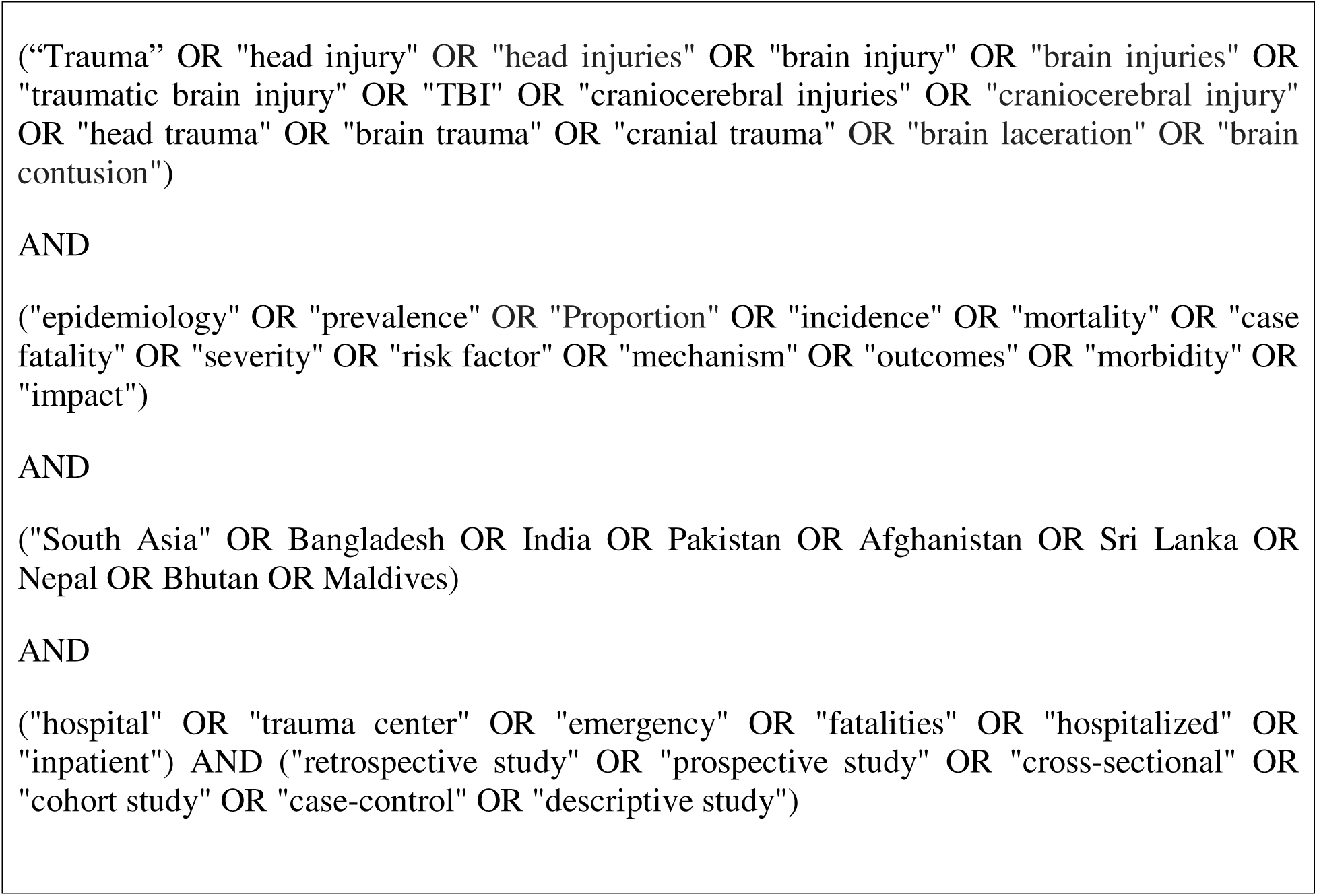

### Appendix 2: Methodological Evaluation of Observational Research (MORE) Checklist for Quality assessment of Included Studies

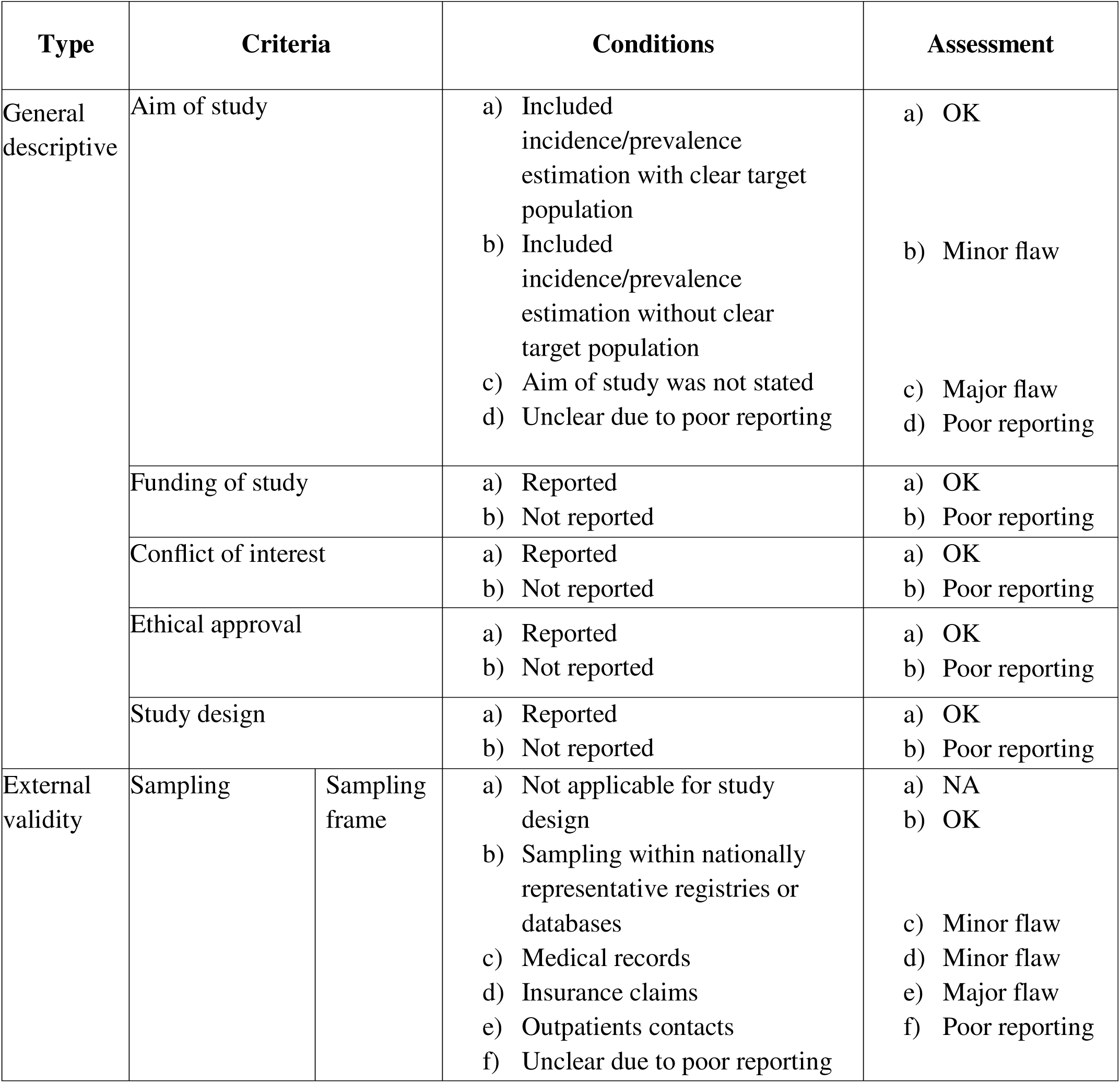
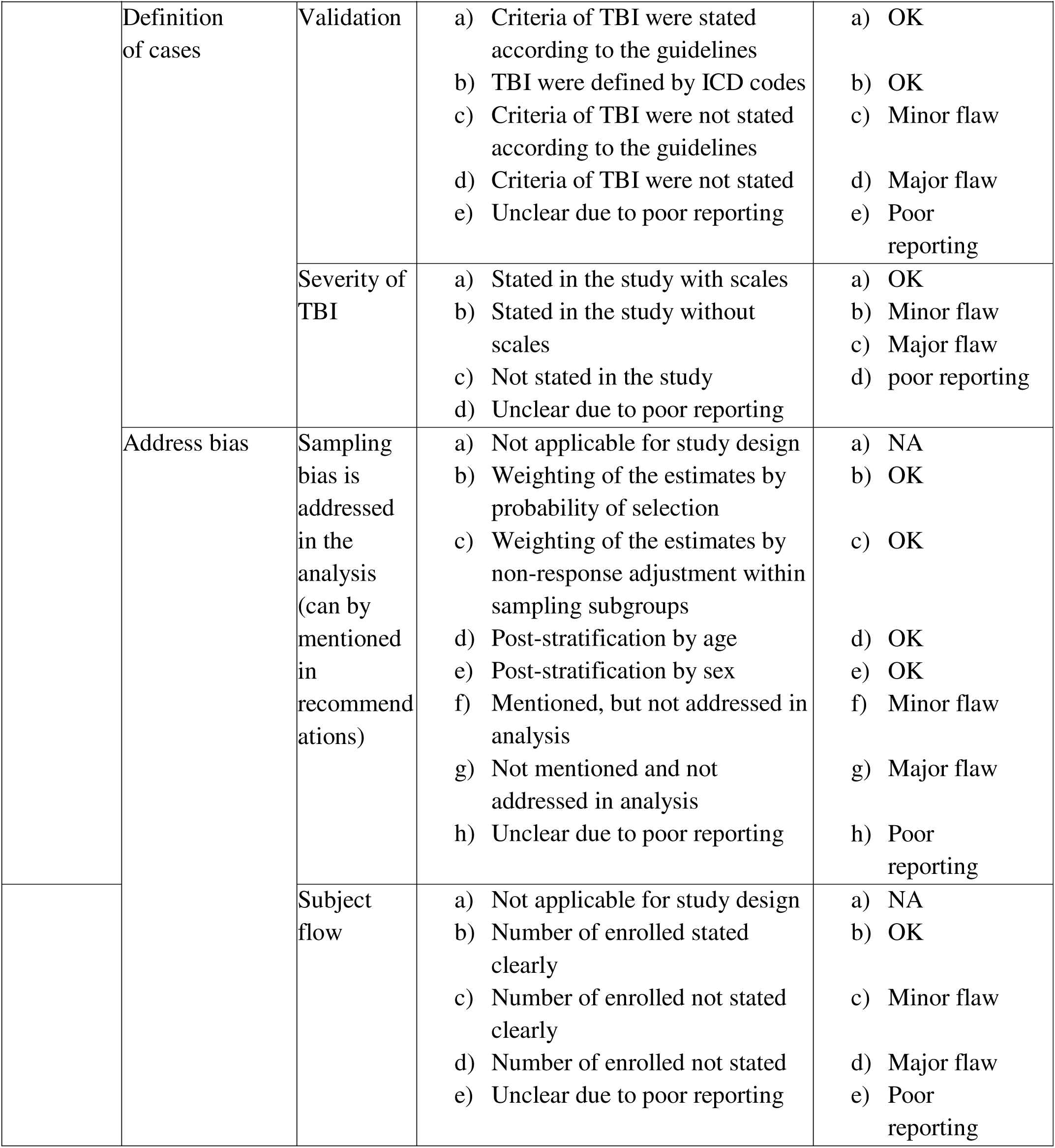
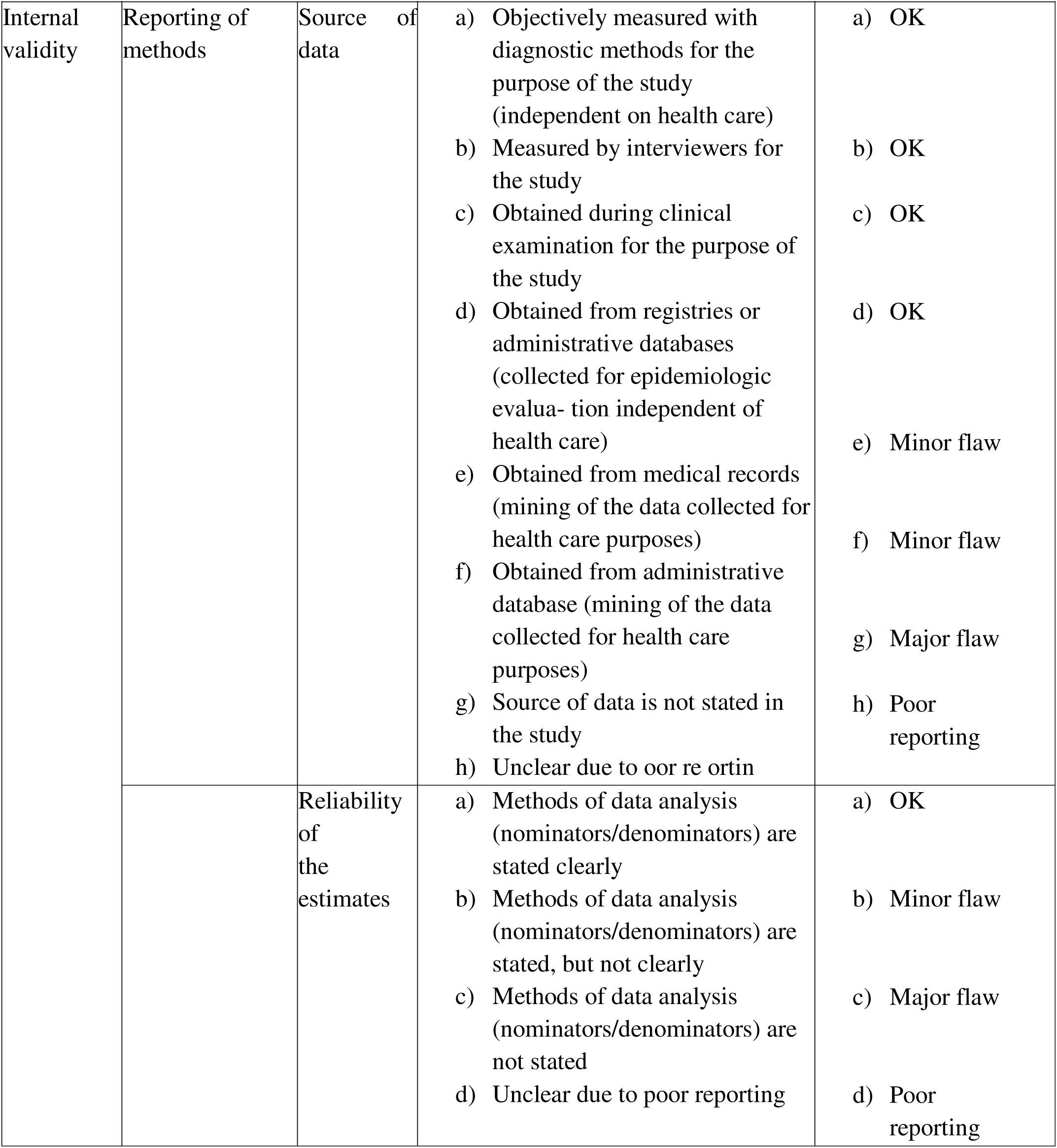
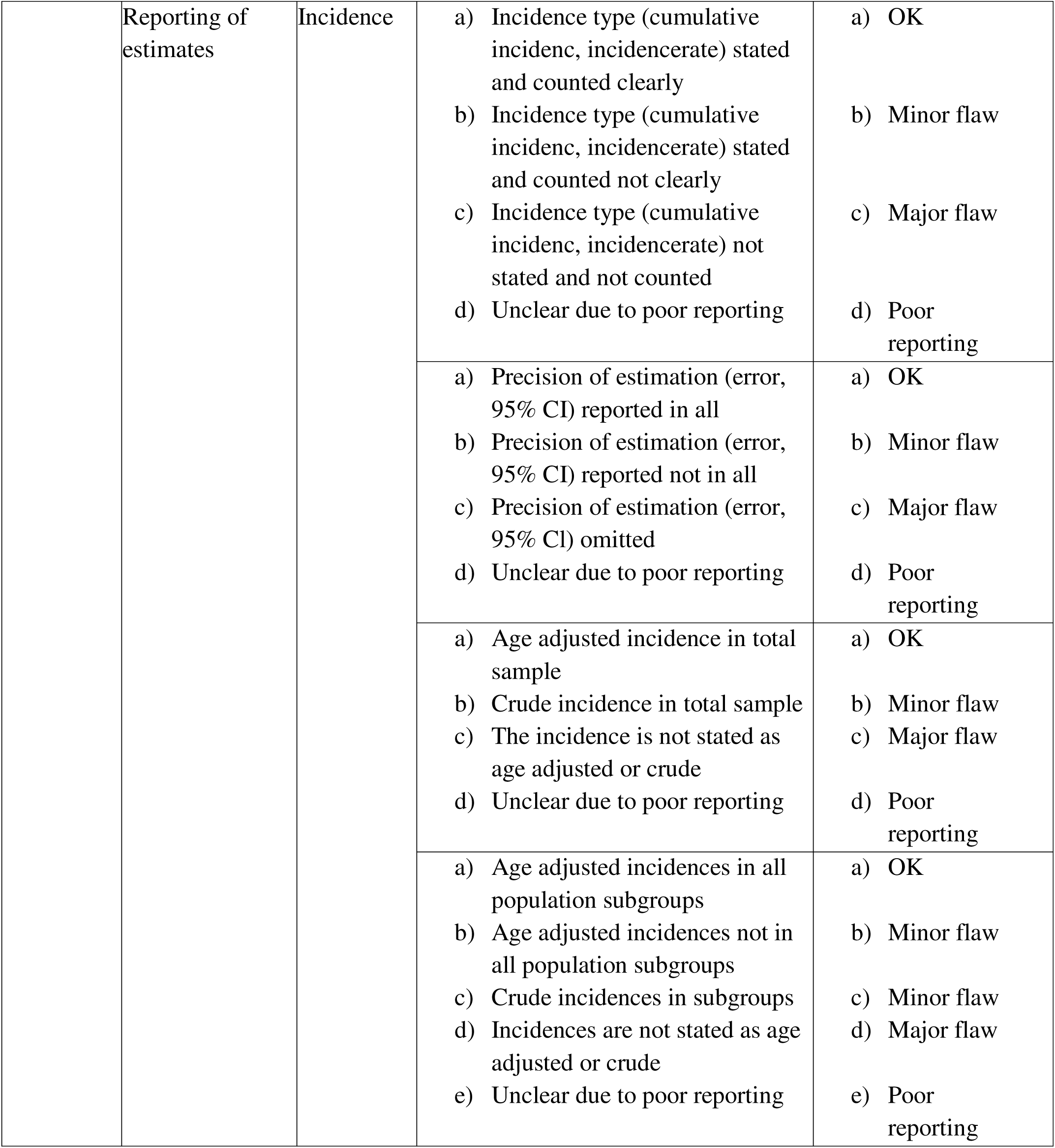
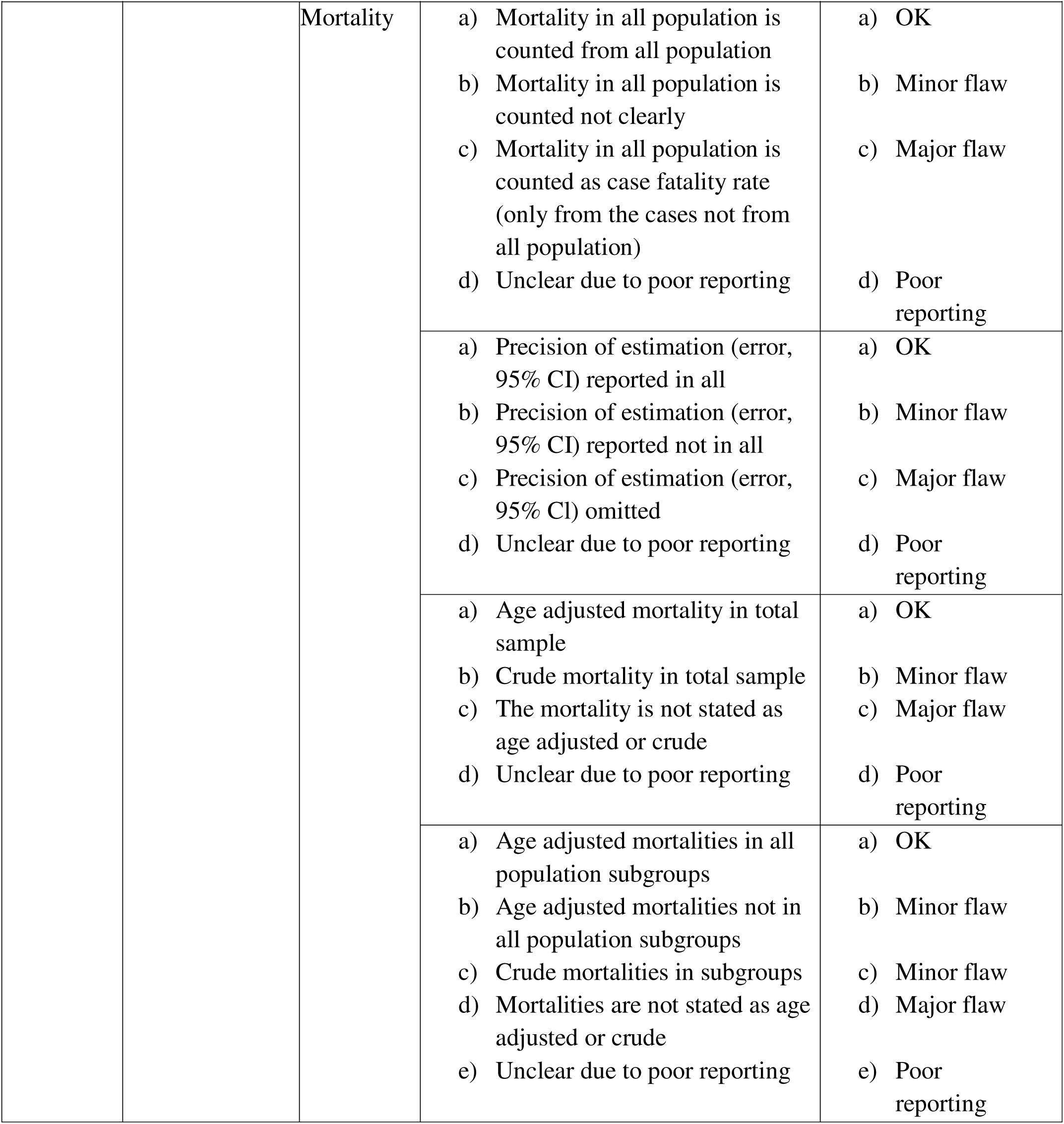

## References

1. Balakin E, Yurku K, Fomina T, et al. A Systematic Review of Traumatic Brain Injury in Modern Rodent Models: Current Status and Future Prospects. Biology. 2024;13(10):813. 10.3390/biology13100813

2. Turner GM, McMullan C, Aiyegbusi OL, et al. Stroke risk following traumatic brain injury: Systematic review and meta-analysis. Int J Stroke. 2021;16(4):370–84. 10.1177/17474930211004277

3. Semple BD, Zamani A, Rayner G, Shultz SR, Jones NC. Affective, neurocognitive and psychosocial disorders associated with traumatic brain injury and post-traumatic epilepsy. Neurobiol Dis. 2019;123:27–41. 10.1016/j.nbd.2018.07.018

4. Fu TS, Jing R, McFaull SR, Cusimano MD. Health & Economic Burden of Traumatic Brain Injury in the Emergency Department. Can J Neurol Sci. 2016;43(2):238–47. 10.1017/cjn.2015.320

5. Sheriff FG, Hinson HE. Pathophysiology and Clinical Management of Moderate and Severe Traumatic Brain Injury in the ICU. Semin Neurol. 2015;35(1):42–9. 10.1055/s-0035-1544238

6. Retel Helmrich IRA, Lingsma HF, Turgeon AF, Yamal JM, Steyerberg EW. Prognostic Research in Traumatic Brain Injury: Markers, Modeling, and Methodological Principles. J Neurotrauma. 2021;38(18):2502–13. 10.1089/neu.2019.6708

7. Lee B, Newberg A. Neuroimaging in Traumatic Brain Imaging. NeuroRx. 2005;2(2):372–83. 10.1602/neurorx.2.2.372

8. Reyes J, Spitz G, Major BP, et al. Utility of Acute and Subacute Blood Biomarkers to Assist Diagnosis in CT-Negative Isolated Mild Traumatic Brain Injury. Neurology. 2023;101(20):e1992–2004. 10.1212/WNL.0000000000207881

9. Huang XF, Ma SF, Jiang XH, et al. Causes and global, regional, and national burdens of traumatic brain injury from 1990 to 2019. Chinese Journal of Traumatology. 2024;27(6):311–22. 10.1016/j.cjtee.2024.03.007

10. Allen BC, Cummer E, Sarma AK. Traumatic Brain Injury in Select Low- and Middle-Income Countries: A Narrative Review of the Literature. Journal of Neurotrauma. 2023;40(7– 8):602–19. 10.1089/neu.2022.0068

11. Liang KWH, Lee JH, Qadri SK, et al. Differences in clinical outcomes and resource utilization in pediatric traumatic brain injury between countries of different sociodemographic indices. J Neurosurg Pediatr. 2024;33(5):461–8.

12. Kumar S, Jha AK, Kumari J, Paul SK. Road traffic accidents are a major cause of traumatic brain injury in north bihar – a prospective study. ijsr. 2024;13(03):76–8. 10.36106/ijsr

13. Newall N, Gajuryal S, Bidari S, et al. Epidemiology and Pattern of Traumatic Brain Injuries at Annapurna Neurological Institute & Allied Sciences, Kathmandu, Nepal. World Neurosurgery. 2020;141:413–20. 10.1016/j.wneu.2020.04.250

14. Rahman FN, Das S, Kader M, Mashreky SR. Epidemiology, outcomes, and risk factors of traumatic brain injury in Bangladesh: a prospective cohort study with a focus on road traffic injury-related vulnerability. Front Public Health. 2025;13:1514011. 10.3389/fpubh.2025.1514011

15. Dewan MC, Rattani A, Gupta S, et al. Estimating the global incidence of traumatic brain injury. Journal of Neurosurgery. 2019;130(4):1080–97. 10.3171/2017.10.JNS17352

16. Naik A, Bederson MM, Detchou D, et al. Traumatic Brain Injury Mortality and Correlates in Low- and Middle-Income Countries: A Meta-Epidemiological Study. Neurosurgery. 2023;93(4):736–44. 10.1227/neu.0000000000002479

17. Brazinova A, Rehorcikova V, Taylor MS, et al. Epidemiology of Traumatic Brain Injury in Europe: A Living Systematic Review. Journal of Neurotrauma. 2021;38(10):1411–40. 10.1089/neu.2015.4126

18. Peeters W, Van Den Brande R, Polinder S, et al. Epidemiology of traumatic brain injury in Europe. Acta Neurochir. 2015;157(10):1683–96. 10.1007/s00701-015-2512-7

19. Bandyopadhyay S, Kawka M, Marks K, et al. 244 Traumatic Brain Injury Related Paediatric Mortality and Morbidity in Low- And Middle-Income Countries: A Systematic Review and Meta-Analysis. British Journal of Surgery. 2021;11;108. 10.1093/bjs/znab258.002

20. Maas AIR, Harrison-Felix CL, Menon D, et al. Standardizing Data Collection in Traumatic Brain Injury. Journal of Neurotrauma. 2011;28(2):177–87. 10.1089/neu.2010.1617

21. Roozenbeek B, Maas AIR, Menon DK. Changing patterns in the epidemiology of traumatic brain injury. Nat Rev Neurol. 2013 Apr;9(4):231–6. 10.1038/nrneurol.2013.22

22. Maas AIR, Menon DK, Steyerberg EW, et al. Collaborative European NeuroTrauma Effectiveness Research in Traumatic Brain Injury (CENTER-TBI): A Prospective Longitudinal Observational Study. Neurosurgery. 2015;76(1):67–80. 10.1227/NEU.0000000000000575

23. Jeffcote T, Battistuzzo CR, Plummer MP, et al. PRECISION-TBI: a study protocol for a vanguard prospective cohort study to enhance understanding and management of moderate to severe traumatic brain injury in Australia. BMJ Open. 2024;14(2):e080614. 10.1136/bmjopen-2023-080614

24. Korley FK, Jain S, Sun X, et al. Prognostic value of day-of-injury plasma GFAP and UCH-L1 concentrations for predicting functional recovery after traumatic brain injury in patients from the US TRACK-TBI cohort: an observational cohort study. The Lancet Neurology. 2022;21(9):803–13. 10.1016/S1474-4422(22)00256-3

25. Schwenkreis P, Gonschorek A, Berg F, et al. Prospective observational cohort study on epidemiology, treatment and outcome of patients with traumatic brain injury (TBI) in German BG hospitals. BMJ Open. 2021;11(6):e045771. 10.1136/bmjopen-2020-045771

26. McCrea MA, Giacino JT, Barber J, et al. Functional Outcomes Over the First Year After Moderate to Severe Traumatic Brain Injury in the Prospective, Longitudinal TRACK-TBI Study. JAMA Neurol. 2021;78(8):982. 10.1001/jamaneurol.2021.2043

27. Menon DK, Schwab K, Wright DW, Maas AI. Position Statement: Definition of Traumatic Brain Injury. Archives of Physical Medicine and Rehabilitation. 2010;91(11):1637–40. 10.1016/j.apmr.2010.05.017

28. H. K, Paparajamurthy P, K. M, K. A. A clinical analysis of outcome in management of head injury in patients with highway road accidents. Int J Res Med Sci. 2016;2079–83. 10.18203/2320-6012.ijrms20161764

29. Mohanta T, Roy S, Chatterjee S. A clinico-epidemiological study of fatal head injuries in an autopsy centre of Kolkata (West Bengal). IOSR Journal of Dental and Medical Sciences. 2019;18(6).10.9790/0853-1806082528

30. Anwar DMM. A Comprehensive Study and Creation of Profile of Trauma Patients Admitted in ICU of a Tertiary Care Hospital in India. jmscr. 2016;04(04). 10.18535/jmscr/v4i4.08

31. Ahmed A, Mustahsan SM, Tariq F, Abidi SMA, Aslam MO. A Cross-Sectional Study: Head Injury in Children of Karachi. Int j endorsing health sci res. 2015;3(2):18. 10.29052/IJEHSR.v3.i2.2015.18-20

32. Sivakumar R, Subrahmanyam BV, Phanindra SV, Munivenkatappa A, Kumar SS, Agrawal A. A descriptive study of cranio-cerebral injuries admitted in tertiary care center of coastal Andhra Pradesh. Romanian Neurosurgery. 2018;32(2):384–90. 10.2478/romneu-2018-0048

33. Takalkar Y, Vashist K, Chakravarthy V, Shinde P. A prospective study of head injury patterns in motorcycle riders wearing/not wearing helmets. Pol Przegl Chir. 2022;95(1):25–8. 10.5604/01.3001.0015.9282

34. Basheer N, Varghese JC, Kuruvilla R, Alappat JP, Mathew J. A Prospective Study on the Incidence and Outcome of Cranial Nerve Injuries in Patients with Traumatic Brain Injuries. Indian Journal of Neurotrauma. 2021;18(01):45–50. 10.1055/s-0041-1724141

35. Vinayak R, Vaishali R, Akshay P, Mayank V, Abhishek M, Ritvij P, et al. A Study of Paediatric Head Injuries and Its Outcome. International Journal of Health Sciences. 2014;(12).

36. Chakravarthi H, Murthy P, P N V, M N C. A study of traumatic brain injuries at a tertiary care hospital in karnataka, South India. jemds. 2014;3(14):3750–7. 10.14260/jemds/2014/2355

37. Agrawal A, Munivenkatappa A, Subrahmanyam BV, Satish Kumar S, Ramamohan P. Admission characteristics and outcome in traumatic brain injury patients: a preliminary report from a tertiary care hospital. Romanian Neurosurgery. 2016;30(2):252–7. 10.1515/romneu-2016-0039

38. Mahore A, Dange N, Vutha R, Garale M. Agonies and nuances of geriatric head injury: experience of a tertiary care institute of Western India. Int Surg J. 2017;4(7):2205. 10.18203/2349-2902.isj20172767

39. Shobhana SS, RaviRaj KG, Abhishek Y, Kumar RL. An Analysis of Pattern of Fatal Head Injuries in Road Traffic Accidents. Medico-Legal Update. 2019;19(1):130. 10.37506/mlu.v19i1.898

40. Gunaratne A, Wadanambi S. An Audit on Admission of Patients with Head Injury to Teaching Hospital Karapitiya, Galle. Sri Lankan J Anaesthesiol. 2009;17(2):51. 10.4038/slja.v17i2.1297

41. Gambhir Singh O. An epidemiological study of fatal head injury cases in a sub urban region of Chennai: A prospective study of 350 autopsy cases. SAJHP. 2022;4(1):10–4. 10.18231/j.sajhp.2021.003

42. Shekhar C, Gupta L, Premsagar I, Sinha M, Kishore J. An epidemiological study of traumatic brain injury cases in a trauma centre of New Delhi (India). J Emerg Trauma Shock. 2015;8(3):131. 10.4103/0974-2700.160700

43. Kumaran KM, Raja P, Jasmine M. An Epidemiological Study of Traumatic Head Injury in a Tertiary Care Center in Kancheepuram, Tamil Nadu, India. Asian Journal of Medicine and Health. 2019;1–5. 10.9734/ajmah/2019/v16i130136

44. Khan S, Nawaz S, Hayat F, Rehman S, Sardar N. Analysis of Head Injuries Due to Motorcycle Accidents Attended in Medical Teaching … 182 Institute (MTI), DHQ, Gomal Medical College (GMC), Dera Ismail Khan, Pakistan. Pak J Neurol Surg. 2019;23(3):182–7. 10.36552/pjns.v23i3.357

45. Rajasekhar DrV, Dinesh DrG. Autopsy findings study among rta cases with traumatic brain injuries. ijsr. 2019;8(11). 10.36106/ijsr

46. Bhat I, Malik N, Kareem K, et al. Burden of Moderate and Severe Head Injury in Kashmir Valley. Indian Journal of Neurotrauma. 2021;18(01):79–83. 10.1055/s-0040-1717217

47. Naz A, Rasheed G, Baig MS, Baqi S. The Characteristics and Outcome of Patients with Traumatic Brain Injury in the Intensive Care Unit of a Public Sector Hospital in Karachi, Pakistan. J Dow Univ Health Sci. 2021;15(3). 10.36570/jduhs.2021.3.1187

48. Adhikari K, Gupta MK, Pant AR, Rauniyar RK. Clinical Patterns and Computed Tomography Findings in Patients with Cranio-Cerebral Trauma in Tertiary Hospital in Eastern Nepal. Journal of Nepal Health Research Council. 2019;17(01):56–60. 10.33314/jnhrc.v17i01.1269

49. Narwade N, Narwade P, Ghosalkar M, et al. Clinical profile and management of head injury at tertiary health care center in rural area, India. Int J Res Med Sci. 2015;3137–40. 10.18203/2320-6012.ijrms20151151

50. Roy JM, Balasubramaniam S, Barve PS, Nadkarni TD. Clinical Profile, Evaluation of Imaging Guidelines, and Management of Pediatric Traumatic Brain Injury at a Tertiary Care Center in India: A Review of 269 Patients. J Pediatr Neurosci. 2023;18(3):196–202. 10.4103/jpn.JPN_30_22

51. Lodha KG, Gupta TK, Jaiswal G, Singh Y. Clinical spectrum of paediatric head injury. A prospective study from tribal region. roneuro. 2020;158–63. 10.33962/roneuro-2020-025

52. Madaan P, Agrawal D, Gupta D, et al. Clinicoepidemiologic Profile of Pediatric Traumatic Brain Injury: Experience of a Tertiary Care Hospital From Northern India. J Child Neurol. 2020;35(14):970–4. 10.1177/0883073820944040

53. Islam MJ, Islam MS, Haseen F, et al. Clinico-Epidemiological Study in Admitted Patients with Traumatic Brain Injury (TBI) in two selected Tertiary Care Centers in Dhaka City. J Natl Inst Neurosci Bangladesh. 2023;8(2):105–11. 10.3329/jninb.v8i2.63693

54. Pal SS, Shukla A, Jain A. Clinicopathological study of paediatric head injury in gandhi medical college bhopal from may 2011 to june 2013. jemds. 2015;04(07):1197–212. 10.14260/jemds/2015/167

55. Dara PK, Parakh M, Choudhary S, Jangid H, Kumari P, Khichar S. Clinico-radiologic Profile of Pediatric Traumatic Brain Injury in Western Rajasthan. Journal of Neurosciences in Rural Practice. 2018;09(02):226–31. 10.4103/jnrp.jnrp_269_17

56. Patil S, Huda T, Jain SC, Pandya B, Narang R. Comparative study of pattern of head injury in a rural community hospital and a tertiary care hospital. Int Surg J. 2019;6(12):4272. 10.18203/2349-2902.isj20195386

57. Jooma R, Ahmed S, Zarden AM. Comparison of Two Surveys of Head Injured Patients presenting during a Calendar Year to an Urban Medical Centre 32 Years Apart. J Pak Med Assoc.2005;55(12):530–2.

58. Rizwan MH, Saddiqa A, Khan M, Khan MAS, Mughal SBK, Pervaiz T. Computed Tomography Scan Head Findings in Patients With Various Glasgow Coma Scales Presenting with Head Injury in Emergency of a Tertiary Care Hospital. PAFMJ. 2023;73(3):888–91. 10.51253/pafmj.v73i3.9401

59. Mahakul D, Doddamani RS, Meena RK, Sawarkar D, Sharma R, Kedia S, et al. Country-Made Firearm (Desi-Katta)-Related Cranial Injuries: An Institutional Experience. Neurology India. 2022;70(5):1976–81. 10.4103/0028-3886.359174

60. Gouda HS, Meghana PR, Prabha BB. Cranio-Cerebral Injuries in Victims of Fatal Road Traffic Accident: A 5 year Post-Mortem Study. International Journal of Medical Toxicology and Forensic Medicine. 2014;4(3(Summer)):77–82. 10.22037/ijmtfm.v4i3(Summer).5888

61. Rashid B, Wani M, Kirmani A, Raina T, Raina S. Craniocerebral missile injuries in civilian Kashmir – India. Afr J Neuro Sci. 2010;29(2):13–28. 10.4314/ajns.v29i2.70402

62. Sharma S, Deshwal Y. Demographic and clinical profile of head injury in Western Uttar Pradesh-A study at SVBP hospital Meerut. IJN. 2020;6(2):116–23. 10.18231/j.ijn.2020.024

63. Thapa S, Baral A, Bajracharya K, Sharma MR. Demographic and Clinical Profiles of Patients with Traumatic Brain Injury Managed in a District Hospital in Nepal. Kathmandu Univ Med J. 2023;84(4):399–403.

64. Kirankumar M, Satri V, Satyanarayana V, Ramesh Chandra V, Madhusudan M, Sowjanya J. Demographic profile, clinical features, imaging and outcomes in patients with traumatic brain injury presenting to emergency room. J Clin Sci Res. 2019;8(3):132. 10.4103/JCSR.JCSR_65_19

65. Bhole AM, Potode R, Agrawal A, Joharapurkar SR. Demographic profile, clinical presentation, management options in cranio-cerebral trauma: an experience of a rural hospital in Central India. PJMC. 2007; 23 (5): 724–727

66. Saxena M, Rai H, Maheshwari O. Demographics, injury characteristics and outcome of traumatic brain injuries at a general surgical unit. The Indian Journal of Neurotrauma. 2008;5(1):39–40. 10.1016/S0973-0508(08)80027-5

67. Kumar D, Bains V, Sharma BR, Harish D. Descriptive Study of Head Injury and its Associated Factors at Tertiary Hospital, Northern India. Journal of Community Medicine & Health Education. 2012;2(4):1–3. 10.4172/2161-0711.1000141

68. Khanal B, Kafle P, Singh SK, Yadav SK, Neupane B, Yadav DK. Early Outcome of Surgery in Pediatric Head Injury: Experience From a Tertiary Care Center in Eastern Nepal. Journal of Institute of Medicine Nepal. 2020;42(2). 10.3126/jiom.v42i2.37529

69. Choudhary A, Kumar A, Sharma RK, et al. Emergency Department Management of Mild Traumatic Brain Injury in New Delhi–A Single Institute Cohort Management Data. Indian Journal of Neurosurgery. 2022;11(02):123–7. 10.1055/s-0040-1719236

70. Yaqoob U, Javeed F, Rehman L, Pahwani M, Madni S, Uddin MM. Emergency Department Outcome of Patients with Traumatic Brain Injury – A Retrospective Study from Pakistan. Pak J Neurol Surg. 2021;25(2):237–44. 10.36552/pjns.v25i2.540

71. Arfat Mohd, Verma AP, Shukla A, Mittal KK. Emerging trends of traumatic brain injury in Western Uttar Pradesh, India: diagnosis and rehabilitation. Int J Res Med Sci. 2019;7(6):2356. 10.18203/2320-6012.ijrms20192527

72. Mangal G. Epidemiological Analysis and Clinical Characteristics of Traumatic Brain Injury in Southern Rajasthan: A Hospital Based Study. jmscr. 2017;05(04):21115–20. 10.18535/jmscr/v5i4.218

73. Sharma M, Pandey S, Kumar P, Singh K, Kumar P, Jha R. Epidemiological and clinico-radiological evaluation of head injury in pediatric population. J Pediatr Neurosci. 2020;15(4):386. 10.4103/JPN.JPN_44_19

74. Goel A, Kumar S, Bagga MK. Epidemiological and Trauma Injury and Severity Score (TRISS) analysis of trauma patients at a tertiary care centre in India. The National Medical Journal Of India. 2004;17(4).

75. Pateria S, Kamath R, Palimar V, L.K H. Epidemiological Pattern of Head Injuries in Road Traffic Accident Victims Presenting to a Tertiary Care Hospital in South India. IJFMT. 2020. 10.37506/ijfmt.v14i4.12345

76. Jha S, Yadav B, Karn A, Aggrawal A, Gautam A. Epidemiological Study of Fatal Head Injury in Road Traffic Accident Cases: A Study from BPKIHS, Dharan. Health Renaissance. 2010;8(2):97–101. 10.3126/hren.v8i2.4420

77. Rao BH, Satyavaraprasad K, Rajiv PK, Kumar PV, Phaneeswar T. Epidemiological Study of Head Injuries in Andhra Medical College, King George Hospital, Visakhapatnam. Ijss. 2015;3(9). 10.17354/ijss/2015/562

78. Chakraborty PN, Sarkar SC. Epidemiological Study of Patterns of Head Injuries in Fatal Road Traffic Accidents in Tripura. Indian Jour of Foren Med & Toxicol. 2017;11(1):89. 10.5958/0973-9130.2017.00019.6

79. Agrawal A, Galwankar S, Kapil V, et al. Epidemiology and clinical characteristics of traumatic brain injuries in a rural setting in Maharashtra, India. 2007-2009. *Int J Crit Illn Inj Sci.* 2012;2(3):167. 10.4103/2229-5151.100915

80. Avtar Malav R, Shankar Shukla U, Nagar M. Epidemiology and clinical characteristics of traumatic head injuries in central part of India, 2018-2019. IJN. 2019;5(3):117–21. 10.18231/j.ijn.2019.017

81. Agrawal A, Agrawal C, Kumar A, et al. Epidemiology and management of paediatric head injury in eastern Nepal. Afr J Paediatr Surg. 2008;5(1):15. 10.4103/0189-6725.41630

82. Das S, Chaurasia B, Ghosh D, Sarker AC. Epidemiology and Treatment Outcomes of Head Injury in Bangladesh: Perspective from the Largest Tertiary Care Hospital. Indian Journal of Neurotrauma. 2023;20(01):011–7. 10.1055/s-0040-1718780

83. Kamal VK, Agrawal D, Pandey RM. Epidemiology, clinical characteristics and outcomes of traumatic brain injury: Evidences from integrated level 1 trauma center in India. Journal of Neurosciences in Rural Practice. 2016;7(04):515–525. 10.4103/0976-3147.188637

84. Shanmugaiah A, Jayakumar J. Epidemiology, treatment strategy & outcome in patients with traumatic basifrontal contusion-a retrospective study in a tertiary care hospital in south india. ijsr. 2023;8–10. 10.36106/ijsr/1006662

85. Hassan N, Ali M, Haq NU, et al. Etiology, clinical presentation and outcome of traumatic brain injury patients presenting to a teaching hospital of Khyber Pakhtunkhwa. Journal of Postgraduate Medical Institute. 2017;31(4).

86. Anand KV, Shahid PT, Shameel KK. Evaluating GCS and FOUR Score in Predicting Mortality of Traumatic Brain Injury Patients (TBI): A Prospective Study in a Tertiary Hospital of South Malabar. Journal of Pharmacy and Bioallied Sciences. 2024;16(Suppl 1):S598–600. 10.4103/jpbs.jpbs_884_23

87. Bhaumik DU. Evaluation of Incidence, Mode of Injury and Clinical Outcomes of Traumatic Brain Injury in Tertiary Health Center in Southern Rajasthan. jmscr. 2018;6(8). 10.18535/jmscr/v6i8.20

88. Anand A, Verma S, Garg P, Noori MT, Kajal A, Verma A. Evaluation of pattern and prognostic factors of head injury cases in a tertiary care centre. Int Surg J. 2020;7(5):1535. 10.18203/2349-2902.isj20201865

89. Singh A, Meena R, Sharma V, Kumar N, Meena L. Evaluation of the incidence, nature of injury and clinical outcomes of head injuries in North Eastern part of India. Int J Med Sci Public Health. 2013;2(2):287. 10.5455/ijmsph.2013.2.285-287

90. Khan SI, Karim R, Khan SI, et al. Factors affecting severity and prognosis of traumatic brain injury among Bangladeshi patients: An institution based cross-sectional study. Traffic Injury Prevention. 2024;25(8):1072–80. 10.1080/15389588.2024.2363470

91. Yattoo GH, Tabish SA, Afzal WM, Kirmani A. Factors influencing outcome of head injury patients at a tertiary care teaching hospital in India. International journal of health sciences. 2009;3(1):59.

92. Singh OG. Fatal craniocerebral injuries in victims who survived for some period. IAIM. 2014;1(1).

93. Henriksson T, Kjellberg J, Shakya Y, Kurlberg G. Head Injuries at the Emergency Department of a University Hospital in Kathmandu. J Inst Med Nepal. 2020;42(3):47–51. 10.59779/jiomnepal.1133

94. Sultana P, Jabin N, Mainuddin KM, Khan RH, Reza AS. Head Injuries in Fatal Road Traffic Accidents in Northern Districts of Bangladesh. Journal of Shaheed Suhrawardy Medical College. 2021;13(1):75–9. 10.3329/jssmc.v13i1.60936

95. Agrawal A, Agrawal CS, Kumar A, Lewis O, Malla G, Chalise P. Head injury at a tertiary referral centre in the eastern region of Nepal. East and central African journal of surgery. 2009;14(1):57–63.

96. Kafle P, Khanal B, Yadav DK, Poudel D, Karki T, Cherian I. Head Injury in Nepal: An Institutional Based Prospective Study on Clinical Profile, Management and Early Outcome of Traumatic Brain Injury in Eastern Part of Nepal. Birat J Health Sci. 2019;4(2):750–4. 10.3126/bjhs.v4i2.25459

97. Malik Y, Chaliha RR. Head Injury in Road Traffic Accidents - A Study from North East India. IJCMR. 2019;6(10). 10.21276/ijcmr.2019.6.10.29

98. Khan M, Yaqoob U, Hassan Z, Uddin MM. Immediate Outcomes of Traumatic Brain Injury at a Tertiary Care Hospital Of Pakistan-A Retrospective Stud. Research Square. 2020. 10.21203/rs.3.rs-84330/v1

99. Bansal V, Patil P, Faria I, et al. Mortality and Risk Factors in Isolated Traumatic Brain Injury Patients: A Prospective Cohort Study. Journal of Surgical Research. 2022;279:480–90. 10.1016/j.jss.2022.05.005

100. Kumar A, Hedaoo K, Bajaj J, et al. Neurotrauma Audit at Netaji Subhash Chandra Bose Medical College, Jabalpur, Madhya Pradesh. IJNT. 2019;16(02/03):109–12. 10.1055/s-0039-3402931

101. Raja IA, Vohra AH, Ahmed M, Mb B. Neurotrauma in Pakistan. World journal of surgery. 2001;25(9):1230. 10.1007/s00268-001-0087-3

102. Khan MK, Hanif SA, Husain M, Huda MF, Sabri I. Pattern of Non-Fatal Head Injury in Adult Cases Reported at J.N.M.C. Hospital, A.M U, Aligarh. Journal of Indian Academy of Forensic Medicine. 2011;33(1). 10.1177/0971097320110107

103. Ahmed S, Khan S, Agrawal D, Sharma B. Out come in head injured patients: Experience at a level 1 trauma centre. The Indian Journal of Neurotrauma. 2009;6(2):119–21. 10.1016/S0973-0508(09)80005-1

104. Sharma S, Bansal H, Singh J, Chaudhary A. Outcome and its predictors in traumatic brain injury in elderly population: Institutional study from Northern India. Journal of Family Medicine and Primary Care. 2021;10(1):289–94. 10.4103/jfmpc.jfmpc_1559_20

105. Agrawal D, Ahmed S, Khan S, Gupta D, Sinha S, Satyarthee G. Outcome in 2068 patients of head injury: Experience at a level 1 trauma centre in India. Asian J Neurosurg. 2016;11(02):143–5.

106. Ahmed S, Rehman L, Afzal A, Javeed F. Outcome of head injury in motorbike riders. Pak J Med Sci. 2023;39(2). 10.12669/pjms.39.2.6371

107. Nath HD, Tandon V, Mahapatra AK, Siddiqui SA, Gupta DK. Outcome of head injury in unknown patients at Level-1 apex trauma centre. The Indian Journal of Neurotrauma. 2011;8(1):11–5. 10.1016/S0973-0508(11)80018-3

108. Khan J, Khan A, Naseem A, Farrukh A. Outcome of Patients Admitted with Head Injury in Intensive Care Unit (ICU) of a Tertiary Care Hospital of Rawalpindi. Pakistan Armed Forces Medical Journal. 2023;73. https://d0i.org/10.51253/pafmj.v73i3.3827

109. Minai FN, Kumar R, Shafiq F, Kumar D. Outcome Of Traumatic Brain Injury: A Retrospective Audit. The Internet Journal of Anesthesiology.2013;32 (2)

110. Banzal RK, Jaiin A, Yadav J, Dubey BP. Pattern and Distribution of Head Injuries in Fatal Road Traffic Accidents in Bhopal Region of Central India. Jou Indian Acad of Foren Med. 2015;37(3):242. 10.5958/0974-0848.2015.00061.5

111. Soni SK, Dadu SK, Singh BK, Pandey D. Pattern and distribution of head injuries in fatal road traffic accidents in Indore region of central india. Sch J App Med Sci. 2016;4(5D):1711–6.

112. Fernandes TB, Mandrekar PN, Visen A, Sinai Khandeparker PV, Dhupar V, Akkara F. Pattern of associated brain injury in maxillofacial trauma: a retrospective study from a high-volume centre. British Journal of Oral and Maxillofacial Surgery. 2022;60(10):1373–8. 10.1016/j.bjoms.2022.09.002

113. Patil AM, Vaz WF. Pattern of Fatal Blunt Head Injury: A Two Year Retrospective / Prospective Medico Legal Autopsy Study. J Indian Acad Forensic Med. 2010; 32(2). 10.1177/0971097320100215

114. Menon A, Pai VK, Rajeev A. Pattern of fatal head injuries due to vehicular accidents in Mangalore. Journal of Forensic and Legal Medicine. 2008;15(2):75–7. 10.1016/j.jflm.2007.06.001

115. Punia RK, Verma LC, Pathak D. Pattern of Fatal Head Injuries in Road Traffic Accidents at SMS Hospital, Jaipur - An Autopsy Based Study. Medico-Legal Update. 2014;14(1):30. 10.5958/j.0974-1283.14.1.007

116. Urfi D, Amir A, Huda F, Khalil S, Kirmani S. Pattern of head injuries among victims of road traffic accidents in a tertiary care teaching hospital. Indian Journal of Community Health. 2013;25(2):126–33.

117. R K C, Mishra A, Chaturvedi P. PATTERN OF HEAD INJURIES IN FATAL ROAD TRAFFIC ACCIDENTS IN INDORE REGION, M. P. jemds. 2014;3(21):5645–51. 10.14260/jemds/2014/2642

118. Sah SK, Neupane BR, Atreya A, Karki N. Pattern of Head Injuries in Western Hilly Region of Nepal: A Hospital-based Cross-sectional Study. J Lumbini Med Coll. 2020;8(1):65–70. 10.22502/jlmc.v8i1.297

119. Pate RS, Hire RC, Rojekar MV. Pattern of head injury in central India population. Int J Res Med Sci. 2017;5(8):3515. 10.18203/2320-6012.ijrms20173553

120. Ahmed S, Jalal MLA, Khan SS, Akhter I, Rizwan M, Chughtai WN. Pattern, Treatment and Outcomes of Children with Head Injuries: A 3 Year experience from a TC Hospital of South Punjab, Pakistan. PJMHS. 2022;16(4):247–9. 10.53350/pjmhs22164247

121. Islam MS, Rahman MF, Islam MA. Patterns and Outcome of Traumatic Brain Injury Patients: A Study in a Tertiary Level Military Hospital. J Armed Forces Med Coll. 2020;15(1):75–8. 10.3329/jafmc.v15i1.48649

122. Arora S, Khajuria B. Patterns of Cranio Cerebral Injuries in Fatal Vehicular Accidents in Jammu Region-J&K State. JK Science. 2016;18(3).

123. Hemalatha N, Singh OG. Patterns of Cranio-intracranial injuries In Fatal Head Injury Cases. Journal of Indian Academy of Forensic Medicine. 2013;35(2):106–8. 10.1177/0971097320130205

124. Sharma B, Harish D, Singh G, Vij K. Patterns of Fatal Head Injury in Road Traffic Accidents. Bahrain Medical Bulletin. 2003;25(1)

125. Tandle RM, Keoliya AN. Patterns of head injuries in fatal road traffic accidents in a rural district of Maharashtra-Autopsy based study. Journal of Indian Academy of Forensic Medicine. 2011;33(3):228–31. 10.1177/0971097320110312

126. Karmacharya BG, Acharya B. Pediatric Head Injuries in a Neurosurgery Center of Nepal: An Epidemiological Perspective. American Journal of Public Health Research. 2015;3(4A):76–9. 10.12691/ajphr-3-4A-16

127. Mukhida K, Sharma MR, Shilpakar SK. Pediatric neurotrauma in Kathmandu, Nepal: implications for injury management and control. Childs Nerv Syst. 2006;22(4):352–62. 10.1007/s00381-005-1235-0

128. A. Ansari S, Panezai AM. Penetrating craniocerebral injuries: an escalating problem in Pakistan. British Journal of Neurosurgery. 1998;12(4):340–3. 10.1080/02688699844853

129. Ahmad M, Rahman FN, Chowdhury MH, Islam A, Hakim MA. Postmortem Study of Head Injury in Fatal Road Traffic Accidents. *Journal of Armed Forces Medical College*, Bangladesh. 2009;5(2):24–8. 10.3329/jafmc.v5i2.4579

130. Goswami B, Nanda V, Kataria S, Kataria D. Prediction of In-Hospital Mortality in Patients With Traumatic Brain Injury Using the Rotterdam and Marshall CT Scores: A Retrospective Study From Western India. Cureus. 2023;15(7). 10.7759/cureus.41548

131. Khan KA, Choudhary M, Sinha VD, Gora N, Bairwa M. Predictors of Outcome After Traumatic Brain Injuries: Experience of a Tertiary Health Care Institution in Northwest India. World Neurosurgery. 2019;126:e699–705. 10.1016/j.wneu.2019.02.126

132. Regmi M, Bhatta OP, Sharma MR. Pre-hospital care, pre-hospital delay, and in-hospital delay in patients with traumatic brain injury in getting neurosurgical care in a tertiary care center: A Cross-Sectional study. J Nepal Med Assoc. 2024;62(275):416–20. 10.31729/jnma.8629

133. Ram K, VaraPrasad K, Krishna MK, et al. Prehospital Factors Associated with Discharge Outcomes: Baseline Data from the Andhra Pradesh Traumatic Brain Injury Project. World Neurosurgery: X. 2019;2:100020. 10.1016/j.wnsx.2019.100020

134. Kar M, Sahu C, Singh P, et al. Prevalence of Traumatic Brain Injury and Associated Infections in a Trauma Center in Northern India. Journal of Global Infectious Diseases. 2023;15(4):137–43. 10.4103/jgid.jgid_66_23

135. Agrawal D, Singh PK, Sinha S, Gupta DK, Satyarthee GD, Misra MC. Remaining unconscious: The burden of traumatic brain injuries in India. Journal of Neurosciences in Rural Practice. 2015;06(04):520–2. 10.4103/0976-3147.165394

136. Kumar A, Jaiswal S, Ojha BK, et al. Retrospective Outcome Analysis of Geriatric Traumatic Brain Injury Treated at a Tertiary Care Center in India. Indian Journal of Neurosurgery. 2025;14(01):034–43. 10.1055/s-0044-1788254

137. Churiwala J, Garale MN, Kawale J, Dandpat SK, Mahore A. Risk factors of deterioration in patients of head injury with non-operative management on first neurosurgical consultation. JNRP. 2022;14:28–34. 10.25259/JNRP-2022-1-41-R2

138. Paudel SS, Luitel R, Bista A, et al. Scenario of Head Injury Patients in Tertiary Care Hospital of Nepal. J Nepal Health Res Counc. 2020;18<otherinfo>(1)</otherinfo>:112–5. 10.33314/jnhrc.v18i1.2276

139. Kannan S, Marudachalam KS, Puri GD, Chari P. Severe head injury patients in a multidisciplinary ICU: are they a burden? Intensive Care Med. 1999;25(8):855–8. 10.1007/s001340050965

140. Jain N, Varun A, Mishra PK, Tomar JS. Socio-demographic profile of head injury victims in road traffic accidents, an autopsy based study at SAMC & PGI, Indore. IJFCM. 2020;7(2):56–60. 10.18231/j.ijfcm.2020.014

141. Satapathy M, Dash D, Mishra S, Tripathy S, Nath P, Jena S. Spectrum and outcome of traumatic brain injury in children <15 years: A tertiary level experience in India. Int J Crit Illn Inj Sci. 2016;6(1):16. 10.4103/2229-5151.177359

142. Pusdekar V, Ambedkar S, Bodade R. Study of cases of head injury in a government hospital in rural Indian setting. Int Surg J. 2018;5(10):3252. 10.18203/2349-2902.isj20184071

143. Dade NB, Teegala R, Medikonda M. Study of Epidemiology of Traumatic Brain Injury and Prevalence of Psychiatric Disorders in Traumatic Brain Injury at 3 Months Follow-Up. Nep J Neurosci. 2019;16(2):8–15. 10.3126/njn.v16i2.25939

144. Patil NJ, Hulwan AB, Kadam RS, Kumar S, Vadhel CR. Study of Head Trauma through Computed Tomography. Journal of Pharmacy and Bioallied Sciences. 2024;16(Suppl 1):S415–7. 10.4103/jpbs.jpbs_636_23

145. Iyer S, Patel G. Study of risk factors, clinical spectrum, and outcome for head injury in pediatric age group in Western India. Afr J Paediatr Surg. 2020;17(1):26. 10.4103/ajps.AJPS_2_18

146. Deotale S, Mukhamale V, Thakur BA, Mudgerikar R. The audit of traumatic head injuries in a tertiary care hospital in Mumbai: a retrospective study. Int Surg J. 2020;7(11):3678. 10.18203/2349-2902.isj20204672

147. Tabish A, Lone NA, Afzal WM, Salam A. The incidence and severity of injury in children hospitalised for traumatic brain injury in Kashmir. Injury. 2006;37(5):410–5. 10.1016/j.injury.2006.01.039

148. Yattoo G, Tabish A. The profile of head injuries and traumatic brain injury deaths in Kashmir. J Trauma Manage Outcomes. 2008;2(1):5. 10.1186/1752-2897-2-5

149. Agrawal A, Munivenkatappa A, Rustagi N, Rammohan P, Subrahmanyam B. Time of admission and outcome in traumatic brain injury patients. Med J DY Patil Univ. 2016;9(4):465. 10.4103/0975-2870.186064

150. Pareek K, Sodhi D. To study epidemiological aspects of acute head trauma at tertiary care center Bikaner Rajasthan. IJMBS. 2020;4(1). 10.32553/ijmbs.v4i1.1048

151. Mr H. Traumatic Brain Injuries and its Outcome at a Tertiary Care Hospital in Northwest Part of Bangladesh. Mymensingh Med J. 2024; 33(4):1230–1237. 10.XXXXX/mmj.2024.v3304.38

152. Rahman U, Hamid M, Shan Dasti M, Nouman T, Vedovelli L, Javid A. Traumatic Brain Injuries: A Cross-Sectional Study of Traumatic Brain Injuries at a Tertiary Care Trauma Center in the Punjab, Pakistan. Disaster med public health prep. 2023;17:e89. 10.1017/dmp.2021.361

153. Umeranı MS, Abbas A, Sharıf S. Traumatic Brain Injuries: Experience from a Tertiary Care Center in Pakistan. Turkish Neurosurgery. 2014;24(1). 10.5137/1019-5149.jtn.7080-12.1

154. Shrestha A, Paudel N, Adhikari G, et al. Traumatic Brain Injury among Patients Admitted in Neurosurgical Unit in a Tertiary Care Centre: A Descriptive Cross-sectional Study. J Nepal Med Assoc. 2023;61(262):514–8. 10.31729/jnma.8197

155. Ashraf M, Kamboh UA, Hussain SS, et al. Traumatic Brain Injury in Underage Motorcycle Drivers: Clinical Outcomes and Sociocultural Attitudes from a Lower-Middle-Income Country. World Neurosurgery. 2022;167:e413–22. 10.1016/j.wneu.2022.08.027

156. Ahmed S, Anwer A, Abdullah M, et al. Trends in Traumatic Brain Injuries During the COVID-19 Pandemic: A Single-Center Review of Patient Charts From Pakistan. Cureus. 2024;16(4): e58745. 10.7759/cureus.58745

157. H. K, Paparajamurthy P, K. M, K. A. A clinical analysis of outcome in management of head injury in patients with highway road accidents. Int J Res Med Sci. 2016;2079–83. 10.18203/2320-6012.ijrms20161764

158. Hazen A, Ehiri JE. Road traffic injuries: hidden epidemic in less developed countries. J Natl Med Assoc. 2006;98(1):73–82.

159. Karamian A, Lucke-Wold B, Seifi A. Prevalence of Traumatic Brain Injury in the General Adult Population of the USA: A Meta-Analysis. Neuroepidemiology. 2024;1–10. 10.1159/000540676

160. Ye Z, Li Z, Zhong S, et al. The recent two decades of traumatic brain injury: a bibliometric analysis and systematic review. Int J Surg. 2024;110(6):3745–59. 10.1097/JS9.0000000000001367

161. Brazinova A, Rehorcikova V, Taylor MS, et al. Epidemiology of Traumatic Brain Injury in Europe: A Living Systematic Review. Journal of Neurotrauma. 2021;38(10):1411–40. 10.1089/neu.2015.4126

162. van Wessem KJP, Benders KEM, Leenen LPH, Hietbrink F. TBI related death has become the new epidemic in polytrauma: a 10-year prospective cohort analysis in severely injured patients. Eur J Trauma Emerg Surg. 2024;50(6):3083–94. 10.1007/s00068-024-02653-1

163. Abate SM, Abafita BJ, Bekele T. Prevalence of Traumatic Brain Injury Among Trauma Patients in Ethiopia: Systematic Review and Meta-Analysis. Annals of African Surgery. 2021;18(1):10–7. 10.4314/aas.v18i1.3

164. Dash DP, Sethi N, Dash AK. Identifying the causes of road traffic accidents in India: An empirical investigation. Journal of Public Affairs. 2020;20(2):e2038. 10.1002/pa.2038

165. Gyawali S, Shah R, Mandal N. Pattern and severity of injuries, risk factors and outcome of Road Traffic Accident victims attending emergency department of Birat Medical College Teaching Hospital. Birat J Health Sci. 2024;9(1):104–8. 10.62065/bjhs549

166. Hamid S, Davoud KZ. Road traffic injuries measures in the Eastern Mediterranean Region: findings from the Global Status Report on Road Safety – 2015. J Inj Violence Res. 2019;11(2):149–58. 10.5249/jivr.v11i2.1122

167. Shafiq S, Dahal S, Siddiquee NKA, Dhimal M, Jha AK. Existing Laws to Combat Road Traffic Injury in Nepal and Bangladesh: A Review on Cross Country Perspective. J Nepal Health Res Counc. 2020;17(4):416–23. 10.33314/jnhrc.v17i4.2363

168. Popescu C, Anghelescu A, Daia C, Onose G. Actual data on epidemiological evolution and prevention endeavours regarding traumatic brain injury. J Med Life. 2015;8(3):272–7.

169. Maleki MS, Mazaheri SA, Hosseini SH, et al. Epidemiology of Traumatic Brain Injury in Iran: A Systematic Review and Meta-Analysis. Iran J Public Health. 2023;52(9):1818–31. 10.18502/ijph.v52i9.13565

170. Iderdar Y, Arraji M, Wachami NA, et al. Predictors of outcomes 3 to 12 months after traumatic brain injury: a systematic review and meta-analysis. Osong Public Health Res Perspect. 2024;15(1):3–17. 10.24171/j.phrp.2023.0288

171. Moher D, Shamseer L, Clarke M, et al. Preferred reporting items for systematic review and meta-analysis protocols (PRISMA-P) 2015 statement. Revista Espanola de Nutricion Humana y Dietetica. 2016;20(2):148–60. 10.1186/2046-4053-4-1

