## Supplementary material for "Epidemiology of traumatic brain injury in South Asia: A Systematic Review": S1 Appendix: Search String

(“Trauma” OR "head injury" OR "head injuries" OR "brain injury" OR "brain injuries" OR "traumatic brain injury" OR "TBI" OR "craniocerebral injuries" OR "craniocerebral injury" OR "head trauma" OR "brain trauma" OR "cranial trauma" OR "brain laceration" OR "brain contusion")

AND

("epidemiology" OR "prevalence" OR "Proportion" OR "incidence" OR "mortality" OR "case fatality" OR "severity" OR "risk factor" OR "mechanism" OR "outcomes" OR "morbidity" OR "impact")

AND

("South Asia" OR Bangladesh OR India OR Pakistan OR Afghanistan OR Sri Lanka OR Nepal OR Bhutan OR Maldives)

AND

("hospital" OR "trauma center" OR "emergency" OR "fatalities" OR "hospitalized" OR "inpatient") AND ("retrospective study" OR "prospective study" OR "cross-sectional" OR "cohort study" OR "case-control" OR "descriptive study")
