## Supplementary material for "Epidemiology of traumatic brain injury in South Asia: A Systematic Review": S2 Appendix: Methodological Evaluation of Observational Research (MORE) Checklist for Quality Assessment of Included Studies

| **Type** | **Criteria** | | **Conditions** | **Assessment** |
| --- | --- | --- | --- | --- |
| General descriptive | Aim of study | | a) Included incidence/prevalence estimation with clear target population b) Included incidence/prevalence estimation without clear target population c) Aim of study was not stated d) Unclear due to poor reporting | a) OK b) Minor ﬂaw c) Major ﬂaw d) Poor reporting |
|  | Funding of study | | a) Reported b) Not reported | a) OK b) Poor reporting |
|  | Conﬂict of interest | | a) Reported b) Not reported | a) OK b) Poor reporting |
|  | Ethical approval | | a) Reported b) Not reported | a) OK b) Poor reporting |
|  | Study design | | a) Reported b) Not reported | a) OK b) Poor reporting |
| External  validity | Sampling | Sampling frame | a) Not applicable for study design b) Sampling within nationally representative registries or databases c) Medical records d) Insurance claims e) Outpatients contacts f) Unclear due to poor reporting | a) NA b) OK c) Minor flaw d) Minor flaw e) Major flaw f) Poor reporting |
|  | Definition  of cases | Validation | a) Criteria of TBI were stated according to the guidelines b) TBI were defined by ICD codes c) Criteria of TBI were not stated according to the guidelines d) Criteria of TBI were not stated e) Unclear due to poor reporting | a) OK b) OK c) Minor flaw d) Major flaw e) Poor reporting |
|  |  | Severity of TBI | a) Stated in the study with scales b) Stated in the study without scales c) Not stated in the study d) Unclear due to poor reporting | a) OK b) Minor flaw c) Major flaw d) poor reporting |
|  | Address bias | Sampling bias is  addressed  in the analysis  (can by  mentioned in  recommendations) | a) Not applicable for study design b) Weighting of the estimates by probability of selection c) Weighting of the estimates by non-response adjustment within sampling subgroups d) Post-stratification by age e) Post-stratification by sex f) Mentioned, but not addressed in analysis g) Not mentioned and not addressed in analysis h) Unclear due to poor reporting | a) NA b) OK c) OK d) OK e) OK f) Minor flaw g) Major flaw h) Poor reporting |
|  |  | Subject flow | a) Not applicable for study design b) Number of enrolled stated clearly c) Number of enrolled not stated clearly d) Number of enrolled not stated e) Unclear due to poor reporting | a) NA b) OK c) Minor flaw d) Major flaw e) Poor reporting |
| Internal  validity | Reporting of  methods | Source of data | a) Objectively measured with diagnostic methods for the purpose of the study (independent on health care) b) Measured by interviewers for the study c) Obtained during clinical examination for the purpose of the study d) Obtained from registries or administrative databases (collected for epidemiologic evalua- tion independent of health care) e) Obtained from medical records (mining of the data collected for health care purposes) f) Obtained from administrative database (mining of the data collected for health care purposes) g) Source of data is not stated in the study h) Unclear due to oor re ortin | a) OK b) OK c) OK d) OK e) Minor flaw f) Minor flaw g) Major flaw h) Poor reporting |
|  |  | Reliability of  the estimates | a) Methods of data analysis (nominators/denominators) are stated clearly b) Methods of data analysis (nominators/denominators) are stated, but not clearly c) Methods of data analysis (nominators/denominators) are not stated d) Unclear due to poor reporting | a) OK b) Minor flaw c) Major flaw d) Poor reporting |
|  | Reporting of  estimates | Incidence | a) Incidence type (cumulative incidenc, incidencerate) stated and counted clearly b) Incidence type (cumulative incidenc, incidencerate) stated and counted not clearly c) Incidence type (cumulative incidenc, incidencerate) not stated and not counted d) Unclear due to poor reporting | a) OK b) Minor flaw c) Major flaw d) Poor reporting |
|  |  |  | a) Precision of estimation (error, 95% CI) reported in all b) Precision of estimation (error, 95% CI) reported not in all c) Precision of estimation (error, 95% Cl) omitted d) Unclear due to poor reporting | a) OK b) Minor flaw c) Major flaw d) Poor reporting |
|  |  |  | a) Age adjusted incidence in total sample b) Crude incidence in total sample c) The incidence is not stated as age adjusted or crude d) Unclear due to poor reporting | a) OK b) Minor flaw c) Major flaw d) Poor reporting |
|  |  |  | a) Age adjusted incidences in all population subgroups b) Age adjusted incidences not in all population subgroups c) Crude incidences in subgroups d) Incidences are not stated as age adjusted or crude e) Unclear due to poor reporting | a) OK b) Minor flaw c) Minor flaw d) Major flaw e) Poor reporting |
|  |  | Mortality | a) Mortality in all population is counted from all population b) Mortality in all population is counted not clearly c) Mortality in all population is counted as case fatality rate (only from the cases not from all population) d) Unclear due to poor reporting | a) OK b) Minor flaw c) Major flaw d) Poor reporting |
|  |  |  | a) Precision of estimation (error, 95% CI) reported in all b) Precision of estimation (error, 95% CI) reported not in all c) Precision of estimation (error, 95% Cl) omitted d) Unclear due to poor reporting | a) OK b) Minor flaw c) Major flaw d) Poor reporting |
|  |  |  | a) Age adjusted mortality in total sample b) Crude mortality in total sample c) The mortality is not stated as age adjusted or crude d) Unclear due to poor reporting | a) OK b) Minor flaw c) Major flaw d) Poor reporting |
|  |  |  | a) Age adjusted mortalities in all population subgroups b) Age adjusted mortalities not in all population subgroups c) Crude mortalities in subgroups d) Mortalities are not stated as age adjusted or crude e) Unclear due to poor reporting | a) OK b) Minor flaw c) Minor flaw d) Major flaw e) Poor reporting |
